# Real-world comparative effectiveness of mRNA-1273 and BNT162b2 vaccines among immunocompromised adults in the United States

**DOI:** 10.1101/2022.05.13.22274960

**Authors:** Katherine E. Mues, Brenna Kirk, Deesha A. Patel, Alice Gelman, Scott Chavers, Carla Talarico, Daina B. Esposito, David Martin, James Mansi, Xing Chen, Nicolle M. Gatto, Nicolas Van de Velde

## Abstract

**Introduction:** Head-to-head studies comparing COVID-19 mRNA vaccine effectiveness in immunocompromised individuals, who are vulnerable to severe disease are lacking, as large sample sizes are required to make meaningful inferences.

**Methods:** This observational comparative effectiveness study was conducted in closed administrative claims data from the US HealthVerity database (December 11, 2020-January 10, 2022, before omicron). A 2-dose mRNA-1273 versus BNT162b2 regimen was assessed for preventing medically-attended breakthrough COVID-19 diagnosis and hospitalizations among immunocompromised adults. Inverse probability of treatment weighting was applied to balance baseline characteristics between vaccine groups. Incidence rates from patient-level data and hazard ratios (HRs) using weighted Cox proportional hazards models were calculated.

**Results:** Overall, 57,898 and 66,981 individuals received a 2-dose regimen of mRNA-1273 or BNT161b2, respectively. Among the weighted population, mean age was 51 years, 53% were female, and baseline immunodeficiencies included prior blood transplant (8%-9%), prior organ transplant (7%), active cancer (12%-13%), primary immunodeficiency (25%-26%), HIV (20%-21%), and immunosuppressive therapy use (60%-61%). Rates per 1,000 person-years (PYs; 95% confidence intervals [CI]s) of breakthrough medically-attended COVID-19 were 25.82 (23.83-27.97) with mRNA-1273 and 30.98 (28.93, 33.18) with BNT162b2 (HR, 0.83; 95% CI, 0.75-0.93). When requiring evidence of an antigen or polymerase chain reaction test before COVID-19 diagnosis, the HR for medically-attended COVID-19 was 0.78 (0.67-0.92). Breakthrough COVID-19 hospitalization rates per 1,000 PYs (95% CI) were 3.66 (2.96-4.51) for mRNA-1273 and 4.68 (3.91-5.59) for BNT162b2 (HR, 0.78; 0.59-1.03). Utilizing open and closed claims for outcome capture only, or both cohort entry/outcome capture, produced HRs (95% CIs) for COVID-19 hospitalization of 0.72 (0.57-0.92) and 0.66 (0.58-0.76), respectively.

**Conclusions:** Among immunocompromised adults, a 2-dose mRNA-1273 regimen was more effective in preventing medically-attended COVID-19 in any setting (inpatient and outpatient) than 2-dose BNT162b2. Results were similar for COVID-19 hospitalization, although statistical power was limited when using closed claims only.

**Study Registration:** NCT05366322

## Introduction

As of April 26, 2022, in the United States, severe acute respiratory syndrome coronavirus 2 (SARS-CoV-2) has caused more than 80.2 million confirmed coronavirus 2019 (COVID-19) cases and more than 983,000 deaths [1], with actual cases estimated to be 5 to 50 times greater than those reported and incidence fluctuating continuously [1, 2]. Comprising ∼3% of the adult US population, immunocompromised individuals are especially vulnerable to COVID-19, with greater risk of serious or prolonged illness [3, 4].

Three COVID-19 vaccines are available in the United States: mRNA-1273 (Spikevax^®^; Moderna, Inc., Cambridge, MA), BNT162b2 (COMIRNATY^®^; BioNTech, Mainz Germany, and Pfizer Inc, New York, NY), and Ad26.COV2.S (Janssen Biotech, Inc, Horsham, PA). mRNA-1273 has been authorized by the US Food and Drug administration (FDA) as a 2-dose series (100-μg mRNA per dose) in individuals aged ≥18 years [5, 6]; BNT162b2 has also been authorized by the FDA as a 2-dose series (30-μg mRNA per dose) in individuals aged ≥16 years and authorized under Emergency Use Authorization (EUA) as a 2-dose series in individuals aged 5 to 15 years [7-9]; and Ad26.COV2.S has been authorized under EUA as a single-dose (5×10^10^ virus particles per dose) in individuals aged ≥18 years [10]. Booster doses have also been authorized under EUA, administered ≥5 months after the primary series as a half dose for mRNA-1273 (50 μg) and a full dose for BNT162b2 (30 μg) and ≥2 months after the primary dose for Ad26.COV2.S [5, 7, 10, 11]. While all 3 vaccines are approved for use in the United States, the Advisory Committee on Immunization Practice (ACIP) and the Centers for Disease Control and Prevention (CDC) advise that the use of an mRNA vaccine is recommended over Ad26.COV2.S due to greater vaccine effectiveness (VE) and lower risk of serious adverse events (ie, thrombosis with thrombocytopenia syndrome [TTS]) [12, 13].

Although high vaccine efficacy was reported in phase 3 trials of mRNA-1273 and BNT162b2 in the general population, these trials excluded immunocompromised individuals, including those with underlying immunocompromising conditions and those prescribed immune-modifying therapies (with the exception of individuals with stable HIV and very low viral load[14-16]. Subsequent real-world data in the vaccinated immunocompromised population showed attenuated VE, with a higher risk of infection, hospitalization, death, persistent infection and shedding, viral evolution, reduced antibody and neutralization titers, and infection of household contacts [17]. Given these increased risks, a third and fourth dose of mRNA-based vaccines have been recommended for this vulnerable group [17-19]. An additional primary dose of mRNA-1273 (100 μg; individuals aged ≥18 years) or BNT162b2 (30 μg; individuals aged ≥12 years) is authorized for administration ≥1 month after completion of the primary series in moderately to severely immunocompromised individuals [5, 7]. For moderately to severe immunocompromised individuals, the CDC recommends 1 mRNA-based booster be given ≥3 months after an mRNA-based primary vaccination series and ≥2 months after an Ad26.COV2.S primary dose [20].

Head-to-head studies comparing the VE of the COVID-19 vaccines in immunocompromised individuals have not been conducted to date; however, numerous studies have investigated antibody-mediated immunogenicity in response to vaccination and reported a more robust immune response to mRNA-1273 compared to BNT162b2 in this vulnerable cohort [3, 17, 21-24]. It is hypothesized that these differences in immunogenicity between mRNA vaccines may translate into differences in VE against COVID-19 infections and hospitalizations in immunocompromised individuals. One observational study of immunocompromised adults vaccinated with 2 doses of an mRNA COVID-19 vaccine reported VE against COVID-19 hospitalizations to be 81% (95% CI, 76%-85%; n=4,337) with mRNA-1273, and 71% (95% CI, 65%-76%; n=6,227) with BNT162b2; however, direct comparison of vaccines in the immunocompromised population cannot be made, as the test-negative study design was not intended for head-to-head statistical comparisons [25].

In this study, we compared the real-world VE specifically of 2-dose mRNA-based vaccines, mRNA-1273 and BNT1262b2, against medically-attended breakthrough COVID-19 diagnosis and COVID-19 hospitalizations in immunocompromised adults (NCT05366322).

## Methods

### Objectives

The primary objective was to compare the real-world VE of 2 doses of mRNA-1273 versus 2 doses of BNT162b2 against medically-attended breakthrough COVID-19 diagnosis among immunocompromised adults. The secondary objective was to compare the RWE of 2 doses of mRNA-1273 versus 2 doses of BNT162b2 vaccine against breakthrough COVID-19 hospitalizations among immunocompromised adults.

### Study Design and Data Source

This observational comparative VE cohort study utilized medical and pharmacy claims data from December 11, 2020, through January 10, 2022 (available time period in the data when the delta variant dominated COVID-19 infection) aggregated by HealthVerity (**Figure 1**). HealthVerity data was drawn from a variety of US sources, including 2 closed claims databases (Private Source 17 and Private Source 20) and numerous open claims data sources, with data elements including provider-submitted claims, adjudicated insurance claims, and pharmacy billing manager claims submissions. These sources include individuals insured under commercial, Medicare (limited available claims data) or Medicaid plans, or providers participating in several large medical and pharmacy insurance claims submission systems (eg, medical/pharmacy claims clearinghouses). The primary analysis was limited to closed medical claims only and data were truncated 3 months (to October 12, 2021) from the most recently available data at the time of the analysis to allow for claims adjudication (**Table S1**).

**Figure 1.**
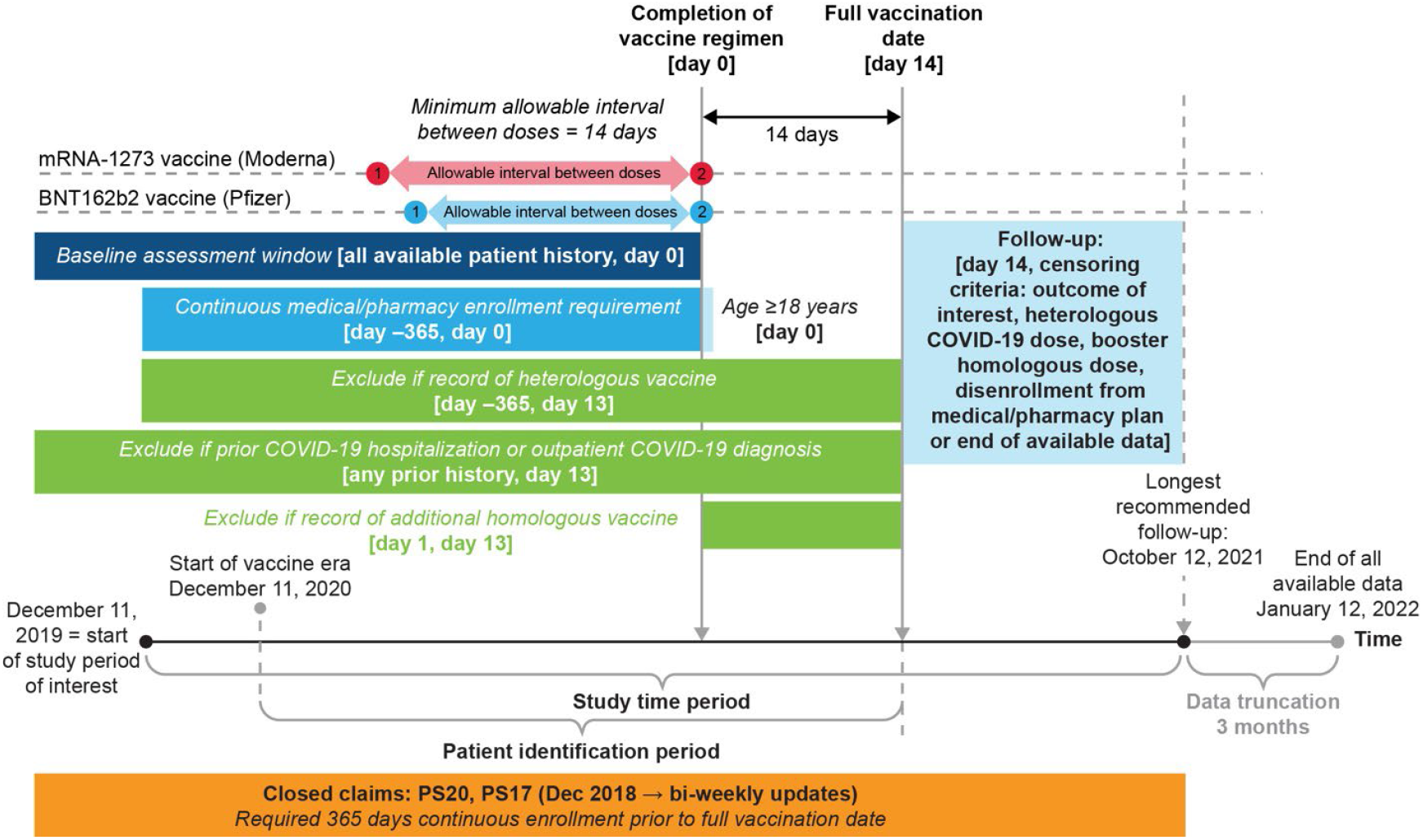
Study design schema. Immunocompromised adults who had completed a 2-dose homologous vaccine regimen were identified via central procedural terminology (CPT) and nation drug codes (NDC) from December 11, 2020, through January 10, 2022.

The HealthVerity database includes demographic variables such as age, sex, and 3-digit zip level; race and ethnicity are not available. Hospitalizations are included in the data at a summary level. Open claims are sourced directly through medical clearinghouses and pharmacy benefit managers; there is no associated enrollment file, resulting in incomplete capture of healthcare system interactions. Closed claims represent claims accepted by and paid by health insurance companies and generally lagged 3 to 6 months to allow for full adjudication.

To create linkages across data feeds and ensure de-identified, longitudinal, de-duplicated patient data, all data partners use the HealthVerity technology within their system to create a unique, secure, encrypted, and non-identifiable patient token that is usable across datasets. Use of data and the precise granularity available is controlled by Health Insurance Portability and Accountability Act (HIPAA) requirements or application of public health exemption. No protected health information or personal identifying information leaves the data owner’s possession, and all research data were certified HIPAA compliant by expert determination.

The study follows the Guidelines for Good Epidemiologic Practice practices laid out in 2005 US FDA Good Pharmacy Practice and the 2008 International Society of Pharmacoepidemiology Good Pharmacy Practice [26, 27]. This study was exempt from review by the New England Institutional Review Board (#1-9757—1). All participant data were de-identified, and all participant-level and provider-level data within the database contained synthetic identifiers to protect the privacy of individuals and data contributors.

### Vaccine Groups

Vaccinations were captured in the database via manufacturer-specific current procedural terminology (CPT) and national drug code (NDC; **Table S2**). Drugs dispensed by a pharmacy were captured, while over-the-counter medications and those provided during an inpatient stay were not. A 2-dose regimen was defined as 2 doses of mRNA-1273 (exposure group) or BNT1262b2 (referent group). An exploratory analysis of the distribution of days between administration of 2 doses of vaccine found the fifth percentile for mRNA-1273 or BNT162b2 to be 25 days and 18 days, respectively. Given these results and the aim of inclusivity with respect to dose 2 in this high-risk population, a minimum of 14 days between doses was required between receiving 2-dose mRNA-1273 or BNT162b2 vaccines The index date was defined as the date of vaccine regimen completion (ie, date of second dose of mRNA-1273 or BNT1262b2).

### Population and Follow-Up

Individuals aged ≥18 years who received 2 doses of mRNA-1273 or BNT162b2 and were continuously enrolled in a medical and pharmacy plan for 365 days before the index date were included in the analysis (**Figure 1)**.

Immunocompromised individuals were identified using an adapted claims-based algorithm aligning with the CDC definition of immunocompromised individuals eligible for COVID-19 vaccine booster doses (**Table S2**)[28]. Immunocompromised individuals were identified as those meeting at least one of the following criteria: evidence of blood or stem cell transplant 2 years before the index date; history of organ transplant and taking immunosuppressive therapy 60 days before the index date; evidence of active cancer treatment 180 days before the index date with an active cancer diagnosis in the year before treatment; any history of primary immunodeficiency disorder; or any history of an HIV.

Exclusion criteria included evidence of COVID-19 infection before the index date or in the 13 days following the index date, receipt of a heterologous COVID-19 vaccine in the 365 days before or in the 13 days following the index date, receipt of additional dose of homologous COVID-19 vaccine in the 13 days following the index date, or missing or unknown sex on the index date. Additional exclusion details are presented in the **Supplementary text**.

The baseline period was defined as the 365 days before the index date. Participants were followed from 14 days following completion of a 2-dose vaccine regimen until an outcome of interest, receipt of a third COVID-19 vaccine dose (heterologous or homologous), disenrollment from a medical/pharmacy plan, or October 12, 2021, whichever occurred first.

### Study Outcomes

The primary outcome of medically attended breakthrough COVID-19 diagnosis was defined as a claim with the ICD-10 diagnostic code for COVID-19 (U07.1) in any setting, including inpatient, outpatient, emergency department, and urgent care. The secondary outcome of breakthrough hospitalization for COVID-19 was defined as a hospital stay with the ICD-10 diagnostic code for COVID-19 (U07.1) listed as the primary diagnosis or within 21 days prior to hospital admission.

A phased approach was used, including an exploratory phase to inform key design decisions (see details in study protocol) and a diagnostic phase to ensure baseline balance and positivity were achieved and to assess the existence of non-differential censoring. Once the pre-specified diagnostic criteria were met, the association between vaccine group and the study outcomes was assessed in the inferential phase during which the outcome models were implemented. Further details are presented in the **Supplementary text**.

### Statistical Analysis

The distributions of baseline variables within each vaccine cohort were described as number and percentage for categorical variables and as mean (standard deviation), median (interquartile range), and range (minimum, maximum) for continuous variables. Baseline variables hypothesized to be confounding variables were included in a propensity score (PS) model; age, sex, payer type, state of residence, and calendar time were assessed at the index date. Healthcare resource utilization, the Charlson comorbidity score, frailty score, and number of unique immunosuppressive therapies used were assessed in the 365 days before the index date. Individual comorbid conditions were assessed using all prior claims available. The algorithms used to identify immunocompromised adults are presented in **Table S3**, and a full list of baseline covariates of interest are presented in **Table S4**. Inverse probability of treatment weights (IPTW) was calculated as 1/PS for participants in the mRNA-1273 group (exposed) and as 1/(1-PS) for participants in the BNT162b2 group (referent). Exposure and referent groups were considered balanced if the absolute standardized differences (ASDs) for all baseline covariates used to generate the PS were <0.10 [29-31].

Incidence rates per 1,000 PYs with corresponding 95% confidence intervals (CIs) were estimated for each treatment group. Kaplan–Meier plots with 95% CIs and Schoenfeld residuals were generated to assess comparative VE over time. Weighted Cox proportional hazard models were executed to estimate hazard ratios (HRs) and corresponding 95% CIs. Subgroup analyses were implemented for age categories, calendar quarter of vaccine receipt, COVID-19 transmission level at index date, and immunocompromised subgroup.

Analyses were conducted using the Aetion Evidence Platform^®^, software for real-world data analysis, which has been scientifically validated for observational cohort studies using large healthcare databases [32]. Transformations of the raw data are preserved for full reproducibility and audit trails are available, including a quality check of the data ingestion process.

### Sensitivity Analyses

Several per-protocol sensitivity analyses were conducted within this study (**Table S1**). These included evaluating the impact of the adjudication lag in closed claims, where the primary analysis was implemented through January 10, 2022, without truncation. Another sensitivity analysis used an alternative definition of medically-attended breakthrough COVID-19 diagnosis that required evidence of a rapid antigen test on the same day or 3 days before the COVID-19 diagnosis code or a polymerase chain reaction test 3 to 14 days before the COVID-19 diagnosis code.

In addition, analyses were conducted utilizing both closed and open claims for cohort entry and outcomes. As open claims are not associated with an enrollment file, individuals without an enrollment file and identified via open claims only were required to have at least one open medical and open pharmacy claim in the 365 days before the index date. During follow-up, individuals without an enrollment file or those who disenrolled from their medical/pharmacy plan continued to be followed-up if there was evidence of a medical or pharmacy claim every 60 days. Open claims with a diagnosis code for COVID-19 were incorporated into the primary and secondary outcomes.

A post hoc sensitivity analysis was performed, which required individuals to have an enrollment file on the index date, with continuous enrollment in the 365 days before and continuous enrollment during follow up, while allowing for capture of the outcomes of interest utilizing both closed and open claims.

## Results

A total of 124,879 immunocompromised adults identified through HealthVerity database records from December 11, 2020, through October 12, 2021, were included in the primary analysis; 57,898 completed a 2-dose vaccination with mRNA-1273 and 66,981 completed a 2-dose vaccination with BNT162b2 (**Figure 2**). Prior to weighting, compared to the BNT162b2 group, individuals in the mRNA-1273 group were slightly older (mean ages of 50 and 52 years, respectively) and there were slightly less women in the population (53% and 52%, respectively; **Table S5**) before weighting. The distribution of individuals in California, New York, and Pennsylvania was higher for the mRNA-1273 group, while BNT162b2 was more represented in the distribution of individuals in Illinois, New Jersey, and Texas. Those in the mRNA-1273 vaccine group had a slightly higher frequency of clinical comorbidities (**Table S5**).

**Figure 2.**
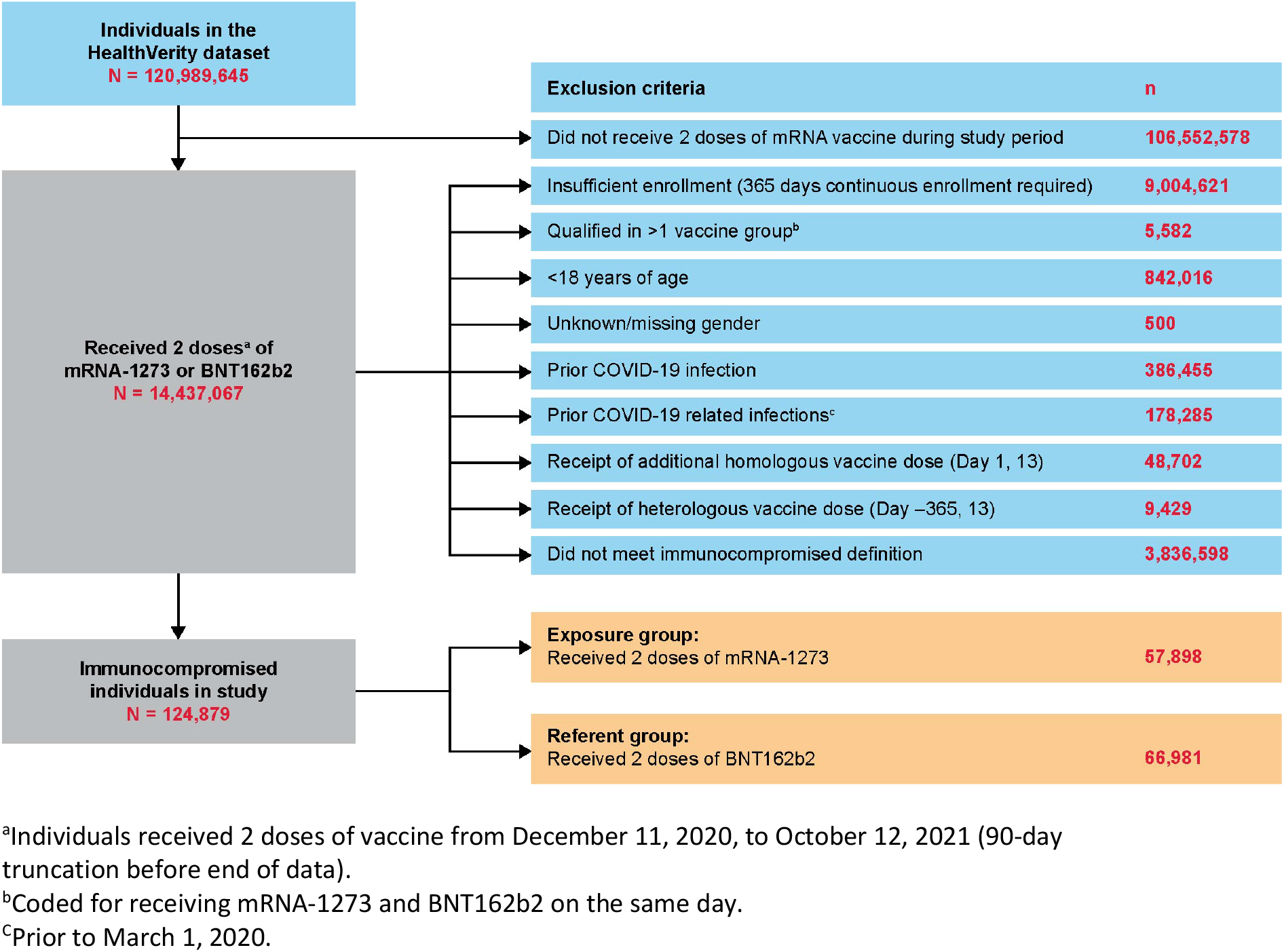
Participant attrition diagram. Participant attrition flow diagram using claims from the HealthVerity database. Numbers represent the patient size before inverse probability of treatment weighting.

Inverse probability of treatment weighting resulted in a pseudopopulation of 124,588 mRNA-1273 individuals and 124,313 BNT162b2 individuals (**Table S7**). Following weighting, exposure and referent groups were considered balanced, as ASDs for all baseline covariates were <0.10. The mean age of the participants was 51 years, 53% were female, and participants were primarily insured by commercial insurance (62%) or Medicaid (32%) (**Table 1**). Texas (16%), Illinois (13%), California (9%), New Jersey (8%), and Michigan (6%) were the top 5 represented states (**Table S5**). Clinical history was balanced between the vaccine groups with respect to a history of arrythmia (25%), prior cardiovascular disease (64%), hypertension (52%), prior heart failure (7%), chronic lung disease (23%), obesity (32%), diabetes (22%), and irritable bowel syndrome (47%). Within this immunocompromised population, 8% to 9% had received a prior blood transplant, 7% had a prior organ transplant, 12% to 13% had a cancer diagnosis with active treatment, 25% to 26% had a primary immunodeficiency, 20% to 21% had HIV, and 60% to 61% used an immunosuppressive therapy 60 days before the index date (**Table 1**).

**Table 1.**
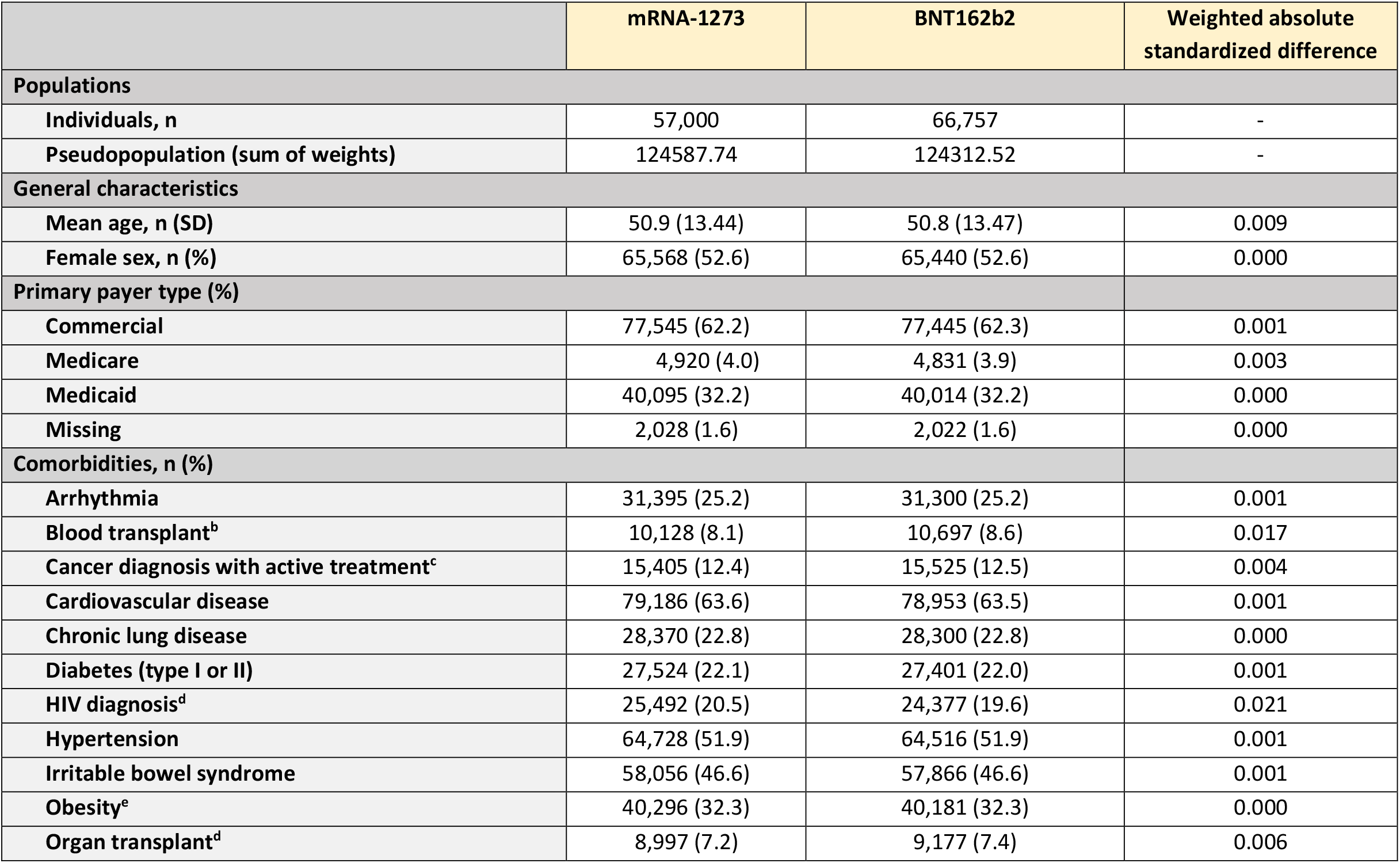
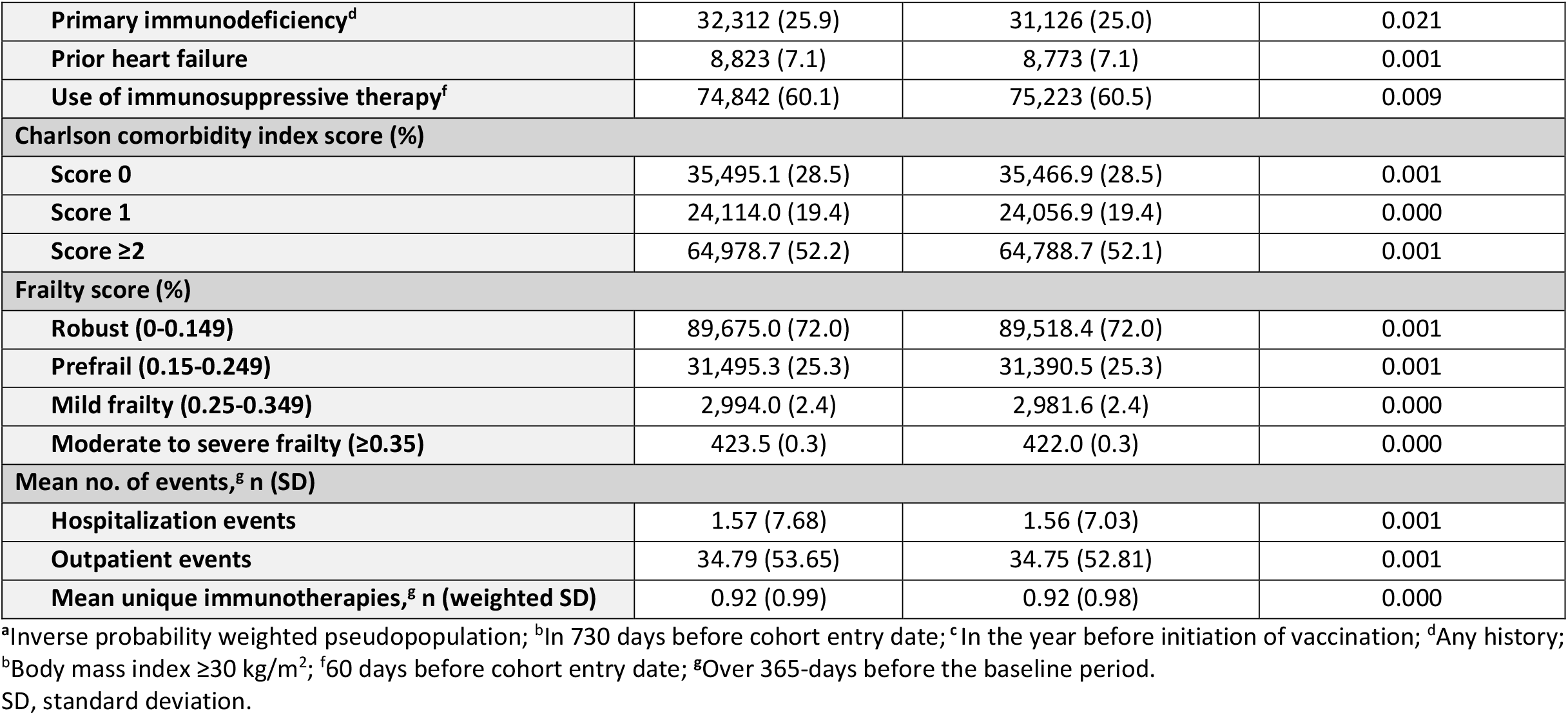
Pseudopopulation baseline characteristics of immunocompromised adults with a 2-dose regimen of mRNA-1273 or BNT162b2^a^.

Results from the primary analysis showed that the overall rate of medically-attended COVID-19 was lower among individuals who received 2 doses of mRNA-1273 (25.82 per 1,000 PYs; 95% CI, 23.83-27.97) compared those who received 2 doses of BNT162b2 (30.98 per 1,000 PYs; 95% CI, 28.93-33.18; HR, 0.83; 95% CI, 0.75-0.93) (**Table 2; Figure 3A**). In subgroup analyses, results were similar across categories of transmission level, calendar quarter of completion of the 2-dose regimen, and age, although the small numbers of individuals in the subgroups of those who received second doses administered in Q3 2021 and individuals ≥65 years, limited interpretation of these results (**Table S8**). Among immunocompromised subgroups, the rate of medically-attended COVID-19 was lower with mRNA-1273 than BNT162b2 vaccination for individuals with active cancer (HR, 0.69; 95% CI, 0.50-0.95), or a primary immunodeficiency (HR, 0.74; 95% CI, 0.58-0.95). The rate of medically-attended COVID-19 was highest for individuals who had undergone blood/stem cell transplant (44.1 and 45.8 per 1,000 PYs for mRNA-1273 and BNT162b2, respectively) or solid organ transplant (56.8 and 58.9 per 1,000 PYs for mRNA-1273 and BNT162b2, respectively), with no difference observed between mRNA vaccines administered.

**Table 2.**
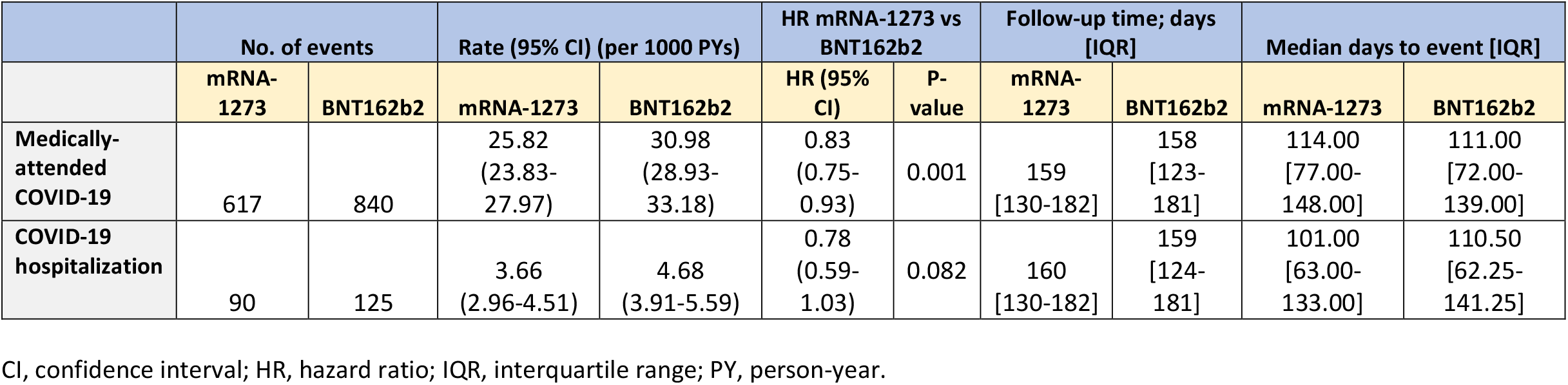
Incidence and VE of mRNA-1273 and BNT162b2 on medically-attended breakthrough COVID-19 diagnosis (primary outcome) and against breakthrough COVID-19 hospitalizations (secondary) among immunocompromised individuals.

**Figure 3.**
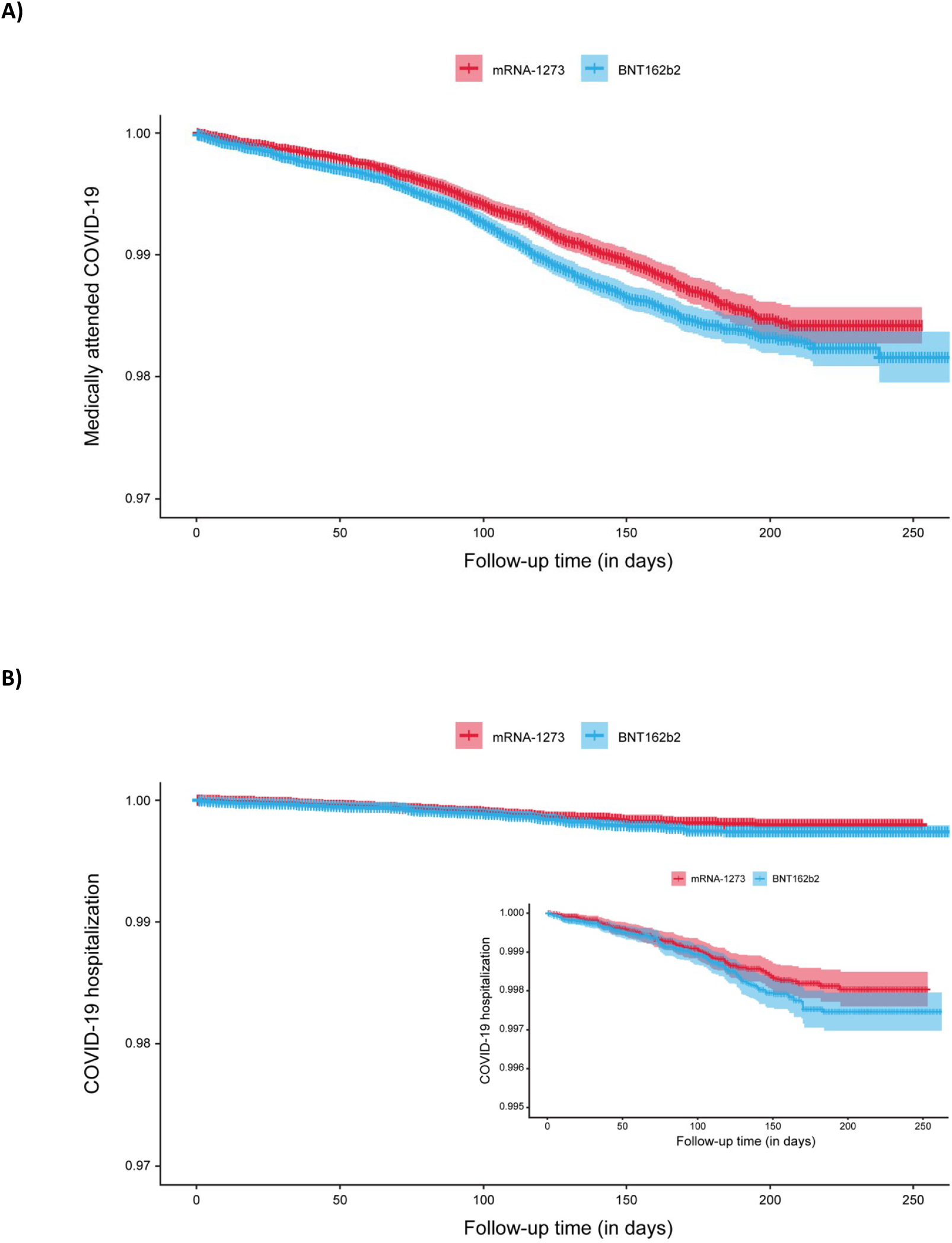
Comparative VE over time. Kaplan–Meier plots with 95% confidence intervals and Schoenfeld residuals over time. A) Medically-attended COVID-19; B) COVID-19 hospitalization.

The rate of breakthrough COVID-19 hospitalization was 3.66 per 1,000 PYs (95% CI, 2.96-4.51) among individuals who received 2 doses of mRNA-1273 and 4.68 per 1,000 PYs (95% CI, 3.91-5.59) among those who received 2 doses of BNT162b2, with a corresponding HR of 0.78 (95% CI, 0.59-1.03) (**Table 2; Figure 3B**). The number of hospitalizations resulted in low precision around the effect estimate and a lack of statistical power, limiting the interpretation of subgroup analyses (**Table S9**).

Sensitivity analyses for medically-attended COVID-19 demonstrated consistent results when not applying data truncation (HR, 0.83; 95% CI, 0.75-0.93) and when utilizing a 1:1 PS matching approach (HR, 0.84; 95% CI, 0.75-0.94) (**Figure 4; Tables S10**). When a COVID-19 test was required prior to a diagnosis code for COVID-19, the HR was 0.78 (95% CI, 0.67-0.92)(**Figure 4; Table S10**). By requiring a continuous closed claim enrollment file for 365 days before the index date and capturing events from closed claims only, the primary analysis was restricted both in the number of individuals included in the analytic cohort and in the number of events captured. Utilizing both open and closed claims during the follow-up period while enforcing the same approaches for cohort inclusion and follow up as the primary analysis, the number of medically-attended COVID-19 events increased from 1457 to 1734 with a resulting HR of 0.82 (95% CI, 0.74-0.90) (**Figure 4; Table S10)**. Utilizing open claims for both cohort entry and outcomes increased the cohort size from 124,457 to 388,965 immunocompromised individuals and increased the number of outcomes to 5,060 (HR, 0.74; 95% CI, 0.70-0.79) (**Figure 4; Table S10**).

**Figure 4.**
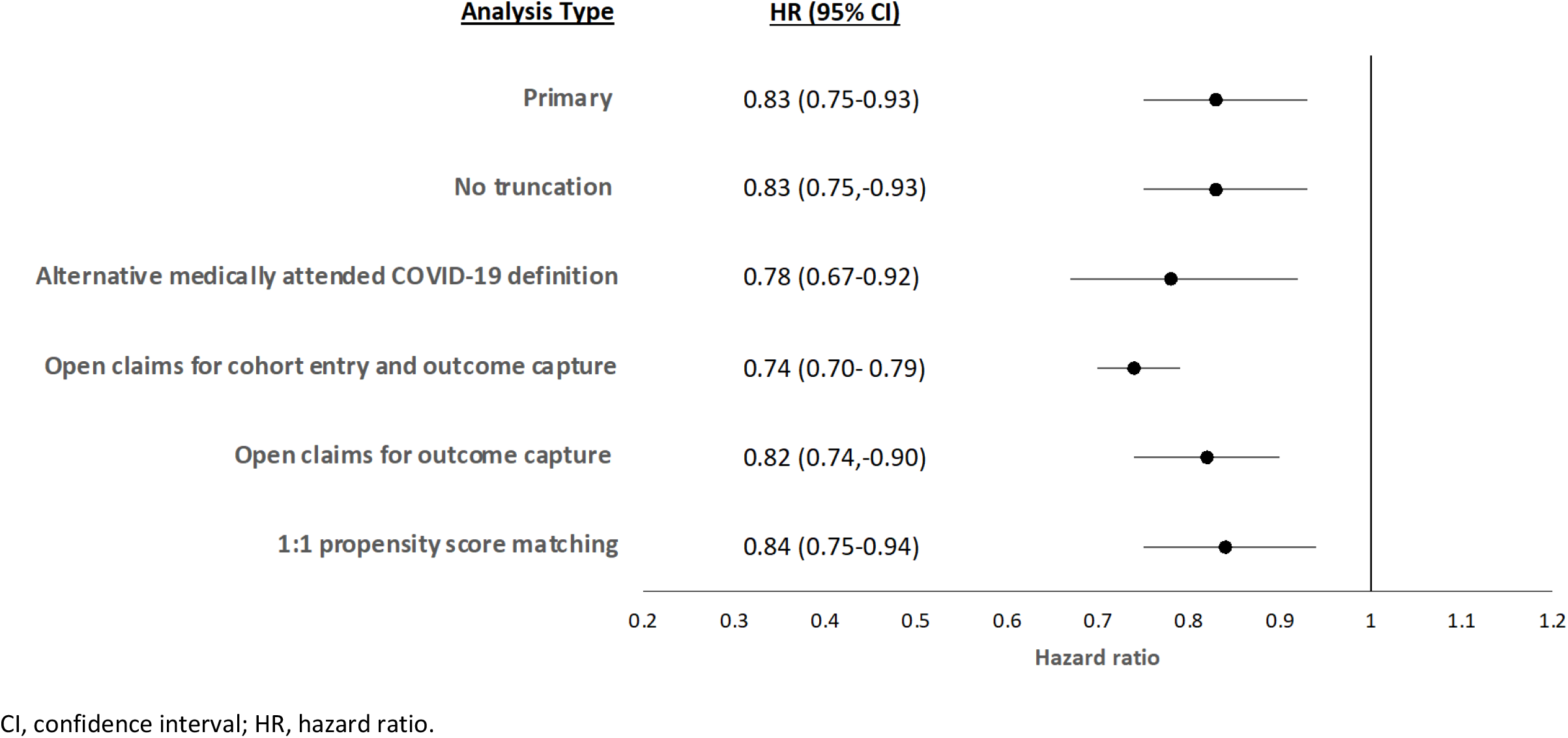
Sensitivity analyses for medically-attended COVID-19. Forest plot of hazard ratios of mRNA-1273 versus BNT-1626b in primary and sensitivity analyses of medically-attended COVID-19 data.

For the outcome of COVID-19 hospitalization, sensitivity analyses were consistent with the primary analysis (**Figure 5; Table S11**). When incorporating open claims for outcomes and utilizing the same approaches for cohort inclusion and follow-up as the primary analysis, the number of COVID-19 hospitalizations increased from 215 to 282, with a corresponding HR of 0.72 (95% CI, 0.57-0.92) (**Figure 5; Table S11**). When utilizing open claims for both cohort entry and outcomes, 874 COVID-19 hospitalizations were captured with a HR of 0.66 (95% CI, 0.58-0.76) (**Figure 5; Table S11**).

**Figure 5.**
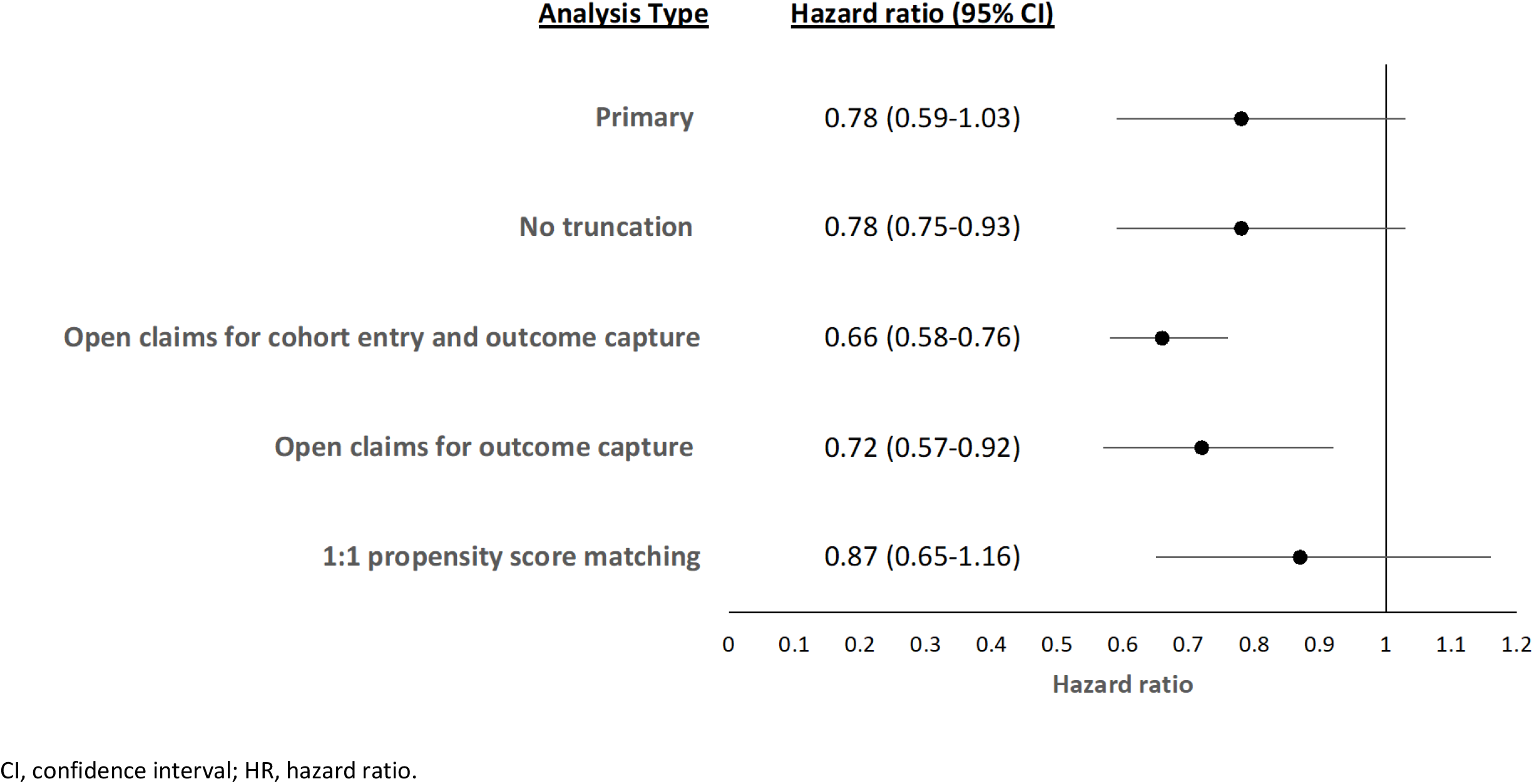
Sensitivity analyses for COVID-19 hospitalization. Forest plot of hazard ratios of mRNA-1273 versus BNT-1626b in primary and sensitivity analyses of COVID-19 hospitalization data.

## Discussion

Given the risk of severe COVID-19 outcomes in immunocompromised individuals, it is imperative to understand the real-world VE of COVID-19 vaccines; however, no studies have assessed the comparative VE of mRNA-based COVID-19 vaccines in this population. In the observational study reported herein, among a population of immunocompromised adults, 2 doses of mRNA-1273 were more effective in preventing breakthrough medically-attended COVID-19 infection compared to 2 doses of BNT162b2 (HR, 0.83; 95% CI, 0.75-0.93). The data also suggest that 2 doses of mRNA-1273 may be more effective compared to BNT162b2 in preventing breakthrough COVID-19 hospitalization (HR, 0.78; 95% CI, 0.59-1.03), although the small number of events resulted in low precision around the effect estimate. Our finding that the mRNA-1273 vaccine may be more effective in preventing medically-attended COVID-19 outcomes is consistent with real-world head-to-head studies in the general population and in adults with chronic comorbidities that were conducted during a time period when the delta and alpha variants dominated) [32-35]. Also at the time of the design of this study, while a third dose of mRNA-based vaccines gained authorization for IC individuals (recognized as a booster at the time), the sample size of the IC population who had received a third dose within claims records may not have been sufficiently large enough for to provide meaningful interpretation, thus limiting the current analysis to 2 dose regimens. Future work may consider the effectiveness of additional vaccine doses in this high-risk population.

Within immunocompromised subgroups, the comparative VE of a 2-dose regimen of mRNA-1273 compared with BNT162b2 against breakthrough medically-attended COVID-19 appeared to be greater among immunocompromised individuals with active cancer and among those with a primary immunodeficiency. Among individuals who had received blood/stem cell transplant, solid organ transplant, or those on active immunosuppressive therapy, the HRs were attenuated. Similar patterns were observed for COVID-19 hospitalization, although the small number of events and resulting lack of precision warrant caution in their interpretation.

Results of sensitivity analyses (incorporating open and closed claims, PS matching, foregoing application of the 3-month data truncation, and an alternative definition of medically-attended COVID-19 requiring evidence of a recent polymerase chain reaction or antigen test) were consistent with the primary analyses.

A strength of this head-to-head study is the utilization of a large claims database, which provided a sufficient sample size to detect statistically significant differences between vaccine groups of less common outcomes (ie, breakthrough COVID-19 diagnoses) among specific subpopulations such as immunocompromised individuals. Furthermore, incorporation of open and closed medical and pharmacy claims in sensitivity analyses provides insight into the implications of open claims. This resulted in real-time capture of outcomes and an almost three-fold increase in cohort size. For medically-attended breakthrough COVID-19 diagnoses (ie non-hospitalized COVID-19), the gains in speed and precision paired with consistent effect estimates with the primary analysis (utilizing closed claims only) provides support for future incorporation of open claims for outcome ascertainment in comparative VE studies, especially when paired with a required enrollment file among eligible individuals.

The results of this study should be interpreted in light of several limitations. First, there remains the potential for residual confounding by unmeasured variables. Second, identifying the primary outcome of medically-attended COVID-19 identified via the ICD-10 U-code for COVID-19 diagnosis in the inpatient or outpatient setting has not been previously validated specifically within the HealthVerity database. However, a recent study in the US Veterans Affairs database estimated the positive predictive value (PPV) of the ICD-10 code U07.1 in all settings (inpatient, outpatient, and emergency department/urgent care) to be 84.2% [36], which supports the robustness of this outcome and also cautiously noting the high PPV dependency associated with high disease prevalence within a pandemic setting. Furthermore, the analysis of an alternative definition of medically-attended COVID-19 requiring the presence of an antigen or polymerase chain reaction test produced an effect estimate slightly further from the null, thereby confirming the hypothesis that misclassification of the primary outcome would be non-differential between vaccine groups, with the resulting bias to be towards the null [37]. The difference in the HR between the base-case U code definition and the alternative definition requiring testing (0.05) is in line with our bias analysis based on the PPV estimate (see **supplementary text** and **Figure S1**). Additionally, in a real-world study of Ad26.COV2.S using the HealthVerity data, there was little difference in the VE when limiting the outcome to the COVID-19 diagnostic U-code in any setting or requiring a nucleic acid amplification lab confirmation of COVID-19 [28]. Regarding hospitalization for COVID-19, a similar algorithm as utilized in the current study has been previously leveraged in two published/ongoing cohort studies utilizing the HealthVerity claims database[28]. Recent validation studies of the ICD-9 U-code for COVID-19 in an inpatient setting also demonstrated a positive predictive value of at least 90%, again noting that PPV is dependent on disease prevalence)[38, 39].

A limitation of the HealthVerity claims data is the high amount of missingness of the place of service field (61.9% in both vaccinated groups across PS 17 and PS 20), which may also lead to misclassification of the outcomes. To mitigate this limitation, we identified suspected inpatient stays through a combination of claim types, billing codes, and place of service (where available) likely indicating an inpatient event. This study is also limited to individuals in the United States with continuous medical and pharmacy insurance coverage and does not include the uninsured, who may have addition co-morbidities or exposure factors placing them at higher risk of breakthrough COVID-19 infection and hospitalization. Also as demonstrated by the subgroup analyses, Medicare coverage is limited in the HealthVerity database and in other US commercial claims databases limiting inferences about, comparative VE in older individuals aged ≥65 years. Finally, due to the absence of variant specific laboratory data in this study, the potential effect modification by the circulating COVID-19 variants cannot be assessed, also noting the truncated data cut-off date (December 2021 prior to the dominant circulation of the omicron variant). The majority of patients included in the analytic cohort received a second dose of vaccine between March and May 2021, when the alpha variant was predominant in the US, with 15-20% receiving a second dose during June-September 2021, when the delta variant became prominent.[40] Despite this limitation, the outcome model included a state-based level of transmission, which incorporated calendar month of the start of follow-up for each patient, and the PS model included month and year of index date, both of which may, in part, control for the dominant circulating variant over the study period. Furthermore, subgroup analyses by calendar quarter provide insight into the impact of the circulating variants at that time.

Importantly, even with a large database such as HealthVerity, identifying a rare event such as breakthrough COVID-19 hospitalizations utilizing only closed claims may have limited the precision of effect estimates. Although use of fully adjudicated closed claims is typically the gold standard for database studies, open claims provide an ability to estimate the VE of vaccines and therapies with less lag (typically days to weeks for open claims versus 3-6 months for closed claims), which is instrumental in the setting of a global pandemic such as COVID-19 with rapid mass vaccination uptake. The consistency of the results from this study and the gain in precision suggest the ability to incorporate real-time open claims into comparative VE analyses. However, using open claims without a corresponding enrollment file may introduce bias, as baseline covariates could be under-captured, resulting in slightly different populations compared to a closed claims analysis. Therefore, our recommendation for future analyses is to use open claims are used in the follow-up phase for outcome ascertainment only.

In conclusion, results from this observational comparative VE database study provide evidence that among immunocompromised adults, 2 doses of mRNA-1273 is more effective in preventing breakthrough medically-attended COVID-19 infection compared to 2 doses of BNT162b2. Results also suggest that mRNA-1273 may have greater VE in preventing serious outcomes in comparison to BNT162b2 in this high-risk patient population.

## Data Availability

Individual-level data reported in this study are not publicly shared. Upon request, and subject to review, Aetion, Inc. may provide the de-identified aggregate-level data that support the findings of this study. De-identified data (including participant data as applicable) may be shared upon approval of an analysis proposal and a signed data access agreement.
Individual-level data reported in this study are not shared publicly but they are shared fully with regulatory agencies.

## Abbreviations

ACIP: Advisory Committee on Immunization Practice
ASD: absolute standardized difference
CDC: Centers for Disease Control and Prevention
COVID-19: coronavirus disease 2019
EUA: Emergency Use Authorization
FDA: Food and Drug administration
HIPAA: Health Insurance Portability and Accountability Act
IC: immunocompromised
PS: propensity score
RWE: real-world effectiveness
SARS-CoV-2: severe acute respiratory syndrome coronavirus 2
TTS: thrombocytopenia syndrome
VE: vaccine effectiveness

## Author contributions

KM, BK, SC, CT, DBT, DM, JM, XC, NMG, NVV conceptualized the study. KM, BK and NVV were responsible for data collection. KM, BK, DAP, AG, SC, CT, DBT, DM, JM, XC, NMG, NVV contributed to the analysis and interpretation of the data. KM, BK, DAP, AG, SC, CT, DBT, DM, JM, XC, NMG, NVV prepared, reviewed and edited the manuscript. All authors confirm that they had full access to all the data in the study and accept responsibility to submit for publication. All authors confirm that they had full access to all the data in the study and accept responsibility to submit for publication.

## Ethics committee approval statement

This study was exempt from approval by the New England Institutional Review Board.

## Acknowledgments

This study was funded by Moderna, Inc. Medical writing and editorial assistance were provided by Kate Russin, PhD, and Clare E. Lee, PhD, of MEDiSTRAVA in accordance with Good Publication Practice (GPP3) guidelines, funded by Moderna, Inc., and under the direction of the authors.

## Data sharing statement

Individual-level data reported in this study are not publicly shared. Upon request, and subject to review, Aetion, Inc. may provide the de-identified aggregate-level data that support the findings of this study. De-identified data (including participant data as applicable) may be shared upon approval of an analysis proposal and a signed data access agreement. Individual-level data reported in this study are shared fully with regulatory agencies.

