## Supplementary Material for "Real-world comparative effectiveness of mRNA-1273 and BNT162b2 vaccines among immunocompromised adults in the United States"

**Supplement**

***Supplementary text***

**Supplemental Methods**

**Study Population**

*Exclusion Criteria Definitions*

Prior COVID-19 infection was identified via the following diagnosis codes on an inpatient or outpatient claim: U07.1: “COVID-19, virus identified,” J12.82: “Pneumonia due to COVID-19,” or Z86.16: “Personal history of COVID-19.” Prior COVID-19 infection was also identified with diagnosis codes utilized early in the pandemic including J12.89: “Other viral pneumonia,” J20.8: “Acute bronchitis due to other specified organisms,” J40: “Bronchitis, not specified as acute or chronic,” J22:“Unspecified acute lower respiratory infection,” 98.8: “Other specified respiratory disorders,” or J80: “Acute respiratory distress syndrome.” The lookback period for these early pandemic codes was limited to March 1, 2020; the early pandemic codes applied before this date were not incorporated into the exclusion criteria.

**Statistical Analysis**

*Comparative Analysis*

The primary comparative analysis was implemented utilizing an exploratory, diagnostic, and inferential phase. Details of the exploratory analyses and resulting design decisions (truncation of claims data, time between vaccine doses, immunocompromised algorithm, and the claims-based definition of medically-attended COVID-19) are detailed in the study protocol. In the diagnostic phase, positivity was assessed for each value of the baseline covariates and baseline balance between vaccine groups was evaluated .

A propensity (PS) model was fit using logistic regression for mRNA-1273 versus BNT1262b2, modeling the probability of receiving the mRNA-1273 vaccine. Baseline covariates were included in the PS model included age (categorial), sex, primary payer type, state of residence, number of hospitalizations in the 365 days before the index date (continuous), number of outpatient interactions in the 365 days before the index date (continuous), frailty score (categorical), comorbidity score (categorical), immunocompromised subgroup (blood transplant, organ transplant, active cancer, primary immunodeficiency, HIV, use of immunosuppressive therapy), number of unique immunosuppressive therapies in the 365 days before the index date (continuous), and clinical comorbidities (see **Table S3** and **S4** for the full list).

The PS was applied utilizing 1:1 matching and weighting approaches. Pre- and post-adjustment diagnostics was assessed, including PS distribution within each vaccine group, PS overlap between vaccine groups, and absolute standard difference (ASDs) for variables included in the PS calculation, were assessed. To evaluate differential censoring, the distribution of follow-up time within each treatment group was described before assessment of the outcome of interest.

The inverse probability of treatment weight (IPTW) was calculated as 1/PS for the mRNA-1273 group (exposed) and 1/(1-PS) for the BNT162b2 group (referent), resulting in a pseudo-population in which the distribution of measured covariates used to calculate the PS becomes independent of the exposure (receipt of the mRNA-1273 vaccine). Following application of the weights, the balance in baseline covariates between the cohorts was assessed via ASDs. Exposure and referent groups were considered balanced if the ASD for all baseline covariates used to generate the propensity score was <0.10.[1-3] Additionally, the distributions of baseline covariates were assessed between the index date and 13-days following completion of the 2-dose vaccine regimen to ensure that the distributions did not change between the index date and the state of follow-up (**Tables S6 and S7**).

Level of COVID-19 transmission by state was determined based on county level transmission data reporting the number of new cases in the previous 7 days. The last available date of each month was used to aggregate county-level data and classify the transmission category at the state level. The cases per 100,000 persons were classified as follows: low = 0-9.99, moderate = 10-49.99, substantial = 50-99.99 and high ≥100. To ensure positivity within the covariate categories, low and moderate were grouped together in the analysis. In addition to the sensitivity analyses described in **Table S1**, 1: 1 PS matching analysis was conducted in closed medical claims data that was truncated 3 months (from December 11, 2020 to October 12, 2021).

*Bias Analysis*

False positives within large claim databases bias effect estimates towards the null. Therefore to evaluate the power of this study in detecting differences for breakthrough COVID-19, the size of the bias was estimated prior to running the analysis based on several assumptions. First, the positive predictive value (PPV) of the ICD-10 COVID-19 U07.1 code was assumed to be the same in the HealthVerity (HV) database as observed by Lynch et al (2021) in the US Veterans Affairs database (VA; equation 1: PPV (VA/HV) = 0.842).[4] Second, an equal number of IC individuals were assumed to be vaccinated with mRNA-1273 or BNT162b2 (equation 2: #IC (Moderna) = #IC (Pfizer) = 60,000). Third, based on other claim studies the true hazard ratio (HR) was estimated to be approximately 0.7 (mRNA-1273/BNT162b2). Fourth, using the notation described in **Figure S1**, the observed HR was calculated using equation 3 (Observed HR= (aM+bM)/(aP+bP)). Fifth, assuming the sensitivity and specificity of COVID tests were 0.9 and 0.98, respectively, the prevalence in VA and HV were calculated using equation 4 (PPV = (Sensitivity x Prevalence) / ((Sensitivity x Prevalence) + ((1-Specificity) x (1 - Prevalence))) and equation 1 (PPV(VA/HV) = 0.842).

Accordingly, based on this prevalence of 0.106 and true HR, the prevalence of adults vaccinated with mRNA-1273 or BNT162b2 was estimated to be 0.087 and 0.125, respectively. Using equation 4 PPV = (Sensitivity x Prevalence) / ((Sensitivity x Prevalence) + ((1-Specificity) x (1 - Prevalence)))) again, the PPV for mRNA-1273 and BNT162b2 were 0.811 and 0.865, respectively. Inputting this into the full 2x2 tables (**Figure S1**) resulted in the following values: aM = 5232, aP = 7474, bM = 1217, bP = 1167. Then inputting values using equation 3 (Observed HR= (aM+bM)/(aP+bP) afforded an observed HR of 0.75 with the expected bias towards the null calculated as 0.05, which align with the difference in results between the sensitivity analysis requiring a COVID-19 test to inform the U code and the base-case analysis.

**Supplementary Results**

The distributions of PS before and after matching are displayed in **Figure S2a** and **Figure S2b**, respectively. While both matching (**Table S5**) and weighting approaches achieved baseline balance between vaccine groups (with no ASDs >0.10 for baseline covariates included in the PS model), matching resulted in 52,729 matched patients in each vaccine group. Weighting maintains the full study population. Furthermore, the distribution of IPT weights was similar between each vaccine group, with no extreme values (range: 1.16-6.91) (**Table S6**). Given the ability to achieve baseline balance in the resulting weighted pseudopopulation and to attain greater precision than matching, weighting was chosen as the primary analytic approach. Given the narrow range of IPT weights, no truncation to the weights was applied in the primary analysis.

*Supplementary Tables*

**Table S1. Summary of analyses, outcomes and evaluated cohorts**

| Analysis type | Outcome | Cohorts |
|---|---|---|
| **Primary analyses** | | |
| **Primary outcome** | Medically-attended COVID-19 diagnosis (ICD-10 diagnostic code for COVID-19 in any setting) | Closed medical claims only and data were truncated 3 months (to October 12, 2021) |
| **Secondary outcome** | Hospitalization for COVID-19 (hospital stay with the ICD-10 diagnostic code for COVID-19 as primary diagnosis code or 21 days before hospital admission) | |
| **Sensitivity analyses** | | |
| **No truncation** | Same as primary analyses | Closed medical claims only. No data truncation/included date through January 10, 2021 |
| **Alternative medically-attended COVID-19 definition** | Medically-attended COVID-19 diagnosis requiring evidence of an antigen (rapid) test on the same day or 3 days before the COVID-19 diagnosis code or a PCR test 3-14 days before the COVID-19 diagnosis | Same as primary analyses |
| **Open claims for cohort entry and outcome capture** | In addition to closed claims, open claims with a diagnosis code for COVID-19 were incorporated into the outcomes of interest | Individuals without an enrollment file, with open claims only, were required to have at least one open medical and open pharmacy claim in the 365 days before the index date. During follow-up, individuals without an enrollment file or those who disenroll from their medical/pharmacy plan continued follow-up if there was evidence of a medical or pharmacy claim every 60 days. |
| **Open claims for outcome capture** | Same as above | Same as above, except individuals were required to have an enrollment file on the index date with continuous enrollment in the 365 days before and during follow-up |

PCR, polymerase chain reaction.

**Table S2. Codes used to identify the exposure and referent groups**

| Variable name | Definition |
|---|---|
| **Exposure group[a]** | |
| **mRNA-1273** | • Individuals were required to have evidence of 2 doses of the mRNA-1273 vaccine, and were identified via the following CPT and/or NDC codes:<br>• CPT codes: 91301-0011A, 91301-0012A, 0011A, 0012A, 91301<br>• NDC codes: 80777027310, 80777027315, 80777027398, 80777027399, 8077727315, 8077727398, 8077727399, 8077727310, 80777-0273-10, 80777-273-10, 80777-0273-15, 80777-0273-98, 0777-273-15, 80777-273-99, 80777-0273-99<br>• The second vaccine NDC/CPT code was at least 14 days after the first dose to be considered a second dose. |
| **Referent group[b]** | |
| **BNT1262b2** | • Fully vaccinated BNT1262b2 individuals were required to have evidence of 2 doses of the BNT1263b2 vaccine, and were identified via the following CPT and/or NDC codes:<br>• CPT codes: 91300-0001A, 91300-0002A, 0001A, 0002A, 91300<br>• NDC codes: 59267100001, 59267100002, 59267100003, 5926710001, 5926710002, 5926710003, 59267-1000-01, 59267-1000-1, 59267-1000-02, 59267-1000-03, 59267-1000-2<br>• The second vaccine NDC/ and CPT code was at least 14 days after the first dose to be considered a second dose. |

[a]Assigned a weight of 1/ PS.
[b]Assigned a weight of 1 / (1 - PS).
CPT, current procedural terminology; NDC, national drug code; PS, propensity score

**Table S3. Claims-based algorithm to identify immunocompromised individuals**

| Immunocompromised condition | Algorithm description |
|---|---|
| **Blood transplant** | Blood transplant 2 years before CED. Defined as at least one of the following ICD-10 diagnosis codes: D84.9, T86.00, T86.09, Z48.290, D81.1, D84.81, Z94.81<br><br>OR<br><br>At least one of the following ICD-10 procedure codes:<br>30230AZ, 30233AZ, 30240AZ, 30253Y1, 30260G1, 30260Y1, 30263G1, 30243AZ, 30250G1, 30250X1, 30250Y1, 30253G1, 30253X1, 30260X1, 30263X1, 30263Y1, 30230G2, 30230G3, 30230G4, 30230X1, 30230X3, 30230X4, 30230Y1, 30230Y2,<br>30230Y3, 30230Y4, 30233G1, 30233G4, 30233X1, 30233X2, 30233X3, 30233X4, 30233Y1, 30240G1, 30240G2, 30240G3, 30240G4, 30240X2, 30240X4, 30240Y2, 30240Y4, 30243G1, 30243G2, 30243G3, 30243X1, 30243X4, 30243Y2, 30243Y3, 30243Y4, 30230G1, 30230X2, 30233G2, 30233G3, 30233Y2, 30233Y3, 30233Y4, 30240X1, 30240X3, 30240Y1, 30240Y3, 30243G4, 30243X2, 30243X3, 30243Y1 |
| **Stem cell transplant** | Stem cell transplant 2 years prior to CED. Defined as evidence of at least one of the following ICD-10 procedure codes: 30230AZ, 30230G1, 30230G3, 30230G4, 30230X1, 30230X2, 30230Y1, 30230Y3, 30233G3, 30233G4, 30233X2, 30233X4, 30233Y1, 30233Y2, 30233Y4, 30240AZ, 30240G1, 30240G2, 30240G3, 30240G4, 30240X1, 30240X3, 30243G2, 30243G4, 30243X1, 30243X2, 30243X4, 30243Y1, 30243Y2, 30243Y4, 30250G1, 30253G1, 30253X1, 30253Y1, 30260G1, 30260Y1, 30263G1, 30263X1, 30230G2, 30230X3, 30230X4, 30230Y2, 30230Y4, 30233AZ, 30233G1, 30233G2, 30233X1, 30233X3, 30233Y3, 30240X2, 30240X4, 30240Y0, 30240Y1, 30240Y2, 30240Y3, 30240Y4, 30243AZ, 30243G1, 30243G3, 30243X3, 30243Y3, 30250X1, 30250Y1, 30260X1, 30263Y1<br><br>OR<br><br>Any of the following HCPCS codes: 38240, 38242, 38243, S2150 |
| **Organ transplant with immunosuppressive therapy** | Any history of an organ transplant and evidence of immunosuppressive therapy (list of immunosuppressive therapies below) in the 60 days before the CED.<br><br>Organ transplant was defined having as any medical claim with at least one of the following ICD-10 diagnosis codes:<br>D84.821, D84.9, T86.19, T86.20, T86.298, T86.39, T86.49, T86.818, T86.859, T86.898, T86.899, Z48.21, Z48.280, Z48.288, Z94.82, Z94.83, T86.10, T86.30, T86.40, T86.819, T86.858, Z48.22, Z48.23, Z48.24, Z94.0, Z94.1, Z94.2, Z94.3, Z94.4 |

| | |
|---|---|
| | OR<br><br>At least one of the following ICD-10 procedure codes: 02YA0Z0, 02YA0Z2, 07YM0Z1, 07YM0Z2, 07YP0Z1, 0BYC0Z1, 0BYC0Z2, 0BYD0Z2, 0BYF0Z0, 0BYG0Z0, 0BYG0Z1, 0BYH0Z0, 0BYH0Z2, 0BYJ0Z1, 0BYJ0Z2, 0BYK0Z1, 0BYK0Z2, 0BYL0Z0, 0BYL0Z1, 0BYL0Z2, 0BYM0Z0, 0BYM0Z2, 0DY60Z0, 0DY60Z1, 0DY60Z2, 0DY80Z1, 0DY80Z2, 0DYE0Z0, 0DYE0Z1, 0DYE0Z2, 0FY00Z1, 0TY10Z0, 0TY10Z1, 3E1M39Z, 5A1D70Z, 5A1D90Z, BT2900Z, BT290ZZ, BT2910Z, BT29Y0Z, BT29ZZZ, BT39ZZZ, 02YA0Z1, 07YM0Z0, 07YP0Z0, 07YP0Z2, 0BYC0Z0, 0BYD0Z0, 0BYD0Z1, 0BYF0Z1, 0BYF0Z2, 0BYG0Z2, 0BYH0Z1, 0BYJ0Z0, 0BYK0Z0, 0BYM0Z1, 0DY50Z0, 0DY50Z1, 0DY50Z2, 0DY80Z0, 0FY00Z0, 0FY00Z2, 0FYG0Z0, 0FYG0Z1, 0FYG0Z2, 0TY00Z0, 0TY00Z1, 0TY00Z2, 0TY10Z2, 5A1D00Z, 5A1D80Z, BT291ZZ, BT29YZZ, BT39Y0Z, BT39YZZ, BT49ZZZ<br><br>OR<br><br>At least one of the following HCPCS codes: 47135, 47136, 50365, A4673, A4674, A4680, A4700, A4705, A4708, A4712, A4714, A4719, A4720, A4723, A4724, A4730, A4736, A4740, A4750, A4765, A4850, A4870, A4900, A4901, A4905, A4912, A4913, A4914, A4918, E1500, E1530, E1575, E1590, E1592, E1594, E1610, E1615, E1620, E1632, E1635, S2053, S2054, S2060, S2065, S2142, 32851, 32852, 32853, 32854, 33935, 33945, 44135, 44136, 48554, 50360, 50370, A4653, A4671, A4672, A4690, A4706, A4707, A4709, A4721, A4722, A4725, A4726, A4728, A4735, A4737, A4755, A4760, A4766, A4802, A4820, A4860, A4880, A4890, A4910, A4911, E1510, E1520, E1540, E1550, E1560, E1570, E1580, E1600, E1625, E1630, E1634, E1636, S2152<br><br>OR<br><br>At least one of the following DRG codes: 103, 302, 480, 495 |
| **Active Cancer: cancer diagnosis in the year before cancer therapy and cancer therapy within 180 days prior to index** | Active cancer therapy (180 days prior to CED) AND cancer Dx (365 days before therapy)<br><br>Cancer malignancy was defined as having any medical claim with at least one of the following ICD-10 diagnosis codes: C00, C01, C02, C03, C04, C05, C06, C07, C08, C09, C10, C11, C12, C13, C14, C15, C16, C17, C18, C19, C20, C21, C22, C23, C24, C25, C26, C30, C31, C32, C33, C34, C37, C38, C39, C40, C41, C43, C45, C46, C47, C48, C49, C50, C51, C52, C53, C54, C55, C56, C57, C58, C60, C61, C62, C63, C64, C65, C66, C67, C68, C69, C70, C71, C72, C73, C74, C75, C76, C81, C82, C83, C84, C85, C88, C90, C91, C92, C93, C94, C95, C96, C97<br><br>Use of cancer therapy was defined as having any medical claim or pharmacy claim with at least one of the following NDC codes (https://seer.cancer.gov/oncologytoolbox/): |

| | |
|---|---|
| | 2298060, 2416502, 2416507, 2416534, 2418402, 2418407, 2418430, 2481554, 2533754, 2750201, 2762301, 2766901, 3029305, 3029320, 3031505, 3049420, 3052711, 3085222, 3085522, 3085722, 3229111, 3232822, 3373413, 3377211, 3452211, 4015549, 4024126, 4024133, 4024208, 4035009, 4110150, 4110175, 6046101, 6046102, 6046130, 6046201, 6046405, 6302602, 6302604, 6306100, 6306104, 6306601, 6306603, 6394101, 6404500, 6404501, 6404541, 6411902, 6503302, 6503402, 7326036, 7326201, 7420101, 7420511, 7420711, 7440106, 8010001, 9000302, 9001104, 9001305, 9001612, 9002001, 9002201, 9003928, 9003932, 9003933, 9004722, 9004902, 9005605, 9006404, 9006406, 9007301, 9011312, 9017601, 9028003, 9028024, 9028025, 9028051, 9028052, 9030612, 9062601, 9069801, 9075801, 9079601, 9090013, 9090020, 9090908, 9092003, 9111101, 9307322, 9307323, 9338901, 9347522, 9509101, 9509301, 9752903, 9752905, 13126683, 13257691, 13258691, 13871762, 13872789, 15050241, 15050401, 15050541, 15050641, 15059501, 15059641, 15303020, 15303120, 15307220, 15308060, 15321030, 15321230, 15340420, 15355427, 15355626, 15356415, 19945201, 19945275, 23590623, 24059010, 24059120, 24061030, 24065401, 24586001, 24586201, 24591701, 46110081, 46110251, 46110281, 46110291, 46110381, 51002221, 51002321, 54001725, 54001729, 54001825, 54001829, 54001920, 54001925, 54008013, 54014308, 54014387, 54016413, 54024822, 54024913, 54026913, 54027121, 54027223, 54032003, 54032103, 54032303, 54032306, 54032403, 54032406, 54038225, 54039925, 54048013, 54048113, 54317644, 54317763, 54372144, 54372250, 54372263, 54412925, 54418025, 54418125, 54418225, 54418325, 54418425, 54449613, 54449625, 54449810, 54449911, 54460325, 54472831, 54817525, 54849619, 54860325, 54860425, 54872216, 54872425, 54874025, 54981725, 54982825, 69007501, 69007801, 69007901, 69008101, 69008407, 69008618, 69009901, 69010303, 69010701, 69013501, 69014602, 69014701, 69014801, 69014901, 69015201, 69015302, 69015401, 69017101, 69017401, 69017601, 69017701, 69017702, 69017801, 69017901, 69018101, 69018601, 69018721, 69018821, 69019202, 69020401, 69020410, 69020510, 69020550, 69022701, 69023101, 69023801, 69029101, 69029110, 69029210, 69029860, 69030501, 69030801, 69031501, 69055038, 69068803, 69070012, 69101001, 69119830, 69130710, 69130810, 69130904, 69131110, 69134002, 69144140, 69153130, 69229930, 69303020, 69303320, 69385810, 69385910, 69400405, 69401510, 69403001, 69403201, 69403701, 69449622, 69454102, 69454302, 69454501, 69454502, 69454701, 69454702, 69814020, 69814120, 74054130, 74056114, 74056611, 74310932, 74334603, 74347303, 74364203, 74368303, 74647932, 74726950, 75800180, 75800301, 75800404, 78001715, 78010215, 78010901, 78018001, 78018061, 78018201, 78018261, 78018425, 78024015, 78024061, 78024161, 78024615, 78024815, 78034261, 78043815, 78049561, 78052651, 78056651, 78056751, 78059061, 78059287, 78059461, 78062651, 78062661, 78062751, 78062761, 78062851, 78062861, 78064515, |

| | |
|---|---|
| | 78064681, 78064881, 78065006, 78065106, 78066615, 78066815, 78066913, 78067201, 78067461, 78067919, 78068166, 78068306, 78068361, 78069061, 78069484, 78070184, 78070956, 78071656, 78081181, 78082581, 78084619, 78092361, 78095819, 85113301, 85123501, 85124201, 85128702, 85136601, 85136602, 85136603, 85136605, 85138801, 85138802, 85141701, 85142501, 85142503, 85142505, 85151902, 85151903, 85300401, 85300402, 85300404, 85300405, 85434801, 85434901, 85435101, 88111114, 88120205, 88120305, 88120632, 88120806, 88120876, 89061012, 93022056, 93023319, 93023333, 93023356, 93023393, 93078201, 93078405, 93078410, 93565556, 93574019, 93574065, 93574119, 93574165, 93574219, 93574265, 93611816, 93611887, 93612619, 93612664, 93723656, 93723693, 93729001, 93729010, 93729056, 93730119, 93730203, 93735505, 93735556, 93735598, 93747489, 93748512, 93748519, 93759941, 93760041, 93760257, 93763056, 93763941, 93766456, 93776624, 93776824, 93901865, 93901965, 93902065, 95008735, 95008851, 115140801, 115143808, 115147623, 115169606, 115169701, 115170001, 121077708, 121090204, 143120201, 143125401, 143125425, 143142501, 143147301, 143147510, 143147525, 143147701, 143147705, 143147710, 143147725, 143242407, 143921701, 143921901, 143924001, 143924101, 143927001, 143927501, 143929001, 143929101, 143930801, 143954601, 143954701, 143954810, 143955110, 143955301, 143960601, 143964201, 143973801, 143973810, 143973901, 143973910, 143974001, 143974505, 143977106, 143989001, 143989105, 143989125, 172496058, 172524160, 172731000, 172731146, 172731200, 173013093, 173044604, 173044702, 173044704, 173048900, 173056900, 173057004, 173071204, 173071225, 173071325, 173075200, 173080409, 173080802, 173080805, 173082101, 173082133, 173084813, 173084865, 178058201, 179009944, 179010044, 179012470, 182186389, 182186400, 185015501, 185015601, 185093230, 185093386, 185740014, 185740085, 187090101, 187552560, 187552675, 310020137, 310048230, 310061060, 310067912, 310067960, 310070510, 310070530, 310095036, 310134930, 310135030, 310450012, 310470001, 310471511, 310781030, 310782030, 338000801, 338006301, 338130501, 338399101, 338399301, 378027493, 378031553, 378034453, 378034493, 378064001, 378064101, 378064210, 378100394, 378145201, 378145405, 378145801, 378204293, 378207105, 378251278, 378309685, 378526014, 378526098, 378526114, 378526198, 378526298, 378526398, 378526498, 378603405, 378661185, 378661293, 378661485, 378661493, 378686801, 378695501, 378709693, 378713293, 378817091, 378817191, 378817291, 378904001, 378904005, 378904501, 409018101, 409018301, 409018325, 409018601, 409020125, 409020126, 409020127, 409030201, 409036601, 409080101, 409080109, 409112011, 409112012, 409112062, 409321705, 409341401, 409421505, 409422901, 409475501, 409475502, 409475512, 409475518, 409568401, 409568423, 409568502, 409568523, 440816710, 469062599, 469072560, 469142590, 517042001, 517490125, 517493025, 527145206, 527172640, 527293037, 527293043, 527293237, 527293337, 527293437, 527293441, 527293537, 555044663, 555048402, 555048527, 555057202, 555060602, 555060703, 555060704, |

| | |
|---|---|
| | 555087202, 555087204, 555087302, 555088602, 555088704, 555089902, 555090401, 555090414, 555105456, 555105486, 555105556, 555105586, 555105686, 555105786, 574014804, 574082105, 591048801, 591222215, 591223218, 591223260, 591223319, 591229219, 591229230, 591236701, 591236730, 591243315, 591243515, 591243615, 591245115, 591246618, 591247218, 591247330, 591250115, 591322126, 591356279, 591356355, 591359260, 591359360, 591438579, 591501902, 591505201, 591544243, 591544305, 597014130, 597014360, 597014560, 603363302, 603363316, 603363321, 603363328, 603390121, 603459315, 603459321, 603533510, 603533532, 603533621, 603533715, 603533815, 603533821, 603533828, 603533921, 603533928, 641036721, 641036725, 641607801, 641607825, 641614501, 641614525, 641617401, 641617601, 641617610, 641617701, 641617801, 703003101, 703003104, 703004301, 703005101, 703005104, 703024101, 703024501, 703301513, 703312508, 703315401, 703315501, 703321301, 703321601, 703321681, 703321701, 703321881, 703330101, 703330104, 703331101, 703332194, 703333301, 703334301, 703342711, 703342911, 703352403, 703367103, 703367501, 703367591, 703367881, 703398501, 703401411, 703401418, 703408551, 703410048, 703410058, 703410068, 703415411, 703415491, 703415611, 703415691, 703418291, 703418301, 703424401, 703424481, 703424681, 703443211, 703443481, 703450284, 703463601, 703468001, 703468501, 703471401, 703471471, 703476481, 703476601, 703476681, 703476701, 703480501, 703485211, 703485291, 703507501, 703507503, 703514501, 703523311, 703523393, 703565301, 703565691, 703572001, 703574811, 703577501, 703585401, 703722102, 703722104, 703722601, 703722603, 703723939, 703789101, 703797301, 781167931, 781168131, 781211901, 781211931, 781232106, 781232268, 781269344, 781269444, 781269475, 781302975, 781307912, 781313171, 781313271, 781313295, 781313670, 781316475, 781320194, 781324375, 781324494, 781324570, 781325394, 781325594, 781328379, 781329680, 781331570, 781341575, 781502201, 781532501, 781532531, 781535631, 781540901, 781540964, 781715209, 781916475, 904601260, 904622961, 904655261, 904674561, 904674604, 904691461, 944262003, 944265503, 944265804, 944381001, 955102001, 955172510, 955173110, 10019090501, 10019090604, 10019092501, 10019092602, 10019093501, 10019093701, 10019093801, 10019094301, 10019094501, 10019095302, 10019095362, 10019095501, 10019098201, 10019098401, 10139006202, 10139006210, 10139006240, 10139006301, 10139006310, 10139006311, 10139006320, 10544027421, 10544047320, 10544053830, 10544091420, 10631000231, 10631000331, 10631013331, 10631013431, 10768708501, 10768773303, 10885000101, 11399000501, 11994001601, 13668045301, 13668059181, 13668059182, 13668059286, 13668059386, 13925050104, 14789060010, 15014021121, 15054006001, 15054009001, 15054106003, 15054106004, 15054109003, 15054112003, 15054112004, 16477050501, 16477050548, 16477051008, 16571042103, 16590014921, 16590032610, 16590032621, 16590032630, 16590036521, 16590037321, 16590062421, 16590062448, 16714002701, 16714011802, 16714013001, 16714013025, 16714013101, 16714013701, 16714014001, 16714015001, 16714022101, 16714022110, 16714022132, 16714046501, 16714046701, 16714047301, |

| | |
|---|---|
| | 16714057101, 16714072501, 16714072601, 16714081501, 16714083401, 16714085701, 16714085801, 16714089001, 16714091501, 16714092801, 16714092901, 16714093001, 16714096301, 16729002301, 16729003510, 16729004853, 16729004953, 16729005054, 16729007329, 16729009016, 16729011408, 16729011411, 16729011505, 16729011638, 16729011711, 16729012953, 16729012954, 16729013053, 16729022405, 16729023164, 16729023165, 16729024330, 16729024331, 16729026231, 16729026764, 16729026765, 16729027605, 16729027730, 16729027735, 16729029512, 16729029533, 16729029783, 16729030610, 16729033203, 16729035192, 16729036566, 16729041903, 16729043630, 17156052401, 17478032705, 17478054602, 17478054604, 17478054605, 17478076106, 17478076206, 17856069104, 17856069105, 17856075902, 21695008021, 21695011100, 21695030530, 21695030620, 21695030628, 21695030630, 21695030639, 21695030642, 21695030712, 21695030714, 21695030720, 21695030730, 21695036001, 21695036010, 21695036508, 21695036516, 21695038204, 21695038220, 21695038260, 21695058005, 21695058014, 21695061330, 21695062330, 21695076521, 21695082940, 21695083412, 21695083430, 21695084910, 21695085005, 21695089610, 21695089620, 21695095205, 21695099030, 23155017931, 23155017932, 23155019631, 23155024041, 23155037731, 23155037831, 23155037842, 23155048331, 23155048431, 23155052931, 23155054731, 23155054742, 23155054831, 23155054841, 23155054931, 23155068631, 23594050501, 23594050502, 23594050550, 23594051008, 23594091508, 24201010104, 24201023701, 24338005008, 25021020110, 25021020205, 25021020251, 25021020351, 25021020661, 25021020705, 25021020725, 25021021250, 25021021305, 25021021317, 25021021598, 25021021599, 25021022204, 25021022207, 25021023005, 25021023926, 25021023952, 25021024501, 25021024602, 25021045301, 25021045405, 25021046274, 25021077801, 25021077866, 25021078220, 25021080166, 25021080810, 25021081467, 25021081530, 25021081667, 25021082666, 25021082682, 29336061012, 30237890006, 30698001701, 30698010230, 30698012005, 30698012105, 30698020101, 30698020130, 30698020230, 31722013130, 31722052510, 31722096060, 33261012921, 33261035221, 33261035248, 33261041705, 33358051230, 35356021030, 35356024900, 35356042630, 35356044530, 35356065210, 35356065220, 35356067320, 35356067418, 35356067430, 35356067715, 35356067720, 35356067740, 35356067790, 35356067830, 35356067910, 35356067930, 35356075565, 35356076321, 35356081810, 35356081815, 35356081818, 35356081920, 35356081940, 36000001225, 36000001301, 38423011001, 39822210002, 39822212001, 39822220001, 41616048588, 42023011201, 42023013601, 42023014901, 42023015101, 42043018003, 42043039000, 42043039020, 42043039040, 42195012106, 42195015021, 42195015049, 42195015110, 42195072121, 42238011112, 42254007730, 42254009710, 42254010208, 42254011030, 42254016001, 42254021310, 42291008530, 42291010530, 42291016660, 42291016850, 42291019060, 42291028050, 42291035190, 42291045060, 42291050501, 42291072610, 42291072690, 42291076901, 42291077050, 42291077101, 42291077150, 42292005105, 42292005205, 42292005703, 42367052125, 42388001314, 42388002326, 42543014050, 42543014101, 42549052221, 42549065730, 42549065760, |

| | |
|---|---|
| | 42549065790, 42658001001, 42799081201, 42806008801, 42806008805, 42806035925, 42858086706, 42858086806, 42858086906, 43063005202, 43063020101, 43063020801, 43063020860, 43063042612, 43063042621, 43063042628, 43063042630, 43063042642, 43063042660, 43063043220, 43063043221, 43063043810, 43063043930, 43063044601, 43063044690, 43063047225, 43063056002, 43063056003, 43063056005, 43063056006, 43063056010, 43063059006, 43063059012, 43063059014, 43063059021, 43063059030, 43063061021, 43063064321, 43063064410, 43063064415, 43063064420, 43063064421, 43063064430, 43063064442, 43063064450, 43063065130, 43063070310, 43063070315, 43063070320, 43063070321, 43063070690, 43063071730, 43063071790, 43063077006, 43063079206, 43066001001, 43066001801, 43353025360, 43353068760, 43353068860, 43353081960, 43598026202, 43598033011, 43598034431, 43598034490, 43598034531, 43598034837, 43598038957, 43598042737, 43598046562, 43598050501, 43598050510, 43598054125, 43598061111, 43598065011, 43598067811, 43598077111, 43598085911, 43598086560, 43598094811, 43975025305, 43975025405, 43975025505, 43975030710, 43975031510, 44087353501, 44087400000, 44087400006, 44087400007, 44087400008, 44087500003, 44206045824, 44523018208, 44567050501, 44567050901, 44567051001, 45629008901, 45629013401, 45802030321, 45802030367, 45802036862, 45865059810, 45963050002, 45963050030, 45963053830, 45963053930, 45963060860, 45963060868, 45963060955, 45963061153, 45963061359, 45963061383, 45963061455, 45963061481, 45963061485, 45963061556, 45963062060, 45963062357, 45963062458, 45963063660, 45963063858, 45963064057, 45963068602, 45963073357, 45963073360, 45963073368, 45963073454, 45963076257, 45963076552, 46026098301, 47335004640, 47335004940, 47335005040, 47335008250, 47335015040, 47335015340, 47335015440, 47335017640, 47335048583, 47335048588, 47335071481, 47335071583, 47335089074, 47335089080, 47335089172, 47335089272, 47335089540, 47335092972, 47335092974, 47335093021, 47335093072, 47335093074, 47335093080, 47335093640, 47335096241, 47426010106, 47426020101, 47781010830, 47781020050, 47781057807, 47781059122, 47781059307, 47781060320, 47781060594, 47781060694, 47781062222, 48102004501, 48102004601, 48102004701, 48102004720, 48102004801, 48102005101, 49281088003, 49663000106, 49884008501, 49884008701, 49884028901, 49884029004, 49884029005, 49884032462, 49884037301, 49884072401, 49884075313, 49884086702, 49884086902, 49884092202, 49999000820, 49999000855, 49999002812, 49999002815, 49999002830, 49999002840, 49999008330, 49999010930, 49999011006, 49999011018, 49999011021, 49999011030, 49999015321, 49999015330, 50090008800, 50090008804, 50090008900, 50090009200, 50090009700, 50090010200, 50090016602, 50090025600, 50090027101, 50090043600, 50090048501, 50090049002, 50090065500, 50090065501, 50090081900, 50090095500, 50090101501, 50090112800, 50090145500, 50090152301, 50090160002, 50090160004, 50090165800, 50090165900, 50090166002, 50090170400, 50090171801, 50090171802, 50090198902, 50090198903, 50090198905, 50090198909, 50090199001, 50090199005, 50090231302, 50090236000, 50090240200, 50090249000, 50090260800, 50090260801, |

| | |
|---|---|
| | 50090263800, 50090278905, 50090278909, 50090346600, 50090478507, 50242006010, 50242006110, 50242006201, 50242006401, 50242007001, 50242009002, 50242009130, 50242009490, 50242010301, 50242010501, 50242013086, 50242024501, 50242026001, 50242071701, 50242091701, 50242091786, 50242091801, 50268029111, 50268032311, 50268042612, 50268047615, 50268062111, 50268062115, 50268062211, 50268062215, 50268069415, 50268076111, 50268076211, 50268076311, 50268076312, 50383004224, 50383004248, 50419000233, 50419017100, 50419017106, 50419020801, 50419038501, 50419039001, 50419039101, 50419039201, 50419048858, 50419051106, 50436013401, 50436075901, 50436432401, 50436432502, 50742018101, 50742018425, 50742018921, 50742040205, 50742040620, 50742042802, 50742043108, 50742043810, 50742046316, 50742048505, 50742051220, 50742051902, 50881000560, 50881001560, 50881002060, 50881002560, 50881002701, 50881002801, 51079032120, 51079032301, 51079032306, 51079043420, 51079047201, 51079047205, 51079051005, 51079052056, 51079052501, 51079058101, 51079058201, 51079058205, 51079067001, 51079067005, 51079069201, 51138003730, 51138005130, 51138014430, 51138014530, 51138014621, 51138014636, 51138014642, 51138014710, 51138014712, 51138014714, 51138014715, 51138014720, 51138014730, 51138015520, 51138015521, 51138015528, 51138015536, 51138015539, 51138015542, 51138015550, 51138015614, 51138015615, 51138015620, 51138015621, 51144000160, 51144000212, 51144000260, 51285036601, 51285036901, 51655040043, 51662126301, 51662126303, 51662126401, 51672133805, 51672406301, 51672409103, 51672413806, 51808011901, 51862008514, 51862008551, 51862008614, 51862033301, 51862044601, 51862044630, 51862044705, 51862044718, 51862045090, 51862045847, 51991000533, 51991006598, 51991037690, 51991056001, 51991062033, 51991073532, 51991074990, 51991075910, 51991079798, 51991082228, 51991082328, 51991089233, 51991093798, 51991094298, 52125026802, 52343013701, 52544009276, 52544015302, 52544015602, 52547080130, 52584011312, 52584016505, 52584041401, 52584042100, 52652200106, 52959012600, 52959012605, 52959012607, 52959012620, 52959012621, 52959012640, 52959012642, 52959012644, 52959012650, 52959012660, 52959012670, 52959012712, 52959012715, 52959012720, 52959012742, 52959022020, 52959022040, 52959022075, 52959039228, 52959039230, 52959054710, 52959054711, 52959054720, 52959054730, 52959054750, 53150031401, 53150031410, 53150031501, 53150031701, 53150032001, 53217019301, 53217023130, 53217028815, 53217030006, 53489013810, 53489013910, 54092006301, 54092006401, 54288010001, 54482005301, 54505033105, 54505033210, 54505033310, 54569032200, 54569032404, 54569032700, 54569033101, 54569033104, 54569033105, 54569033201, 54569033202, 54569033203, 54569033209, 54569033601, 54569080900, 54569080902, 54569081103, 54569081200, 54569081301, 54569155500, 54569155501, 54569181809, 54569304000, 54569304300, 54569304301, 54569304302, 54569304306, 54569304307, 54569341300, 54569376501, 54569380602, 54569402604, 54569464801, 54569472800, 54569490700, 54569490702, 54569490800, 54569571600, 54569584000, 54569612402, 54569616800, 54569616802, 54569616803, |

| | |
|---|---|
| | 54569619800, 54569620802, 54868010900, 54868010903, 54868010906, 54868010908, 54868021800, 54868021801, 54868021803, 54868021804, 54868021807, 54868021808, 54868023400, 54868023500, 54868023502, 54868025802, 54868025803, 54868025804, 54868025806, 54868025807, 54868025808, 54868025809, 54868028400, 54868029004, 54868036500, 54868036502, 54868036503, 54868045100, 54868045102, 54868059000, 54868060500, 54868076800, 54868083600, 54868083601, 54868083602, 54868083603, 54868083604, 54868083608, 54868087101, 54868090801, 54868090804, 54868090805, 54868091600, 54868092301, 54868101001, 54868101003, 54868111901, 54868111902, 54868111904, 54868112600, 54868112602, 54868112603, 54868112604, 54868118300, 54868118302, 54868118303, 54868118307, 54868118308, 54868118309, 54868118500, 54868162900, 54868162901, 54868162902, 54868174304, 54868252200, 54868252201, 54868270201, 54868298400, 54868298500, 54868298501, 54868298503, 54868300403, 54868305000, 54868315701, 54868318800, 54868318902, 54868334400, 54868334801, 54868362300, 54868363700, 54868363701, 54868382600, 54868382604, 54868382605, 54868382609, 54868389600, 54868389601, 54868389602, 54868392400, 54868403000, 54868403002, 54868403100, 54868414200, 54868414202, 54868414204, 54868414205, 54868414206, 54868414300, 54868417001, 54868428701, 54868428702, 54868428703, 54868433901, 54868433903, 54868437000, 54868437002, 54868450300, 54868462800, 54868471600, 54868474800, 54868474901, 54868477300, 54868477302, 54868477303, 54868495201, 54868503900, 54868508901, 54868508902, 54868508904, 54868521300, 54868521800, 54868521801, 54868522900, 54868523000, 54868523101, 54868523103, 54868526000, 54868526001, 54868526002, 54868526003, 54868526009, 54868526100, 54868528201, 54868528202, 54868528900, 54868528901, 54868530500, 54868532500, 54868535002, 54868535003, 54868535400, 54868538500, 54868547400, 54868563600, 54868563602, 54868566702, 54868567301, 54868573801, 54868573802, 54868573803, 54868574900, 54868575900, 54868588701, 54868588800, 54868590300, 54868593400, 54868593401, 54868609900, 54868613300, 54868619900, 54868625200, 54868630700, 54868630800, 54868662401, 54879000308, 54879002201, 55045125909, 55045324804, 55111011381, 55111013581, 55111013781, 55111015313, 55111015330, 55111015430, 55111015611, 55111017290, 55111049660, 55111049704, 55111055430, 55111064730, 55111068507, 55150012502, 55150012620, 55150018605, 55150023701, 55150026303, 55150026605, 55150038601, 55154287605, 55154394205, 55154473108, 55154491400, 55154495000, 55154936405, 55154937105, 55289016007, 55289016013, 55289016030, 55289016040, 55289016042, 55289016050, 55289026630, 55289035205, 55289035207, 55289035212, 55289035214, 55289035215, 55289037321, 55289037360, 55289037372, 55289043820, 55289043821, 55289043836, 55289043838, 55289043840, 55289043860, 55289055905, 55289058228, 55289060321, 55289060330, 55289064930, 55289076190, 55289090310, 55289090320, 55289090842, 55292081155, 55292091151, 55390000901, 55390003010, 55390003210, 55390003310, 55390003410, 55390005110, 55390005401, 55390006002, 55390008301, 55390008401, 55390008501, 55390010810, 55390011405, |

| | |
|---|---|
| | 55390012101, 55390013210, 55390016010, 55390016110, 55390016210, 55390016301, 55390021010, 55390021501, 55390021601, 55390021701, 55390021801, 55390023801, 55390025010, 55390025301, 55390025801, 55390029101, 55390029201, 55390029301, 55390030701, 55390030710, 55390030803, 55390037010, 55390037610, 55390037701, 55390045301, 55390049101, 55390080610, 55390080901, 55390081810, 55390082401, 55390082501, 55390082601, 55513000204, 55513000304, 55513000504, 55513001001, 55513001101, 55513001104, 55513001201, 55513001204, 55513001301, 55513001304, 55513002101, 55513002301, 55513002701, 55513002704, 55513003701, 55513003901, 55513004101, 55513004104, 55513004301, 55513004304, 55513004401, 55513004601, 55513004801, 55513005301, 55513005304, 55513005704, 55513009001, 55513009201, 55513009401, 55513009501, 55513009701, 55513012601, 55513012610, 55513014410, 55513016001, 55513020901, 55513020910, 55513020991, 55513022401, 55513047810, 55513048802, 55513052001, 55513053010, 55513073001, 55513095401, 55513095601, 55566830101, 55566830102, 55566830301, 55566840102, 55700004921, 55700006410, 55700006412, 55700006420, 55700020314, 55700020315, 55700020815, 55700020820, 55700020915, 55700020948, 55700048430, 55700052230, 55700062710, 55700062712, 55700067210, 57237006205, 57237006230, 57237006290, 57237007530, 57237007810, 57881044410, 57881044801, 57881044810, 57894015012, 57894015025, 57894015512, 57894019515, 57894050205, 57894050220, 57894050301, 57902024905, 57962001428, 57962014009, 57962014012, 57962028028, 57962056028, 58118024008, 58160083043, 58177036456, 58177091005, 58178001701, 58181303205, 58181304005, 58181304105, 58181304205, 58181304305, 58463001601, 58463001701, 58468010001, 58468010002, 58468014001, 58468018002, 58468018101, 58468018102, 58468035701, 58468184904, 58468784003, 58517020030, 59148004670, 59148004791, 59148007091, 59212011114, 59212070012, 59212070048, 59212070112, 59353000210, 59353000310, 59353000410, 59353022010, 59366280702, 59572020597, 59572021015, 59572021095, 59572021513, 59572021593, 59572022016, 59572030101, 59572040200, 59572040228, 59572040528, 59572041000, 59572041028, 59572041521, 59572042021, 59572042521, 59572050100, 59572050300, 59572072012, 59572073014, 59572074007, 59572077501, 59630022230, 59630022290, 59630070048, 59630070114, 59630070148, 59630070214, 59630071008, 59651020460, 59651020508, 59651023630, 59651023690, 59651024130, 59676003056, 59676003084, 59676004028, 59676004056, 59676005028, 59676030302, 59676030400, 59676030401, 59676030402, 59676031001, 59676031002, 59676031204, 59676060012, 59676061001, 59676096002, 59746000103, 59746000204, 59746000314, 59746001504, 59746017106, 59746017110, 59746017206, 59746017210, 59746017309, 59746017310, 59746017506, 59746017509, 59746017510, 59762005501, 59762005801, 59762005802, 59762007401, 59762085007, 59762332701, 59762374001, 59762374004, 59762374101, 59762374202, 59762374208, 59923060110, 59923060310, 59923070125, 59923070205, 59923070414, 59923070505, 59923070614, 59923070814, 59923070905, 59923071014, 59923071305, 59923071505, 59923072430, 60429001501, 60429013010, 60429013101, |

| | |
|---|---|
| | 60429013110, 60429013201, 60429013210, 60429017701, 60429017705, 60429022630, 60429027218, 60429032701, 60429032801, 60429032805, 60429043301, 60429083401, 60429083405, 60429083505, 60429091030, 60432014050, 60432021208, 60505035401, 60505074406, 60505076402, 60505131103, 60505264201, 60505290009, 60505290103, 60505387709, 60505463103, 60505611000, 60505611306, 60505611400, 60505611502, 60505613000, 60505613005, 60505613207, 60505613208, 60505617708, 60687010511, 60687010521, 60687011221, 60687013211, 60687013221, 60687014501, 60687022711, 60687022794, 60687025240, 60687025246, 60687028611, 60760017935, 60760030610, 60760063610, 60760063710, 60760063715, 60760063730, 61314030401, 61314030410, 61314031210, 61314086601, 61570007401, 61570018001, 61570018101, 61570018201, 61703030346, 61703030916, 61703030926, 61703031922, 61703032322, 61703032518, 61703032618, 61703033218, 61703033918, 61703033922, 61703033950, 61703033956, 61703034222, 61703034318, 61703034366, 61703034916, 61703034936, 61703035009, 61703036018, 61703036022, 61703036135, 61703036250, 61703036322, 61703040825, 61703040841, 61748030113, 61748030213, 61748030311, 61748030413, 61755000801, 61919008710, 61919008730, 61919008760, 61919021810, 61919023515, 61919023521, 61919032610, 61919032615, 61919032621, 61919034221, 61919036521, 61919037330, 61919040421, 61919046430, 61919046530, 61919054505, 61919054506, 61919054510, 61919056512, 61919073390, 61919098630, 61919099507, 61958170101, 62175024124, 62175024224, 62175024324, 62175024419, 62484002002, 62559017331, 62559067030, 62559092114, 62559092214, 62559092451, 62559092551, 62584082721, 62756009440, 62756013001, 62756013101, 62756018101, 62756023920, 62756024064, 62756024083, 62756025013, 62756025083, 62756034844, 62756035040, 62756035144, 62756035666, 62756043860, 62756045236, 62756053360, 62756058140, 62756058142, 62756061460, 62756097460, 62856017708, 62856038901, 62856060122, 62856060301, 62856060422, 62856070430, 62856070805, 62856071005, 62856071405, 62856071430, 62856072005, 62856072030, 62856072430, 62856079601, 62935022205, 62935045345, 62935075275, 63020004901, 63020053630, 63187000230, 63187002010, 63187002036, 63187003721, 63187006530, 63187024305, 63187024308, 63187026590, 63187026610, 63187026615, 63187026620, 63187030027, 63187030036, 63187030042, 63187037910, 63187037912, 63187037920, 63187038230, 63187038305, 63187046664, 63187051320, 63187051330, 63187056110, 63187063601, 63187063610, 63187063630, 63187070906, 63187080706, 63187080712, 63187080718, 63187080730, 63304015801, 63304015830, 63304096201, 63323010161, 63323010351, 63323010405, 63323010450, 63323011718, 63323011720, 63323011728, 63323011759, 63323011768, 63323011769, 63323012302, 63323012550, 63323012553, 63323012594, 63323012600, 63323012930, 63323013212, 63323013720, 63323014010, 63323014212, 63323016516, 63323016526, 63323017215, 63323017420, 63323019302, 63323019352, 63323019410, 63323019420, 63323019606, 63323025503, 63323025803, 63323027810, 63323031904, 63323036501, 63323036504, 63323037601, 63323037604, 63323037701, 63323037704, 63323037805, 63323050616, 63323057270, |

| | |
|---|---|
| | 63323065010, 63323065017, 63323065027, 63323067389, 63323071050, 63323071100, 63323071505, 63323073310, 63323073510, 63323075010, 63323076020, 63323076294, 63323076305, 63323076306, 63323076350, 63323076352, 63323077139, 63323082520, 63323088310, 63323096198, 63459010310, 63459010450, 63459030547, 63459039008, 63459039602, 63459060106, 63459091012, 63459091015, 63459091017, 63459091018, 63459091215, 63459091236, 63539018911, 63629157900, 63629157902, 63629157904, 63629157909, 63629158501, 63629158504, 63629158703, 63629158704, 63629158708, 63629160505, 63629160506, 63629160507, 63629160508, 63629186201, 63629206801, 63629261201, 63629374203, 63629374204, 63629374209, 63629401401, 63629402301, 63629402302, 63629402303, 63629402304, 63629402305, 63629402308, 63629412701, 63629412901, 63629430601, 63629430603, 63739016110, 63739016510, 63739026942, 63739033310, 63739051810, 63739051910, 63739093211, 63874041321, 64067021601, 64116001101, 64116001112, 64205004830, 64370053201, 64370053202, 64679063201, 64679063302, 64679066101, 64679066103, 64679072701, 64679081008, 64720019802, 64720033210, 64720033310, 64842102001, 64842102002, 64842102003, 64842102501, 64980027712, 64980033305, 64980033314, 64980033414, 64980033505, 64980033605, 64980033614, 64980040403, 64980041812, 64980050924, 65162005703, 65162005710, 65162066788, 65162069179, 65162075003, 65162080114, 65162080151, 65162080214, 65162080314, 65162080351, 65162084306, 65174088000, 65174088050, 65219016010, 65293037302, 65293037420, 65483011607, 65580025101, 65597040228, 65597040601, 65862014901, 65862014905, 65862018703, 65862018705, 65862018710, 65862018803, 65862018830, 65862020804, 65862039010, 65862039019, 65862039110, 65862068630, 65862068690, 66215001660, 66220001703, 66220011001, 66267006712, 66267006721, 66267014010, 66336009421, 66336021930, 66336079310, 66658011201, 66658011206, 66733094823, 66733095823, 66758004001, 66758004008, 66758004301, 66758004401, 66758004501, 66758004701, 66758004702, 66758004801, 66758004802, 66758005001, 66758005002, 66758005003, 66758095002, 66758095003, 66758095004, 66993021238, 66993066102, 66993066130, 66993084435, 66993084462, 66993084635, 67253019103, 67253019110, 67253032010, 67253032036, 67296024701, 67296024702, 67296032601, 67296119403, 67296150506, 67386041151, 67386081155, 67457014800, 67457014810, 67457020725, 67457021501, 67457022110, 67457023901, 67457024600, 67457031725, 67457039300, 67457039400, 67457041800, 67457041901, 67457042000, 67457042010, 67457042100, 67457042130, 67457042200, 67457042254, 67457042312, 67457042920, 67457043010, 67457043451, 67457044022, 67457044120, 67457044917, 67457045110, 67457045450, 67457048101, 67457048300, 67457048310, 67457048430, 67457048599, 67457048604, 67457048699, 67457049215, 67457049461, 67457051399, 67457051920, 67457052810, 67457053208, 67457060020, 67457060130, 67457061730, 67457066205, 67457084744, 67457086301, 67457088910, 67457092005, 67457099620, 67877017130, 67877018420, 67877028830, 67877053707, 67877053807, 67877053814, 67877053914, 67877054014, 67877054107, 67877054114, 67877063390, 67877075360, 67877075560, |

| | |
|---|---|
| | 67979000101, 67979000102, 67979050001, 68001015508, 68001024604, 68001024616, 68001024617, 68001024717, 68001026522, 68001026523, 68001026627, 68001026630, 68001026632, 68001028222, 68001028327, 68001028425, 68001028434, 68001028528, 68001028536, 68001028540, 68001028638, 68001028639, 68001031356, 68001034136, 68001034234, 68001034836, 68001035937, 68001037132, 68001039179, 68001041737, 68001042237, 68001042485, 68001043725, 68001044226, 68001046836, 68001046837, 68001048485, 68001048907, 68001049005, 68001049104, 68047070201, 68047070235, 68047070251, 68071154701, 68071186601, 68071190401, 68084014901, 68084014911, 68084017401, 68084017411, 68084017521, 68084022111, 68084024701, 68084024711, 68084028411, 68084032511, 68084037411, 68084039901, 68084039911, 68084046901, 68084055311, 68084055321, 68084078921, 68084078925, 68084078995, 68084080311, 68084092125, 68084092411, 68084093521, 68094032559, 68094076359, 68152010303, 68152011401, 68180039009, 68180069001, 68258890305, 68258898702, 68382011006, 68382020906, 68382020910, 68382022406, 68382022410, 68382036306, 68382077501, 68382082701, 68382091506, 68382091777, 68382091818, 68382091911, 68387017001, 68387024025, 68387024115, 68462015811, 68462015813, 68682000310, 68682000431, 68727074501, 68788459302, 68788697601, 68788697603, 68788714203, 68788720704, 68788726700, 68788726703, 68788734302, 68788783302, 68788789201, 68788893703, 68788909602, 68788940203, 68788967003, 68788989201, 68791010404, 68817013450, 69097051607, 69097091591, 69097094808, 69101041001, 69238101703, 69238117503, 69238117603, 69238142301, 69315018403, 69315018512, 69315018524, 69448000211, 69448000338, 69448000512, 69448000531, 69448000533, 69448000534, 69448000538, 69468015104, 69468015110, 69468015120, 69488000301, 69639010101, 69639010301, 69639010305, 69639012001, 69656010102, 69668012020, 69918072010, 69945045102, 69945045200, 69945045220, 70020191001, 70069002125, 70069002225, 70069002510, 70121100001, 70121100101, 70121100105, 70121116801, 70121116901, 70121123901, 70121124001, 70121145105, 70121146302, 70121157201, 70121157301, 70121157405, 70121164401, 70121165105, 70121165201, 70121165401, 70121165501, 70255001002, 70437024026, 70518030600, 70518051600, 70518142200, 70518202300, 70710152509, 70710153001, 70710153101, 70710161006, 70720095036, 70720095130, 70756081622, 70771134901, 70860020005, 70860020017, 70860020050, 70860020110, 70860020650, 70860021610, 70860021803, 70860021805, 70860078010, 70934009530, 70934009542, 70934009620, 70934010121, 70934029130, 70934058030, 70954005940, 70954006010, 70954006020, 70954006030, 71205040305, 71205040312, 71205042110, 71205042130, 71258001502, 71287021902, 71288010005, 71288010015, 71288010120, 71288010410, 71288010518, 71288010610, 71288010920, 71288011310, 71288011450, 71777039001, 71777039101, 71779012502, 71839010401, 71921019033, 71930001730, 71930001752, 71930001830, 72064021060, 72064021090, 72143023130, 72143023430, 72187040101, 72205002601, 72205005030, 72205005401, 72205006201, 72205006301, 72205008130, 72237010101, 72237010103, 72237010104, 72237010105, 72237010202, 72237010207, |

| | |
|---|---|
| | 72266010101, 72266012510, 72266012601, 72266012610, 72266012801, 72485020330, 72485020460, 72485020512, 72485021008, 72485021401, 72485021730, 72485021930, 72493010303, 72603010101, 72603010502, 72603010601, 72603020001, 72603032601, 72606055401, 72606055601, 72606055701, 72606055801, 72607010000, 72611071601, 72647033104, 72694051501, 72694095401, 72893001524, 72974012001, 73150020012, 73207010130, 73380470001, 73462010101, 73657002001, 75840011101, 75987002201, 75987011111, 75987014013, 76045010320, 76045010610, 76075010201, 76189053530, 76189053560, 76282041230, 76282041290, 76310011001, 76346007301, 76346007302, 76420007701, 79043020025, 79672082502, 79952011001, 81561041305, 89141044801, 89141044830, 99207026012, 2298026, 2397760, 2416530, 2416579, 2448354, 2621654, 2719001, 2750101, 2764001, 2767801, 2771601, 2892601, 3029328, 3031520, 3052411, 3052811, 3083050, 3232711, 3377412, 4015649, 4016949, 4023909, 4024009, 4035039, 4035239, 4035730, 4036030, 4036530, 4110020, 6007201, 6007228, 6007231, 6007258, 6007282, 6046106, 6046206, 6046230, 6046401, 6046410, 6056840, 6302902, 6306101, 6306102, 6386203, 6388432, 6394132, 6410901, 6410902, 6410906, 6410909, 6411901, 6411903, 6412101, 6412102, 7326031, 7326101, 7440101, 8117901, 8451001, 9000501, 9001103, 9001201, 9001306, 9001820, 9003101, 9003906, 9003930, 9004401, 9004704, 9004725, 9004726, 9004727, 9005002, 9005011, 9005602, 9005603, 9005604, 9019009, 9027401, 9028002, 9028603, 9030602, 9030624, 9069802, 9076502, 9082501, 9090916, 9091205, 9111102, 9307301, 9307303, 9347501, 9347503, 9347523, 9752904, 9766304, 13013202, 13111683, 13113691, 13114691, 13115679, 13117687, 13128683, 13220001, 13220101, 13220201, 13252686, 13259691, 15050301, 15191012, 15191113, 15301260, 15303220, 15321130, 15321330, 15321430, 15321530, 15321630, 15335222, 15335322, 19945005, 19945106, 19945200, 23590204, 23590412, 24022205, 24060545, 24065601, 24079375, 24515010, 24515175, 24582411, 24584001, 24584101, 24584301, 24584305, 37122150, 46110091, 46110181, 46110481, 46110491, 51002121, 52060202, 54001720, 54001820, 54006447, 54032006, 54032106, 54032203, 54032206, 54032503, 54038325, 54039513, 54039522, 54048014, 54048114, 54049714, 54317757, 54413025, 54417925, 54418625, 54449705, 54449710, 54455015, 54455025, 54458111, 54458127, 54460425, 54472825, 54474125, 54474131, 54474225, 54817425, 54817625, 54817925, 54818025, 54818125, 54818325, 54855025, 54873925, 54981729, 54982831, 69006701, 69007001, 69007401, 69007601, 69010901, 69013601, 69014501, 69014601, 69014702, 69015111, 69015202, 69015301, 69015501, 69016901, 69016902, 69017001, 69017802, 69017902, 69018102, 69018921, 69019201, 69019301, 69019730, 69020101, 69024901, 69028403, 69029201, 69029310, 69029410, 69029630, 69032401, 69034201, 69048603, 69077038, 69083038, 69098038, 69119530, 69130510, 69130610, 69130910, 69131810, 69134005, 69134016, 69303120, 69303220, 69303420, 69385710, 69402625, 69403101, 69403301, 69403401, 69449522, 69454101, |

| | |
|---|---|
| | 69454301, 69914111, 69914122, 69914211, 69914222, 69914411, 69932122, 74056111, 74056607, 74057611, 74057622, 74057634, 74057928, 74310832, 74646332, 75800120, 78001705, 78010205, 78010961, 78011022, 78018101, 78018161, 78018325, 78024115, 78024661, 78024861, 78024915, 78027422, 78034061, 78034161, 78038725, 78040134, 78052687, 78056661, 78056761, 78059251, 78059451, 78062051, 78062061, 78064070, 78064781, 78064930, 78065206, 78066961, 78067066, 78067119, 78067301, 78067515, 78067615, 78068019, 78068266, 78069802, 78069819, 78069851, 78069899, 78070802, 78070891, 78071502, 78071591, 78081881, 78086001, 78086714, 78086742, 78087421, 78087463, 78090961, 78091661, 85053901, 85057102, 85111001, 85116801, 85125401, 85128703, 85131201, 85131202, 85136604, 85138101, 85141702, 85141703, 85142502, 85142504, 85143001, 85143002, 85143003, 85143004, 85143005, 85151901, 85151904, 85151905, 85300403, 85434701, 85435001, 85435201, 88120243, 88120329, 88120343, 88120926, 93022001, 93078205, 93078210, 93078256, 93078406, 93078486, 93112589, 93551006, 93565598, 93723619, 93723633, 93730165, 93730219, 93730265, 93735501, 93747306, 93748520, 93753656, 93759957, 93760057, 93760141, 93760157, 93762056, 93762998, 93763841, 93763857, 93763957, 93766356, 93776724, 95008921, 115143810, 115147659, 115167572, 115167573, 115703701, 121075908, 121077308, 143147310, 143147325, 143147501, 143242201, 143242230, 143242330, 143920201, 143920301, 143920401, 143921801, 143927701, 143930601, 143930701, 143938401, 143953101, 143954801, 143954901, 143954910, 143955001, 143955201, 143955401, 143955501, 143955801, 143956501, 143958301, 143959721, 143970001, 143970101, 143970201, 143973805, 143974010, 143974401, 143974410, 143974501, 143975001, 143983001, 143989010, 143989101, 172496070, 172524060, 172731046, 172731100, 172731246, 172731320, 173044200, 173044202, 173044600, 173044602, 173044700, 173057000, 173063535, 173071215, 173082102, 173084608, 173084708, 173084765, 173084913, 173089601, 178058208, 182186489, 185093287, 185093330, 185093387, 187090201, 187122103, 187320447, 245057501, 310020130, 310062528, 310062560, 310065758, 310066812, 310066860, 310070539, 310072010, 310072025, 310072050, 310095130, 310135095, 310461150, 310772010, 310783030, 310784030, 338006701, 338008001, 338008601, 338130503, 338176241, 378001401, 378014405, 378014491, 378027401, 378031593, 378064010, 378064110, 378064201, 378064205, 378145205, 378145401, 378145805, 378145877, 378204201, 378207193, 378224577, 378224693, 378251191, 378309785, 378309885, 378315101, 378315177, 378315193, 378326694, 378354725, 378354752, 378471022, 378471522, 378473022, 378479106, 378500193, 378503677, 378503693, 378526214, 378526314, 378526414, 378526598, 378603477, 378603493, 378661193, 378661285, 378661488, 378686901, 378692078, 378692191, 378701705, 378701793, 378709601, 378713193, 378713393, 378773293, 378773493, 378773497, 378800977, 378800993, 378904505, 409018125, 409018201, 409018225, 409018501, 409018701, 409020102, 409020110, 409020120, 409030225, 409032320, |

| | |
|---|---|
| | 409036701, 409036801, 409111201, 409250410, 409321805, 409421501, 409475503, 409475901, 409476013, 409485605, 430072024, 430072124, 430072224, 440816512, 440816715, 469012599, 517044001, 517074501, 517074601, 517092008, 517490525, 527145006, 527145106, 527293137, 527293243, 527293341, 548540125, 555030102, 555030138, 555044605, 555044609, 555048401, 555057235, 555060702, 555077902, 555077904, 555080802, 555087304, 555088202, 555088604, 555088702, 555090405, 555105756, 574010601, 574010603, 574079201, 574086610, 574087005, 574087205, 591048701, 591048705, 591048805, 591052801, 591079001, 591079021, 591222315, 591222455, 591223330, 591236710, 591243415, 591247260, 591247319, 591289749, 591359160, 591413054, 591505210, 591505221, 591505243, 591544201, 591544205, 591544210, 591544221, 591544301, 591544310, 597013730, 597013830, 603114756, 603156756, 603156758, 603389919, 603390021, 603418016, 603533521, 603533528, 603533721, 603533731, 603533732, 603533831, 603533832, 603533932, 603938856, 641607901, 641608001, 641608025, 641614601, 641614625, 641617410, 641617501, 641617510, 703004501, 703006301, 703024301, 703301812, 703301912, 703306711, 703306911, 703321381, 703321801, 703324911, 703331104, 703332101, 703332104, 703352401, 703367101, 703367191, 703367193, 703367301, 703367801, 703398601, 703400401, 703409401, 703415511, 703415591, 703418201, 703418391, 703423901, 703423981, 703424601, 703424801, 703424881, 703424891, 703440211, 703441211, 703443281, 703443411, 703450204, 703450294, 703468601, 703476401, 703476801, 703476881, 703480503, 703504001, 703504301, 703504303, 703504601, 703514001, 703514591, 703523313, 703523391, 703565601, 703565701, 703565791, 703573001, 703574711, 703577801, 703722101, 703722103, 703787103, 703797101, 703797103, 781167933, 781168133, 781232151, 781232246, 781232368, 781269144, 781269175, 781269244, 781269275, 781269375, 781269544, 781269575, 781269675, 781301072, 781301095, 781301180, 781303075, 781303175, 781308475, 781313195, 781313692, 781313775, 781313980, 781316575, 781323394, 781324572, 781324575, 781328275, 781331275, 781331780, 781349212, 781349775, 781400332, 781406336, 781502207, 781523801, 781523806, 781523864, 781523901, 781523906, 781523964, 781523980, 781540931, 781916575, 781925394, 781931570, 781931780, 832008600, 832028500, 832059530, 904267460, 904357161, 904601946, 904619546, 904620846, 904620946, 904655161, 904657461, 904694804, 904714504, 944262001, 944262002, 944262004, 944265504, 944265603, 955102104, 955102208, 955172720, 955173320, 955174601, 10019090502, 10019090503, 10019090517, 10019090603, 10019090605, 10019090663, 10019093601, 10019093901, 10019094201, 10019094401, 10019095301, 10019095601, 10019095701, 10139006312, 10139006350, 10370026801, 10544021206, 10544057506, 10544091330, 10544091421, 10544091510, 10544091530, 10631000531, 10631000731, 10631011531, 10631011631, 10631011731, 10631011831, 10768728303, 10768728304, 10768773304, 11399000530, 12634068604, 13632012301, 13668046201, 13668059284, 13668059487, 13925016604, 13925052301, 15054004301, 15054012001, 15054109004, 16477050521, 16590026910, 16590029137, 16590032615, 16590032620, |

| | |
|---|---|
| | 16590037330, 16590087921, 16714000101, 16714008801, 16714008825, 16714008901, 16714009001, 16714012001, 16714022112, 16714022130, 16714046801, 16714047201, 16714050001, 16714052201, 16714052203, 16714052204, 16714052205, 16714052210, 16714057102, 16714067101, 16714067102, 16714072701, 16714072801, 16714074201, 16714077701, 16714081601, 16714081602, 16714085601, 16714085901, 16714088601, 16714090801, 16714090901, 16714092701, 16729002310, 16729003410, 16729003415, 16729003515, 16729003516, 16729004854, 16729004954, 16729005053, 16729005153, 16729007212, 16729009001, 16729009010, 16729009015, 16729009203, 16729010811, 16729011431, 16729011838, 16729012049, 16729013054, 16729015131, 16729022361, 16729022850, 16729023163, 16729024003, 16729024605, 16729024711, 16729024838, 16729026763, 16729027603, 16729027611, 16729027638, 16729027667, 16729027668, 16729027703, 16729028811, 16729028838, 16729029531, 16729029534, 16729029805, 16729033205, 16729039130, 16729042333, 16729042605, 16729048601, 17478032745, 17478054601, 17478054701, 17478076306, 17856069102, 17856075904, 17856075905, 17856372201, 20482033530, 20536032201, 21695011130, 21695029030, 21695030621, 21695030636, 21695030650, 21695030710, 21695030715, 21695030721, 21695038030, 21695038208, 21695061300, 21695062300, 21695072812, 21695074512, 21695076421, 21695076548, 21695083404, 21695083415, 21695083530, 21695089630, 23155016831, 23155017031, 23155019641, 23155019642, 23155019643, 23155021331, 23155021431, 23155026141, 23155037841, 23155052831, 23155054741, 23155054842, 23155055031, 23155064941, 23155068531, 23594050521, 23594050548, 24987011114, 25021020111, 25021020166, 25021020167, 25021020168, 25021020169, 25021020215, 25021020245, 25021020325, 25021020401, 25021020405, 25021020505, 25021020606, 25021020751, 25021020810, 25021020950, 25021021120, 25021021350, 25021021402, 25021021405, 25021022160, 25021022201, 25021023002, 25021023120, 25021023310, 25021023320, 25021023410, 25021023550, 25021023604, 25021023706, 25021023905, 25021024110, 25021024202, 25021024504, 25021045101, 25021045201, 25021045505, 25021077702, 25021077901, 25021078104, 25021078305, 25021078874, 25021080167, 25021080210, 25021080310, 25021080705, 25021081030, 25021081310, 25021081366, 25021081430, 25021081567, 25021081630, 25021082406, 25021082667, 25021082850, 29336061024, 30698001730, 30698020201, 31722013190, 31722052501, 31722052530, 31722052590, 31722096160, 31722096260, 33261012900, 33261047501, 33261062510, 33261074701, 33358049830, 35356019730, 35356025000, 35356025100, 35356067321, 35356067330, 35356067415, 35356067420, 35356067721, 35356067728, 35356067730, 35356067760, 35356081820, 35356081821, 35356081830, 35356081915, 35356081921, 35356081930, 35356081942, 35356085306, 35356085309, 35356085310, 35356085320, 35356085330, 36000001205, 36000001406, 39822218001, 41616048583, 41616093640, 42023011001, 42023011101, 42023013401, 42023013501, 42043039002, 42043039021, 42195012707, 42195014912, 42195022106, 42238011101, 42254007710, 42254016130, 42254021210, 42254024330, 42291002412, 42291008590, 42291016712, 42291016830, 42291019112, |

| | |
|---|---|
| | 42291025790, 42291028030, 42291028090, 42291032101, 42291035230, 42291037490, 42291044960, 42291045160, 42291059401, 42291072710, 42291076801, 42291084301, 42292000701, 42292000710, 42292005305, 42367012121, 42367012125, 42367012129, 42367052025, 42388001114, 42388001214, 42388002426, 42388002526, 42543014201, 42549064714, 42658002101, 42747032730, 42747072601, 42747076101, 42799081301, 42806008701, 42806008705, 42806008901, 42806008905, 42806035801, 42806035830, 42806040001, 42806040021, 43063005204, 43063009706, 43063010910, 43063020130, 43063020190, 43063020830, 43063026607, 43063027304, 43063038306, 43063038315, 43063038630, 43063041501, 43063041530, 43063042610, 43063042620, 43063042640, 43063042650, 43063043210, 43063043212, 43063043215, 43063043230, 43063043805, 43063043890, 43063044630, 43063056004, 43063056020, 43063059005, 43063059009, 43063059010, 43063059015, 43063059018, 43063059020, 43063059025, 43063059204, 43063059210, 43063064355, 43063064440, 43063070306, 43063070309, 43063070318, 43063070325, 43063070330, 43063070630, 43063074612, 43066000101, 43066000601, 43066001401, 43288010410, 43288010801, 43288010810, 43353065760, 43547025801, 43598025811, 43598025940, 43598028335, 43598030330, 43598030390, 43598030562, 43598034530, 43598035804, 43598039248, 43598050530, 43598061040, 43598068235, 43598068325, 43598077311, 43975025205, 43975025214, 43975025314, 43975025414, 43975025514, 43975025605, 43975025614, 43975025705, 43975030810, 44087400004, 44087400005, 44087400009, 44087500006, 44567050601, 44567050701, 44567051101, 44567053001, 45802007662, 45802036853, 45802073321, 45802073367, 45963044055, 45963050008, 45963060755, 45963060756, 45963061159, 45963061257, 45963061353, 45963061356, 45963061386, 45963061389, 45963061451, 45963061959, 45963062151, 45963063749, 45963064077, 45963073355, 45963073452, 45963073474, 45963079056, 47335003540, 47335004740, 47335008350, 47335015140, 47335017840, 47335028440, 47335028541, 47335032340, 47335040181, 47335047281, 47335047583, 47335058140, 47335058142, 47335071483, 47335071513, 47335071581, 47335089021, 47335089072, 47335089121, 47335089174, 47335089180, 47335089221, 47335089274, 47335089280, 47335089374, 47335089380, 47335092921, 47335092980, 47781059229, 47781059407, 47781059507, 47781060427, 47781060925, 47781061023, 47781062291, 48818000101, 48818000102, 49349094801, 49349096905, 49663000225, 49884008401, 49884008601, 49884011991, 49884012591, 49884012791, 49884012901, 49884029001, 49884032562, 49884036826, 49884075305, 49884086802, 49884092204, 49999000800, 49999000821, 49999000830, 49999000840, 49999002814, 49999002820, 49999002821, 49999002848, 49999002860, 49999002865, 49999005906, 49999005912, 49999005930, 49999010900, 49999010990, 49999011000, 49999011007, 49999011010, 49999011015, 49999011020, 49999045830, 50090009004, 50090009901, 50090016600, 50090016705, 50090022100, 50090027100, 50090029409, 50090029501, 50090031201, 50090043900, 50090049101, 50090055600, 50090094200, 50090100100, 50090101500, 50090101503, 50090120000, 50090120001, 50090120002, 50090158200, 50090160000, 50090165801, 50090165802, |

| | |
|---|---|
| | 50090165803, 50090166000, 50090166001, 50090170402, 50090171800, 50090182300, 50090185303, 50090187600, 50090192200, 50090194100, 50090198901, 50090199000, 50090199002, 50090199007, 50090199100, 50090209800, 50090210301, 50090234200, 50090234509, 50090236300, 50090240304, 50090262400, 50090280400, 50090381100, 50242005110, 50242005121, 50242005306, 50242006001, 50242006101, 50242006301, 50242007701, 50242008701, 50242008801, 50242009001, 50242010801, 50242010886, 50242010901, 50242013001, 50242013201, 50242013210, 50242013468, 50242014001, 50242014501, 50242021060, 50242091886, 50268007515, 50268015413, 50268029011, 50268029015, 50268029115, 50268029211, 50268029215, 50268031415, 50268032315, 50268042712, 50268076112, 50268076212, 50383004004, 50419005014, 50419005030, 50419017101, 50419017103, 50419039501, 50436013101, 50436013201, 50436013301, 50436018701, 50436172001, 50436172005, 50436175004, 50633021011, 50742012330, 50742012390, 50742018130, 50742018212, 50742018224, 50742018324, 50742018901, 50742040102, 50742040401, 50742040510, 50742041605, 50742043001, 50742044505, 50742044615, 50742044745, 50742044860, 50742046450, 50742049417, 50742049525, 50742052005, 50881001060, 50881002601, 51079032101, 51079032156, 51079043401, 51079043501, 51079043520, 51079052001, 51079052020, 51079052030, 51079052401, 51079052420, 51079052520, 51079058106, 51079069203, 51138005030, 51138014515, 51138014520, 51138014615, 51138014620, 51138014628, 51138014639, 51138014650, 51138014721, 51138015415, 51138015420, 51138015430, 51138015515, 51138015610, 51138015612, 51138015630, 51144002001, 51144003001, 51144005001, 51285036701, 51285036801, 51407018112, 51655001203, 51672133803, 51672411806, 51862008314, 51862008351, 51862008414, 51862008451, 51862008651, 51862008714, 51862008751, 51862008851, 51862033201, 51862033305, 51862033401, 51862033405, 51862036240, 51862044605, 51862044610, 51862044710, 51862044760, 51862044918, 51862044960, 51862045030, 51862045801, 51862046001, 51862046047, 51991000590, 51991018801, 51991018831, 51991021898, 51991037733, 51991062010, 51991073520, 51991074933, 51991075933, 51991082128, 51991089033, 51991089133, 51991092298, 51991092398, 51991093898, 52125061952, 52152050002, 52152050008, 52152050030, 52544015402, 52544018876, 52544018976, 52584003930, 52584004725, 52584006900, 52584023805, 52584023930, 52584042010, 52584042200, 52609000105, 52609300100, 52652200101, 52959012610, 52959012612, 52959012615, 52959012618, 52959012625, 52959012630, 52959012707, 52959012710, 52959012718, 52959012721, 52959012725, 52959012730, 52959012737, 52959022000, 52959022010, 52959022021, 52959022030, 52959022036, 52959022060, 52959039212, 52959039221, 52959054712, 52959054716, 52959099103, 53014025001, 53150024701, 53150025001, 53150032010, 53150087101, 53217023121, 53217028810, 53225361001, 53225365001, 53225366001, 53489013801, 53489013850, 53489013901, 53489013905, 53489014001, 53489014005, 53489014010, 53964000101, 53964000202, 54348081904, 54482030101, 54569032203, 54569033000, 54569033001, 54569033003, 54569033004, 54569033007, 54569033100, 54569033102, 54569033107, |

| | |
|---|---|
| | 54569033108, 54569033205, 54569033300, 54569081205, 54569103600, 54569304305, 54569330200, 54569330201, 54569380701, 54569464802, 54569572900, 54569587300, 54569587302, 54569587303, 54569587304, 54569604800, 54569604801, 54569612400, 54569612401, 54569616801, 54569620800, 54569620801, 54569660700, 54771354701, 54838055550, 54868010901, 54868010902, 54868010905, 54868010907, 54868017300, 54868021802, 54868021805, 54868021806, 54868021809, 54868023501, 54868025800, 54868025801, 54868025805, 54868029000, 54868029002, 54868029003, 54868045106, 54868045300, 54868077601, 54868082100, 54868083605, 54868083607, 54868083609, 54868087100, 54868087106, 54868090800, 54868090802, 54868090803, 54868092700, 54868095100, 54868095500, 54868111903, 54868111905, 54868112601, 54868118301, 54868118304, 54868118306, 54868118501, 54868162903, 54868174300, 54868174301, 54868174302, 54868174303, 54868174400, 54868199400, 54868252301, 54868270200, 54868291300, 54868291301, 54868291302, 54868291303, 54868298402, 54868298403, 54868298502, 54868300401, 54868300402, 54868300404, 54868300405, 54868308400, 54868308401, 54868308402, 54868318900, 54868318901, 54868331000, 54868331001, 54868331003, 54868331004, 54868361300, 54868382601, 54868382602, 54868382603, 54868382606, 54868382607, 54868382608, 54868403001, 54868403101, 54868403102, 54868403103, 54868409500, 54868409600, 54868410000, 54868410001, 54868413800, 54868413801, 54868415100, 54868417000, 54868428700, 54868428704, 54868433900, 54868433904, 54868437001, 54868474900, 54868477301, 54868486500, 54868495200, 54868500000, 54868500500, 54868502000, 54868504100, 54868504300, 54868508900, 54868508903, 54868508905, 54868511400, 54868511401, 54868523102, 54868524200, 54868526005, 54868528200, 54868528902, 54868528903, 54868528904, 54868529000, 54868533400, 54868534801, 54868535000, 54868535004, 54868538501, 54868542700, 54868542701, 54868542702, 54868542703, 54868542800, 54868542900, 54868544302, 54868544700, 54868552200, 54868566700, 54868566701, 54868573800, 54868574901, 54868580100, 54868580101, 54868580102, 54868580200, 54868586700, 54868591500, 54868598000, 54868598500, 54868613000, 54868613001, 54868621100, 54868621101, 54868621200, 54868621201, 54868623200, 54868629300, 54868636600, 54879002101, 54879003664, 55045324305, 55045324801, 55045372903, 55045384809, 55111013681, 55111015305, 55111015413, 55111017230, 55111055490, 55111055610, 55111064630, 55111068607, 55111068725, 55111069407, 55135013201, 55150023805, 55150023930, 55150026203, 55150026550, 55150029901, 55150030425, 55150030510, 55150035201, 55150035301, 55150035401, 55154287205, 55154391405, 55154392905, 55154393205, 55154393905, 55154394005, 55154394105, 55154490100, 55154494900, 55154511805, 55154511808, 55154536900, 55154707505, 55154832905, 55154938305, 55154955705, 55289016005, 55289016010, 55289033005, 55289033007, 55289033010, 55289035209, 55289035210, 55289035220, 55289035221, 55289035230, 55289037301, 55289037330, 55289037336, 55289037342, 55289037346, 55289037355, 55289043815, 55289043830, 55289043842, 55289043850, 55289055903, 55289055906, 55289055910, 55289058204, |

| | |
|---|---|
| | 55289058206, 55289058210, 55289060307, 55289060390, 55289076101, 55289076130, 55289081630, 55289090312, 55289090830, 55289092430, 55390001402, 55390003110, 55390004501, 55390004801, 55390005210, 55390005301, 55390006901, 55390007001, 55390009110, 55390010801, 55390011501, 55390012110, 55390012210, 55390012401, 55390013110, 55390013301, 55390013401, 55390013501, 55390014210, 55390014301, 55390016401, 55390020910, 55390021901, 55390023701, 55390025101, 55390025201, 55390025901, 55390026601, 55390030405, 55390031405, 55390034701, 55390037510, 55390039110, 55390039150, 55390045101, 55390045201, 55390049201, 55390049301, 55390080510, 55390080710, 55390080801, 55513000201, 55513000301, 55513000401, 55513000404, 55513000501, 55513000601, 55513001004, 55513001401, 55513001501, 55513002104, 55513002304, 55513002501, 55513002504, 55513002801, 55513003201, 55513003704, 55513003904, 55513005401, 55513005404, 55513005701, 55513005801, 55513005804, 55513007801, 55513007901, 55513009101, 55513009301, 55513009601, 55513009801, 55513009804, 55513011001, 55513011101, 55513013201, 55513014101, 55513014401, 55513014801, 55513014810, 55513019001, 55513019201, 55513020601, 55513020701, 55513026701, 55513026710, 55513028301, 55513028310, 55513032601, 55513047801, 55513048824, 55513052006, 55513053001, 55513054601, 55513054610, 55513071001, 55513082301, 55513082310, 55513092401, 55513092410, 55513092491, 55513095501, 55566840100, 55566840101, 55566840301, 55700006406, 55700006430, 55700007215, 55700007218, 55700007220, 55700007221, 55700007230, 55700020310, 55700020318, 55700020320, 55700020321, 55700020330, 55700020821, 55700020830, 55700020840, 55700020920, 55700020921, 55700020930, 55700020942, 55700045830, 55700045890, 55700050490, 55700062720, 55700062730, 55700063106, 55700063110, 55700063120, 55700085421, 57237007630, 57237007710, 57237007730, 57237007830, 57664002297, 57665033101, 57881044401, 57894019506, 57894042001, 57894042101, 57894050101, 57962007028, 57962042028, 58160083001, 58160083011, 58160083032, 58160083046, 58160083052, 58177036322, 58177036422, 58177091007, 58177093205, 58178001703, 58181303005, 58181303105, 58463001008, 58463001401, 58463001501, 58468003002, 58468035703, 58468782003, 59212070102, 59212070148, 59212070212, 59212070248, 59353001010, 59353012010, 59366281703, 59366282703, 59385004130, 59572010201, 59572020514, 59572020517, 59572020594, 59572022096, 59572040500, 59572041500, 59572042000, 59572042500, 59572050121, 59572050200, 59572050221, 59572050321, 59572050400, 59572050421, 59572070530, 59572071030, 59572071101, 59572073007, 59572074014, 59572098301, 59572098401, 59630070014, 59630070248, 59630071010, 59651018030, 59651018090, 59651018201, 59651024090, 59651029960, 59651030030, 59676030200, 59676030201, 59676030202, 59676030300, 59676030301, 59676031000, 59676031200, 59676031201, 59676032000, 59676032001, 59676032004, 59676034000, 59676034001, 59676060099, 59676096001, 59676096601, 59676096602, 59746000106, 59746000206, 59746017306, 59762004901, 59762005001, 59762005101, 59762005502, 59762007301, 59762007501, 59762085002, 59762085003, 59762285801, |

| | |
|---|---|
| | 59762332702, 59762374005, 59762374104, 59762444002, 59762444003, 59762509101, 59762509301, 59923070100, 59923070202, 59923070305, 59923070705, 59923071105, 59923071214, 59923071402, 59923072160, 59923072212, 59923072390, 59923072530, 59923072630, 59923072730, 60258086603, 60429013001, 60429017730, 60429022605, 60429026201, 60429026501, 60429028630, 60429028690, 60429032901, 60429032905, 60429043305, 60429083301, 60429083501, 60429090960, 60429092590, 60429092630, 60429093530, 60429093590, 60432046600, 60432046608, 60505013300, 60505013400, 60505038105, 60505050105, 60505074401, 60505131203, 60505264203, 60505290109, 60505298503, 60505325503, 60505325508, 60505387703, 60505463003, 60505610501, 60505613206, 60505616600, 60505619301, 60687011211, 60687012201, 60687012211, 60687013401, 60687013411, 60687014511, 60687014994, 60687019221, 60687025286, 60687028621, 60687037501, 60687038621, 60687042801, 60687042865, 60687045521, 60687051101, 60687058201, 60760025540, 60760025560, 60760031310, 60760037321, 60760062921, 60760063620, 60760063630, 60760063720, 60760071521, 61314031201, 61570007201, 61570007301, 61570007550, 61703030436, 61703030538, 61703030906, 61703030925, 61703032418, 61703032722, 61703034106, 61703034109, 61703034209, 61703034250, 61703034365, 61703034418, 61703034909, 61703035010, 61703035037, 61703035038, 61703035959, 61703035993, 61703036050, 61703036318, 61703040822, 61748030111, 61748030211, 61748030313, 61748030411, 61786002757, 61919014921, 61919019830, 61919023510, 61919023540, 61919026921, 61919026955, 61919027830, 61919029137, 61919032142, 61919032620, 61919032623, 61919034210, 61919037321, 61919040430, 61919046410, 61919046510, 61919054530, 61919056530, 61919056590, 61919082721, 61958030101, 61958170201, 62175024219, 62175024519, 62484002001, 62559054015, 62559054055, 62559068030, 62559089030, 62559092151, 62559092251, 62559092314, 62559092414, 62559093001, 62559093101, 62584082711, 62756000860, 62756002340, 62756007360, 62756009444, 62756010260, 62756018201, 62756021960, 62756023886, 62756032160, 62756034944, 62756035240, 62756035664, 62756051183, 62756074660, 62756082640, 62756082740, 62778002701, 62856000110, 62856060001, 62856060210, 62856070830, 62856071030, 62856071230, 62856071805, 62856071830, 62856072405, 62856079701, 62856079801, 62935022305, 62935030230, 62935030330, 62935045245, 62935075375, 63020007801, 63020007802, 63020007901, 63020007902, 63020008001, 63020008002, 63187002015, 63187002021, 63187003720, 63187003730, 63187016021, 63187021564, 63187023610, 63187025620, 63187026630, 63187030006, 63187030015, 63187030021, 63187030028, 63187030040, 63187037903, 63187037905, 63187037915, 63187037930, 63187038210, 63187038220, 63187047401, 63187051310, 63187052603, 63187052610, 63187052630, 63187056115, 63187056130, 63187056160, 63187063603, 63187063606, 63187063612, 63187063615, 63187066101, 63187067030, 63187070910, 63187070915, 63187070930, 63187074130, 63187074190, 63187080221, 63187080705, 63187080707, 63187080710, 63187080715, 63187080720, 63187080721, 63304009530, 63304009630, 63304013530, 63304045830, 63304045930, 63304096230, |

| | |
|---|---|
| | 63323010213, 63323010294, 63323010364, 63323010365, 63323010425, 63323011710, 63323011719, 63323011751, 63323011758, 63323011761, 63323012020, 63323012102, 63323012250, 63323012259, 63323012310, 63323012603, 63323012710, 63323012820, 63323013210, 63323013215, 63323013610, 63323014210, 63323014507, 63323016501, 63323016505, 63323016530, 63323017205, 63323017245, 63323017260, 63323017460, 63323017530, 63323017650, 63323019202, 63323019305, 63323019355, 63323019405, 63323021110, 63323021220, 63323026530, 63323031701, 63323031801, 63323037302, 63323037420, 63323037905, 63323050601, 63323051610, 63323063110, 63323063150, 63323063710, 63323065020, 63323067305, 63323071059, 63323073311, 63323073410, 63323075020, 63323076210, 63323076217, 63323076316, 63323076317, 63323077230, 63323088305, 63323088330, 63323097210, 63459017714, 63459030343, 63459034804, 63459039120, 63459039502, 63459060010, 63459091001, 63459091011, 63459091036, 63459091201, 63459091211, 63459091212, 63459091217, 63459091218, 63459091859, 63539048603, 63539068803, 63629147201, 63629147202, 63629157901, 63629157903, 63629157905, 63629157906, 63629157907, 63629157908, 63629158502, 63629158503, 63629158701, 63629158702, 63629158705, 63629158706, 63629158707, 63629160501, 63629160502, 63629160503, 63629160504, 63629205601, 63629206701, 63629261202, 63629261301, 63629261302, 63629261303, 63629269601, 63629374201, 63629374202, 63629375501, 63629375502, 63629391001, 63629401402, 63629401403, 63629401404, 63629401405, 63629402306, 63629402307, 63629402309, 63629409307, 63629430602, 63629430604, 63629430605, 63629430606, 63629430607, 63629456201, 63717091508, 63739013310, 63739026910, 63739052010, 63739058810, 63739097117, 64205004890, 64205008790, 64205048890, 64380076304, 64380076305, 64380097025, 64380097106, 64380097206, 64679002101, 64679009601, 64679063401, 64679063501, 64679063502, 64679066102, 64679066201, 64679072601, 64720019898, 64720033105, 64842072709, 64842102502, 64842102503, 64980022403, 64980022409, 64980027606, 64980033405, 64980033514, 64980033705, 64980033714, 64980033805, 65162005709, 65162005711, 65162066790, 65162075009, 65162080251, 65162080414, 65162080451, 65162080514, 65162080551, 65162080651, 65162084416, 65174088025, 65293037325, 65293037401, 65597040220, 65862014930, 65862014990, 65862018730, 65862018799, 65862018805, 65862039030, 65862039101, 65862039130, 65862039166, 65862070901, 66267096121, 66302001401, 66435041005, 66435041530, 66530024940, 66658011203, 66758003501, 66758003601, 66758003702, 66758004002, 66758004101, 66758004201, 66758004202, 66758004302, 66758004303, 66758004403, 66758004502, 66758004601, 66758004703, 66758004704, 66993041702, 66993041730, 66993041788, 66993048935, 66993066188, 66993084535, 66993084562, 66993084662, 67108356509, 67386091151, 67457019501, 67457020850, 67457022102, 67457022140, 67457023802, 67457023900, 67457024500, 67457024501, 67457024601, 67457025430, 67457031105, 67457031625, 67457039054, 67457039354, 67457039410, 67457039525, 67457039610, 67457041805, 67457041900, 67457042300, 67457042410, 67457042551, 67457043111, 67457043650, 67457044000, |

| | |
|---|---|
| | 67457044220, 67457044360, 67457044610, 67457045010, 67457045220, 67457045500, 67457045552, 67457046201, 67457046302, 67457046420, 67457046610, 67457046721, 67457046799, 67457046910, 67457047152, 67457047210, 67457047404, 67457047610, 67457047810, 67457047953, 67457048040, 67457048400, 67457048508, 67457049154, 67457049346, 67457051305, 67457051805, 67457052040, 67457052920, 67457053035, 67457053102, 67457057901, 67457060820, 67457060920, 67457061010, 67457061610, 67457061810, 67457078108, 67457083306, 67457086404, 67457088605, 67457092802, 67457099115, 67457099740, 67877016930, 67877017030, 67877017110, 67877018402, 67877028801, 67877028805, 67877028810, 67877028890, 67877053714, 67877053907, 67877054007, 67877054207, 67877063430, 67877075460, 67979050140, 68001000501, 68001015504, 68001024701, 68001024704, 68001024716, 68001024755, 68001026524, 68001026525, 68001026526, 68001026527, 68001026631, 68001028225, 68001028226, 68001028227, 68001028332, 68001028522, 68001028529, 68001028537, 68001034137, 68001034526, 68001034536, 68001035525, 68001036625, 68001037027, 68001037232, 68001038936, 68001039077, 68001041636, 68001041838, 68001042122, 68001044327, 68001044432, 68001048022, 68001048035, 68001048225, 68047070221, 68083029201, 68083038101, 68084007511, 68084007521, 68084017511, 68084022001, 68084022011, 68084022101, 68084028401, 68084032521, 68084037421, 68084044811, 68084044821, 68084046911, 68084061211, 68084061221, 68084078911, 68084080321, 68084087925, 68084087995, 68084092195, 68084092421, 68084093025, 68084093095, 68084093511, 68094032562, 68094076362, 68152010100, 68152010404, 68152010809, 68152010900, 68152011201, 68180039106, 68180080136, 68220005510, 68258301301, 68258706801, 68382011001, 68382022401, 68382022405, 68382038306, 68382082614, 68382082706, 68382091306, 68382091406, 68382091601, 68382091634, 68382091711, 68382091801, 68382099710, 68387024010, 68462010530, 68462010630, 68462015713, 68462031732, 68462079201, 68645044670, 68645054154, 68682008531, 68727071201, 68727074502, 68727074505, 68788147303, 68788641402, 68788697606, 68788714205, 68788740103, 68788917808, 68788932901, 68788989301, 68791010004, 68850000108, 69076091302, 69076091325, 69097031602, 69097043935, 69097051707, 69097080540, 69097083037, 69097092735, 69097094903, 69117000301, 69117000401, 69171039801, 69238116507, 69238117403, 69238125001, 69238142306, 69238175406, 69315018624, 69315018725, 69448000105, 69543037110, 69639010201, 69639010501, 69656010330, 69656010390, 69660020191, 69660020291, 69660020391, 69918072002, 69945045201, 69945045203, 69945045275, 70020191101, 70069002325, 70112055502, 70114010101, 70121100005, 70121104902, 70121104905, 70121109901, 70121123801, 70121124407, 70121148202, 70121156701, 70121157305, 70121157401, 70121163101, 70121165101, 70121165301, 70121165701, 70121165705, 70255002001, 70255002501, 70255002503, 70437024018, 70518007500, 70518108800, 70518153400, 70518202301, 70518245900, 70529004501, 70700016922, 70700017022, 70756081560, 70771135004, 70860020120, 70860020550, 70860020651, 70860020910, 70860021051, 70860021174, 70860021461, 70860077602, |

| | |
|---|---|
| | 70860077720, 70860078210, 70882011721, 70882011730, 70934009520, 70934009521, 70934009610, 70934009615, 70934009630, 70934013705, 70934014804, 70934025930, 70934047121, 70954005710, 70954005810, 70954005820, 70954005830, 70954005840, 70954005910, 70954005920, 70954005930, 70954006110, 71205001221, 71205001349, 71205040710, 71205040718, 71205040720, 71205040721, 71205040730, 71205042118, 71258001522, 71287011902, 71288010045, 71288010051, 71288010110, 71288010720, 71288011290, 71288011728, 71288041810, 71288055586, 71329010501, 71334010001, 71335152501, 71777039201, 71779011502, 71837584305, 71905040011, 71930001852, 72064011030, 72064012030, 72064013030, 72143023330, 72189000821, 72189019890, 72205000660, 72205000792, 72205003092, 72205003101, 72205003601, 72205004501, 72205004601, 72205006101, 72205008030, 72205008230, 72237010102, 72237010106, 72237010107, 72237010206, 72237010305, 72237010401, 72266010801, 72266012001, 72266012101, 72266012325, 72266012401, 72266012501, 72266016101, 72266016201, 72485020101, 72485020290, 72485021102, 72485021205, 72485021315, 72485021504, 72485021608, 72485021830, 72485022102, 72485022210, 72485022320, 72572012225, 72572052025, 72579001101, 72579001102, 72603010301, 72603010401, 72603010701, 72603010801, 72603020201, 72603030101, 72603040101, 72606055501, 72606055902, 72606056601, 72606056901, 72611078502, 72647033101, 72865014030, 72865014090, 72893001506, 73535020801, 74527002202, 75987002001, 75987002101, 75987014014, 75987014513, 75987014514, 76045010120, 76045010910, 76075010101, 76075010301, 76128015575, 76189011318, 76189011321, 76189053430, 76282041205, 76310001750, 76388063525, 76388063550, 76388071325, 76388088025, 79672001801, 79672001901, 79672082602, 89141044401, 89141044430<br><br>OR<br><br>At least one of the following cancer therapy generic names: ABARELIX, ABEMACICLIB, ABIRATERONE ACETATE, ACALABRUTINIB, ADO-TRASTUZUMAB EMTANSINE, ALDESLEUKIN, ALEMTUZUMAB, ALITRETINOIN, ALPELISIB, ALTRETAMINE, ANASTROZOLE, APALUTAMIDE, APREPITANT, ARSENIC TRIOXIDE, ASPARAGINASE, ATEZOLIZUMAB, AVAPRITINIB, AVELUMAB, AXICABTAGENE CILOLEUCEL, AXITINIB, AZACITIDINE, BELANTAMAB MAFODOTIN-BLMF, BELINOSTAT, BENDAMUSTINE HCL, BEVACIZUMAB, BEVACIZUMAB-AWWB, BEVACIZUMAB-BVZR, BEXAROTENE, BICALUTAMIDE, BINIMETINIB, BLEOMYCIN SULFATE, BLINATUMOMAB, BORTEZOMIB, BOSUTINIB, BRENTUXIMAB VEDOTIN, BREXUCABTAGENE AUTOLEUCEL, BRIGATINIB, BROMOCRIPTINE MESYLATE, BUSULFAN, CABAZITAXEL, CALASPARGASE PEGOL-MKNL, CAPECITABINE, CARBOPLATIN, CARFILZOMIB, CARMUSTINE, CEMIPLIMAB-RWLC, CERITINIB, CETUXIMAB, CHLORAMBUCIL, CISPLATIN, CLADRIBINE, CLOFARABINE, CORTISONE ACETATE, CRIZOTINIB, CYCLOPHOSPHAMIDE, CYCLOSPORINE, CYSTEAMINE BITARTRATE, CYTARABINE, DACARBAZINE, DACOMITINIB, DACTINOMYCIN, DARATUMUMAB, DAROLUTAMIDE, DASATINIB, DECITABINE, DENILEUKIN |

| | |
|---|---|
| | DIFTITOX, DENOSUMAB, DEXAMETHASONE, DEXAMETHASONE ACETATE, DEXAMETHASONE SODIUM PHOSPHATE, DIETHYLSTILBESTROL, DINUTUXIMAB, DOCETAXEL, DOLASETRON MESYLATE, DRONABINOL, DURVALUMAB, DUTASTERIDE, DUVELISIB, ELOTUZUMAB, ENASIDENIB MESYLATE, ENCORAFENIB, ENFORTUMAB VEDOTIN-EJFV, ENTRECTINIB, ENZALUTAMIDE, EPOETIN ALFA, EPOETIN ALFA-EPBX, ERDAFITINIB, ERIBULIN MESYLATE, ESTRADIOL, ESTRADIOL VALERATE, ESTRAMUSTINE PHOSPHATE SODIUM, ESTROGENS, CONJUGATED, ETIDRONATE DISODIUM, ETOPOSIDE, ETOPOSIDE PHOSPHATE, EVEROLIMUS, EXEMESTANE, FAM-TRASTUZUMAB DERUXTECAN-NXKI, FILGRASTIM, FILGRASTIM-AAFI, FILGRASTIM-SNDZ, FINASTERIDE, FLOXURIDINE, FLUDARABINE PHOSPHATE, FLUOROURACIL, FLUOXYMESTERONE, FLUTAMIDE, FOSAPREPITANT DIMEGLUMINE, FULVESTRANT, GEFITINIB, GEMCITABINE HCL, GEMTUZUMAB OZOGAMICIN, GLUCARPIDASE, GOSERELIN ACETATE, GRANISETRON, HISTRELIN ACETATE, HYDROCORTISONE, HYDROXYPROGESTERONE CAPROATE, HYDROXYUREA, IBRUTINIB, IDELALISIB, IFOSFAMIDE, IMATINIB MESYLATE, IMIQUIMOD, INOTUZUMAB OZOGAMICIN, IPILIMUMAB, ISOTRETINOIN, IVOSIDENIB, IXABEPILONE, KETOCONAZOLE, LANREOTIDE ACETATE, LENALIDOMIDE, LETROZOLE, LEUCOVORIN CALCIUM, LEUPROLIDE ACETATE, LEVOLEUCOVORIN, LEVOLEUCOVORIN CALCIUM, LOMUSTINE, LORLATINIB, LURBINECTEDIN, LUSPATERCEPT-AAMT, LUTETIUM LU 177 DOTATATE, MEDROXYPROGESTERONE ACETATE, MEGESTROL ACETATE, MELPHALAN, MERCAPTOPURINE, MESNA, METHOTREXATE, METHOTREXATE SODIUM, METHOXSALEN, METHYLPREDNISOLONE, METHYLPREDNISOLONE ACETATE, METHYLPREDNISOLONE SODIUM SUCCINATE, METHYLTESTOSTERONE, MIDOSTAURIN, MIFEPRISTONE, MITOMYCIN, MITOTANE, MOGAMULIZUMAB-KPKC, MOXETUMOMAB PASUDOTOX-TDFK, NABILONE, NECITUMUMAB, NELARABINE, NILUTAMIDE, NIVOLUMAB, OBINUTUZUMAB, OCTREOTIDE ACETATE, OFATUMUMAB, OLAPARIB, OLARATUMAB, OMACETAXINE MEPESUCCINATE, ONDANSETRON, ONDANSETRON HCL, OXALIPLATIN, OXYMETHOLONE, PACLITAXEL, PALBOCICLIB, PALIFERMIN, PAMIDRONATE DISODIUM, PANITUMUMAB, PEGASPARGASE, PEGFILGRASTIM, PEGFILGRASTIM-BMEZ, PEGFILGRASTIM-CBQV, PEGINTERFERON ALFA-2A, PEGINTERFERON ALFA-2B, PEMBROLIZUMAB, PEMETREXED DISODIUM, PEMIGATINIB, PENTOSTATIN, PERTUZUMAB, PLERIXAFOR, PLICAMYCIN, POMALIDOMIDE, PORFIMER SODIUM, PRALATREXATE, PRALSETINIB, PREDNISOLONE, PREDNISOLONE ACETATE, PREDNISOLONE SODIUM PHOSPHATE, PREDNISONE, RAMUCIRUMAB, RASBURICASE, REGORAFENIB, RIPRETINIB, RITUXIMAB, RITUXIMAB-ABBS, RITUXIMAB-PVVR, ROMIDEPSIN, SACITUZUMAB GOVITECAN-HZIY, SAMARIUM SM 153 LEXIDRONAM, SARGRAMOSTIM, SELINEXOR, SELPERCATINIB, SILTUXIMAB, STREPTOZOCIN, SUNITINIB MALATE, TAFASITAMAB-CXIX, TAGRAXOFUSP-ERZS, TALIMOGENE LAHERPAREPVEC, TAMOXIFEN CITRATE, TBO-FILGRASTIM, TEMOZOLOMIDE, TEMSIROLIMUS, TENIPOSIDE, TESTOLACTONE, TESTOSTERONE ENANTHATE, THALIDOMIDE, THIOGUANINE, THIOTEPA, THYROTROPIN ALFA, TISAGENLECLEUCEL, |

| | |
|---|---|
| | TOREMIFENE CITRATE, TRABECTEDIN, TRASTUZUMAB, TRASTUZUMAB-ANNS, TRASTUZUMAB-DKST, TRASTUZUMAB-DTTB, TRASTUZUMAB-PKRB, TRASTUZUMAB-QYYP, TRETINOIN, TRIAMCINOLONE, TRIAMCINOLONE ACETONIDE, TRIAMCINOLONE HEXACETONIDE, TRILOSTANE, TRIMETREXATE GLUCURONATE, TRIPTORELIN PAMOATE, TUCATINIB, URACIL MUSTARD, URIDINE TRIACETATE, VALRUBICIN, VANDETANIB, VEMURAFENIB, VENETOCLAX, VINBLASTINE SULFATE, VINCRISTINE SULFATE, VINORELBINE TARTRATE, VISMODEGIB, VORINOSTAT, ZANUBRUTINIB, ZIV-AFLIBERCEPT, ZOLEDRONIC ACID<br><br>OR<br><br>At least one of the following cancer therapy brand names: A-HYDROCORT, A-METHAPRED, ABIRATERONE ACETATE, ABRAXANE, ABSORICA, ABSORICA LD, ACCUTANE, ACTIMMUNE, ADCETRIS, ADRIAMYCIN, ADRUCIL, AFINITOR, AFINITOR DISPERZ, AGRYLIN, AKYNZEO, ALDARA, ALECENSA, ALIMTA, ALIQOPA, ALKERAN, ALOXI, ALUNBRIG, AMIFOSTINE, AMNESTEEM, ANADROL-50, ANASTROZOLE, ANDROID, ANDROXY, ANZEMET, APREPITANT, ARANESP, ARIMIDEX, ARISTOSPAN, AROMASIN, ARRANON, ARSENIC TRIOXIDE, ARZERRA, ASPARLAS, AVASTIN, AVODART, AYVAKIT, AZACITIDINE, BALVERSA, BAVENCIO, BAYCADRON, BELEODAQ, BELRAPZO, BENDEKA, BESPONSA, BEXAROTENE, BEXXAR, BICALUTAMIDE, BICNU, BLENREP, BLINCYTO, BORTEZOMIB, BOSULIF, BRAFTOVI, BROMOCRIPTINE MESYLATE, BRUKINSA, BUSULFAN, BUSULFEX, CABOMETYX, CALQUENCE, CAMPATH, CAMPTOSAR, CAPECITABINE, CAPRELSA, CARBOPLATIN, CARMUSTINE, CASODEX, CEENU, CERVARIX, CESAMET, CINVANTI, CISPLATIN, CLADRIBINE, CLARAVIS, CLOFARABINE, CLOLAR, COMETRIQ, COPIKTRA, CORTEF, CORTISONE ACETATE, COSMEGEN, COTELLIC, CYCLOPHOSPHAMIDE, CYCLOSPORINE, CYCLOSPORINE MODIFIED, CYRAMZA, CYSTAGON, CYTARABINE, CYTOXAN, DACARBAZINE, DACOGEN, DACTINOMYCIN, DARZALEX, DARZALEX FASPRO, DAUNOXOME, DAURISMO, DECADRON, DECITABINE, DELATESTRYL, DELESTROGEN, DELTASONE, DEPO-MEDROL, DEPO-PROVERA, DEPOCYT, DEXABLISS, DEXAMETHASONE, DEXAMETHASONE INTENSOL, DEXAMETHASONE SODIUM PHOSPHATE, DEXASONE, DEXRAZOXANE, DIDRONEL, DOCEFREZ, DOCETAXEL, DOXIL, DRONABINOL, DUTASTERIDE, DXEVO, EFUDEX, ELIGARD, ELITEK, ELLENCE, ELOXATIN, ELSPAR, ELZONRIS, EMCYT, EMEND, EMPLICITI, ENHERTU, EPOGEN, ERBITUX, ERGAMISOL, ERIVEDGE, ERLEADA, ERWINAZE, ESTRACE, ESTRADIOL, ESTRADIOL VALERATE, ETHYOL, ETOPOPHOS, ETOPOSIDE, EVEROLIMUS, EVISTA, EVOMELA, EXEMESTANE, FARESTON, FARYDAK, FASLODEX, FEMARA, FINASTERIDE, FIRMAGON, FLO-PRED, FLOXURIDINE, FLUDARA, FLUDARABINE PHOSPHATE, FLUOROURACIL, FLUTAMIDE, FOLOTYN, FOSAPREPITANT DIMEGLUMINE, FULPHILA, FULVESTRANT, FUSILEV, GARDASIL, GARDASIL 9, GAVRETO, GAZYVA, GEMZAR, GENGRAF, GILOTRIF, GLEEVEC, GLEOSTINE, GLIADEL, GRANISOL, GRANIX, HALAVEN, HEMADY, HERCEPTIN, HERCEPTIN HYLECTA, HERZUMA, HEXALEN, HICON, HIZENTRA, HYCAMTIN, HYDREA, HYDROCORTISONE, HYDROCORTONE, |

| | |
|---|---|
| | HYDROXYPROGESTERONE CAPROATE, HYDROXYUREA, IBRANCE, ICLUSIG, IDAMYCIN, IDAMYCIN PFS, IDHIFA, IFEX, IFOSFAMIDE, IMATINIB MESYLATE, IMBRUVICA, IMFINZI, IMIQUIMOD, IMLYGIC, INFUGEM, INLYTA, INQOVI, INREBIC, INTRON A, IRESSA, ISOTRETINOIN, ISTODAX, IXEMPRA, JAKAFI, JELMYTO, JEVTANA, KADCYLA, KANJINTI, KENALOG-10, KENALOG-40, KENALOG-80, KEPIVANCE, KEYTRUDA, KHAPZORY, KISQALI, KISQALI FEMARA CO-PACK, KORLYM, KOSELUGO, KYMRIAH, KYPROLIS, KYTRIL, LAPATINIB, LARTRUVO, LEMTRADA, LENVIMA, LETROZOLE, LEUCOVORIN CALCIUM, LEUKERAN, LEUKINE, LEUPROLIDE ACETATE, LEVOLEUCOVORIN CALCIUM, LIBTAYO, LIPODOX 50, LOMUSTINE, LONSURF, LORBRENA, LUMOXITI, LUPRON DEPOT, LUTATHERA, LYNPARZA, LYSODREN, MAKENA, MARINOL, MARQIBO, MATULANE, MAVENCLAD, MEDROL, MEDROXYPROGESTERONE ACETATE, MEGACE, MEGESTROL ACETATE, MEKINIST, MEKTOVI, MELPHALAN, MENEST, MERCAPTOPURINE, MESNA, MESNEX, METASTRON, METHITEST, METHOTREXATE, METHOTREXATE SODIUM, METHYLPRED DP, METHYLPREDNISOLONE, METHYLPREDNISOLONE ACETATE, METHYLTESTOSTERONE, METOCLOPRAMIDE HYDROCHLORIDE, MILLIPRED, MITOMYCIN, MITOSOL, MONJUVI, MOZOBIL, MUSTARGEN, MUTAMYCIN, MVASI, MYLERAN, MYLOTARG, MYORISAN, NAVELBINE, NEORAL, NERLYNX, NEULASTA, NEUPOGEN, NEXAVAR, NILANDRON, NILUTAMIDE, NINLARO, NIPENT, NIVESTYM, NOLVADEX, NUBEQA, OCTREOTIDE ACETATE, ODOMZO, OFEV, OGIVRI, ONCASPAR, ONDANSETRON HCL, ONIVYDE, ONTAK, ONTRUZANT, ONUREG, OPDIVO, ORAPRED, ORAPRED ODT, OXALIPLATIN, PACLITAXEL, PADCEV, PAMIDRONATE DISODIUM, PANRETIN, PARAPLATIN, PARLODEL, PEDIAPRED, PEGASYS, PEMAZYRE, PERJETA, PHESGO, PHOTOFRIN, PIQRAY, PLATINOL-AQ, PLENAXIS, POLIVY, POMALYST, PORTRAZZA, POTELIGEO, PREDNISOLONE, PREDNISOLONE SODIUM PHOSPHATE, PREDNISONE, PREDNISONE INTENSOL, PREMARIN, PROCRIT, PROCYSBI, PROLEUKIN, PROLIA, PROSCAR, PROVENGE, PROVERA, PURINETHOL, PURIXAN, QINLOCK, QUADRAMET, RAYOS, READYSHARP DEXAMETHASONE, READYSHARP TRIAMCINOLONE, REBLOZYL, REGLAN, RETACRIT, RETEVMO, REVLIMID, RITUXAN, RITUXAN HYCELA, ROMIDEPSIN, ROZLYTREK, RUBEX, RUBRACA, RUXIENCE, RYDAPT, SANCUSO, SANDIMMUNE, SANDOSTATIN, SANDOSTATIN LAR, SANDOSTATIN LAR DEPOT, SARCLISA, SODIUM IODIDE I-131, SOLTAMOX, SOLU-CORTEF, SOLU-MEDROL, SOMATULINE DEPOT, SPRYCEL, STIVARGA, SUPPRELIN LA, SUSTOL, SUTENT, SYLATRON, SYLVANT, SYMPROIC, SYNDROS, SYNRIBO, TABLOID, TABRECTA, TAFINLAR, TAGRISSO, TALZENNA, TAMOXIFEN CITRATE, TARCEVA, TARGRETIN, TASIGNA, TAXOL, TAXOTERE, TAZVERIK, TECARTUS, TECENTRIQ, TEMODAR, TEMOZOLOMIDE, TEMSIROLIMUS, TENIPOSIDE, TEPADINA, TESLAC, TESTOSTERONE ENANTHATE, TESTRED, THALOMID, THERACYS, THIOTEPA, THYROGEN, TIBSOVO, TICE BCG, TOPOSAR, TOREMIFENE CITRATE, TORISEL, TOTECT, TRAZIMERA, TREANDA, TRELSTAR, TRETINOIN, TREXALL, TRIAMCINOLONE ACETONIDE, TRISENOX, TRODELVY, TRUXIMA, TUKYSA, TURALIO, TYKERB, UDENYCA, UNITUXIN, UVADEX, VALCHLOR, VALSTAR, VANDETANIB, VANTAS, VARUBI, VECTIBIX, VELCADE, VENCLEXTA, VERIPRED 20, VERZENIO, VESANOID, VIADUR, VIDAZA, VINBLASTINE |

| | |
|---|---|
| | SULFATE, VINCASAR PFS, VINCRISTINE SULFATE, VINORELBINE TARTRATE, VISTOGARD, VITRAKVI, VIZIMPRO, VORAXAZE, VOTRIENT, VYXEOS, XALKORI, XATMEP, XELODA, XGEVA, XOFIGO, XOSPATA, XPOVIO, XTANDI, XURIDEN, YERVOY, YESCARTA, YONDELIS, YONSA, ZALTRAP, ZANOSAR, ZARXIO, ZCORT, ZEJULA, ZELBORAF, ZENATANE, ZEPZELCA, ZEVALIN, ZIEXTENZO, ZINECARD, ZIRABEV, ZODEX, ZOFRAN, ZOFRAN ODT, ZOLADEX, ZOLEDRONIC ACID, ZOLINZA, ZOMETA, ZUPLENZ, ZYDELIG, ZYKADIA, ZYTIGA<br>a. At least one of the following cancer therapy HCPCS or CPT codes: A9545, C9021, C9025, C9058, C9062, C9064, C9117, C9213, C9215, C9235, C9243, C9257, C9259, C9260, C9273, C9284, C9287, C9295, C9297, C9415, C9421, C9423, C9429, C9431, C9432, C9433, C9440, C9455, C9483, J0128, J0640, J0641, J0642, J0885, J0896, J0897, J1030, J1040, J1050, J1051, J1260, J1380, J1436, J1440, J1446, J1453, J1675, J1710, J1720, J1725, J1930, J1950, J2405, J2469, J2502, J2505, J2650, J2783, J3120, J3121, J3305, J7509, J7512, J7527, J8520, J8530, J8540, J8560, J8562, J8600, J8655, J8670, J8700, J8705, J8999, J9001, J9015, J9017, J9019, J9020, J9025, J9030, J9032, J9033, J9043, J9050, J9055, J9060, J9062, J9092, J9093, J9095, J9096, J9098, J9120, J9140, J9151, J9160, J9171, J9176, J9180, J9181, J9185, J9190, J9198, J9200, J9202, J9203, J9204, J9207, J9211, J9225, J9246, J9262, J9263, J9264, J9266, J9268, J9269, J9270, J9290, J9291, J9295, J9301, J9303, J9305, J9307, J9308, J9310, J9320, J9325, J9328, J9330, J9358, J9360, J9370, J9371, J9380, J9390, J9395, Q0162, Q0166, Q0179, Q2025, Q2043, Q2048, Q2050, Q5101, Q5111, Q5113, Q5115, Q5117, Q5119, Q5120, S0108, S0115, S0116, S0119, S0146, S0156, S0174, S0176, S0177, S0178, S0179, S0181, S2107, S9338, A9534, C1086, C9004, C9012, C9027, C9065, C9066, C9110, C9118, C9127, C9131, C9205, C9210, C9214, C9216, C9237, C9239, C9240, C9252, C9253, C9262, C9265, C9276, C9280, C9289, C9292, C9293, C9296, C9414, C9417, C9418, C9420, C9422, C9424, C9425, C9426, C9427, C9428, C9430, C9437, C9474, C9492, J0202, J0207, J0594, J0881, J0894, J1020, J1094, J1100, J1190, J1441, J1442, J1447, J1626, J1627, J1726, J1729, J2353, J2354, J2425, J2562, J2820, J2860, J2920, J2930, J3130, J3240, J3300, J3301, J3315, J3487, J3489, J3590, J7506, J7510, J7684, J8499, J8501, J8510, J8521, J8561, J8565, J8610, J8650, J9000, J9002, J9010, J9027, J9031, J9034, J9035, J9036, J9039, J9040, J9041, J9042, J9045, J9047, J9065, J9070, J9080, J9090, J9091, J9094, J9097, J9100, J9110, J9118, J9130, J9150, J9155, J9170, J9177, J9178, J9179, J9182, J9201, J9206, J9208, J9209, J9213, J9214, J9216, J9217, J9218, J9219, J9226, J9227, J9228, J9230, J9245, J9250, J9260, J9261, J9265, J9267, J9271, J9280, J9285, J9293, J9299, J9300, J9302, J9304, J9306, J9313, J9315, J9340, J9350, J9351, J9354, J9355, J9356, J9357, |

| | |
|---|---|
| | J9375, J9400, J9600, J9999, Q0167, Q0168, Q0180, Q2017, Q2024, Q2049, Q2051, Q5114, Q5118, S0087, S0088, S0091, S0145, S0165, S0170, S0172, S0182, S0187, S0190 |
| **Primary immunodeficiency** | Any prior history of a primary immunodeficiency<br>defined as a medical claim with any of the following ICD-10 diagnosis codes:<br>B20, D82, D82.0, D82.1, D82.3, D82.4, D82.9, B97.35, D45, D46.22, D47.1, D47.4, D47.9, D47.Z1, D47.Z9, D61.82, D75.81,<br>D82.2, D82.8, R75, Z21 |
| **HIV infection** | Any prior history of HIV defined as having as any medical claim with one of the following ICD-10 diagnosis codes:<br>B20, B97.35, R75, Z21<br><br>OR<br><br>The following HCPCS code: 3490F |
| **Immunosuppressive therapy** | A fill or claim for one of the following immunosuppressive therapies in the 60 days before CED identified any medical claim or pharmacy claim with at least one of the following generic names:<br>BELATACEPT, BRODALUMAB, CERTOLIZUMAB PEGOL, CYCLOSPORINE/CHONDROITIN SULFATE A SODIUM, CYTARABINE LIPOSOME/PF, DAUNORUBICIN/CYTARABINE LIPOSOMAL, DECITABINE/CEDAZURIDINE, EFALIZUMAB, EMAPALUMAB-LZSG, INFLIXIMAB-DYYB, NATALIZUMAB, OCRELIZUMAB, PEMETREXED DISODIUM, RAVULIZUMAB-CWVZ, RILONACEPT, TACROLIMUS IN VEHICLE BASE NO.238, TACROLIMUS/NIACINAMIDE, TERIFLUNOMIDE, TILDRAKIZUMAB-ASMN, TOCILIZUMAB, UPADACITINIB, ANAKINRA, ECULIZUMAB, FINGOLIMOD HCL, FLUOROURACIL/ADHESIVE BANDAGE, INEBILIZUMAB-CDON, INFLIXIMAB, INFLIXIMAB-AXXQ, NELARABINE, OZANIMOD HYDROCHLORIDE, SARILUMAB, SATRALIZUMAB-MWGE, SECUKINUMAB, SIPONIMOD, STREPTOZOCIN, TACROLIMUS, MICRONIZED, TACROLIMUS/HYALURONATE SODIUM/NIACINAMIDE, TEPROTUMUMAB-TRBW, VEDOLIZUMAB, ABATACEPT, ABATACEPT/MALTOSE, ALEFACEPT, AZATHIOPRINE SODIUM, BARICITINIB, BASILIXIMAB, CARMUSTINE IN POLIFEPROSAN 20, DIROXIMEL FUMARATE, GUSELKUMAB, INFLIXIMAB-ABDA, PRALATREXATE, SILTUXIMAB, TACROLIMUS ANHYDROUS, USTEKINUMAB, CANAKINUMAB/PF, GOLIMUMAB, MELPHALAN HCL/BETADEX SULFOBUTYL ETHER SODIUM, MUROMONAB-CD3, PIRFENIDONE, RISANKIZUMAB-RZAA, TOFACITINIB CITRATE, ALEMTUZUMAB, CLOFARABINE, GEMCITABINE HCL IN 0.9 % SODIUM CHLORIDE, POMALIDOMIDE, APREMILAST, BENDAMUSTINE HCL, DACLIZUMAB, IXEKIZUMAB, FLOXURIDINE, OFATUMUMAB, IFOSFAMIDE/MESNA, LOMUSTINE, DIMETHYL FUMARATE, MYCOPHENOLATE MOFETIL HCL, BELIMUMAB, CARMUSTINE, MELPHALAN, TEMSIROLIMUS, THIOTEPA, LENALIDOMIDE, THALIDOMIDE, CLADRIBINE, FLUDARABINE PHOSPHATE, ADALIMUMAB, CHLORAMBUCIL, MELPHALAN HCL, DECITABINE, METHOTREXATE, CYTARABINE/PF, ETANERCEPT, |

| | |
|---|---|
| | AZACITIDINE, IFOSFAMIDE, BUSULFAN, LEFLUNOMIDE, DACARBAZINE, MERCAPTOPURINE, MYCOPHENOLATE SODIUM, METHOTREXATE/PF, CYTARABINE, EVEROLIMUS, SIROLIMUS, CYCLOSPORINE, MODIFIED, CAPECITABINE, CYCLOSPORINE<br><br>OR<br><br>At least one of the following CPT/HCPC procedure codes: 80158, 80180, 80197, C9106, C9126, C9211, C9212, C9230, C9236, C9239, C9249, C9261, C9264, C9436, C9438, C9455, J0129, J0215, J1300, J1438, J1602, J1628, J2350, J3357, J7501, J7502, J7504, J7507, J7508, J7513, J7517, J7518, J7525, J7527, J8561, J8610, J9070, J9080, J9090, J9092, J9093, J9096, J9250, J9260, J9311, J9312, K0119, K0122, K0412, Q5103, S0087, S0162, S9359, 80169, 80195, C9006, C9020, C9026, C9110, C9219, C9286, C9419, C9420, C9421, J0135, J0202, J0480, J0485, J0490, J0638, J0717, J0718, J1745, J2323, J2793, J2860, J3245, J3262, J3358, J3380, J7500, J7503, J7505, J7511, J7515, J7520, J8530, J9010, J9065, J9091, J9094, J9095, J9097, J9310, J9330, K0120, K0121, K0123, Q2019, Q2044, Q4079, Q5104, Q5109, S0193 |

CED, coding entry date; DRG, diagnosis-related group; Dx, diagnosis; HCPCS, healthcare common procedure coding system; ICD-10, *International Statistical Classification of Diseases and Related Health Problems-10th Revision*; NDC, national drug code.

**Table S4. Claims-based algorithms for covariates of interest**

| Covariate | Algorithm |
|---|---|
| **Age** | Patient characteristic assessed on date of cohort entry |
| **Sex** | Patient characteristic assessed on date of cohort entry |
| **Payer type** | Patient characteristic assessed on date of cohort entry |
| **State of residence** | Patient characteristic assessed on date of cohort entry |
| **Level of COVID-19 transmission by state** | Case counts were defined based on data reported for the previous 7 days on the last day of each month per state based on county population data from the US. Census rolled up to the state level. Transmission data from the CDC were used to calculate the state-level incidence proportion by dividing the state-level case count on the last day of each month by the state-level population and multiplying by 100,000. Transmission level was assigned based on the CDC's transmission levels of low, moderate, substantial, and high transmission (grouping low and moderate together into one category in our analysis).<br><br>County-level transmission data sourced from: https://data.cdc.gov/Public-Health-Surveillance/United-States-COVID-19-County-Level-of-Community-T/nra9-vzzn<br><br>County-level population data sourced from:<br><br>https://www.census.gov/data/datasets/time-series/demo/popest/2010s-counties-total.html#par_textimage_70769902 |
| **Calendar time** | NA |
| **Place of service** | Patient characteristics assessed on date of cohort entry<br>● Defined as categorization of: pharmacy: pharmacy, \1<br>● Non-medical Setting: school, homeless shelter, temporary lodging, mobile unit, 02<br>● Outpatient/clinic/doctor's office: 72, 58, 19, urgent care facility, walk-in retail health clinic, state or local public health clinic, end-stage renal disease treatment facility, comprehensive outpatient rehabilitation facility, community mental health center, federally qualified health center, independent clinic, 25, ambulatory surgical center, outpatient hospital, office, 26<br>● Inpatient/hospital/ER: inpatient hospital, emergency room, inpatient psychiatric facility<br>● Ambulance: ambulance - air or water, ambulance - land<br>● Mass immunization center: mass immunization center<br>● Long term facility: hospice, nursing facility, skilled nursing facility, psychiatric facility partial hospitalization, intermediate care facility for mentally retarded, residential substance abuse treatment facility, comprehensive inpatient rehabilitation facility, psychiatric residential treatment facility<br>● Other: independent laboratory, other unlisted facility, non-residential substance abuse treatment facility, 28, 29, 30, 37, 38, 43, 59, 73, 76, 79, 82, 83, 85, 86, 92, 95, \N, 0\, 00, |

| | |
|---|---|
| | 0E, 0H, 0I, 0J, 0k, 0L, 0N, 0O, 0X, 10, 18, 2`, AH, AT, CG, G0, HA, HO, M7, NU, NV, O2, OF, UN<br><br>https://www.cms.gov/Medicare/Coding/place-of-service-codes/Place_of_Service_Code_Set |
| **Healthcare resource utilization** | NA |
| **Charlson Quan comorbidity score** | Coding Algorithms for Defining Comorbidities in<br>ICD-9-CM and ICD-10 Administrative Data<br>(Quan et al. 2005) |
| **Frailty** | Use Frailty index score from FDA ML<br>https://www.ncbi.nlm.nih.gov/pmc/articles/PMC6001883/pdf/glx229.pdf<br>https://www.ncbi.nlm.nih.gov/pmc/articles/PMC6001883/bin/glx229_suppl_data-supplement.docx |
| **Alcohol use** | ICD-10-CM: F10.1, F10.10, F10.11, F10.12, F10.120, F10.121, F10.129, F10.13, F10.130, F10.131, F10.132, F10.139, F10.14, F10.15, F10.150, F10.151, F10.159, F10.18, F10.180, F10.181, F10.182, F10.188, F10.19, F10.2, F10.20, F10.21, F10.22, F10.220, F10.221, F10.229, F10.23, F10.230, F10.231, F10.232, F10.239, F10.24, F10.25, F10.250, F10.251, F10.259, F10.26, F10.27, F10.28, F10.280, F10.281, F10.282, F10.288, F10.29, F10.9, F10.92, F10.920, F10.921, F10.929, F10.93, F10.930, F10.931, F10.932, F10.939, F10.94, F10.95, F10.950, F10.951, F10.959, F10.96, F10.97, F10.98, F10.980, F10.981, F10.982, F10.988, F10.99, E52, F10, G62.1, I42.6, K70.0, K29.2, K70.3, K70.9, Z71.4, T51, T51.0, T51.0X, T51.0X1, T51.0X1A, T51.0X1D, T51.0X1S, T51.0X2, T51.0X2A, T51.0X2D, T51.0X2S, T51.0X3, T51.0X3A, T51.0X3D, T51.0X3S, T51.0X4, T51.0X4A, T51.0X4D, T51.0X4S, T51.1, T51.1X, T51.1X1, T51.1X1A, T51.1X1D, T51.1X1S, T51.1X2, T51.1X2A, T51.1X2D, T51.1X2S, T51.1X3, T51.1X3A, T51.1X3D, T51.1X3S, T51.1X4, T51.1X4A, T51.1X4D, T51.1X4S, T51.2, T51.2X, T51.2X1, T51.2X1A, T51.2X1D, T51.2X1S, T51.2X2, T51.2X2A, T51.2X2D, T51.2X2S, T51.2X3, T51.2X3A, T51.2X3D, T51.2X3S, T51.2X4, T51.2X4A, T51.2X4D, T51.2X4S, T51.3, T51.3X, T51.3X1, T51.3X1A, T51.3X1D, T51.3X1S, T51.3X2, T51.3X2A, T51.3X2D, T51.3X2S, T51.3X3, T51.3X3A, T51.3X3D, T51.3X3S, T51.3X4, T51.3X4A, T51.3X4D, T51.3X4S, T51.8, T51.8X, T51.8X1, T51.8X1A, T51.8X1D, T51.8X1S, T51.8X2, T51.8X2A, T51.8X2D, T51.8X2S, T51.8X3, T51.8X3A, T51.8X3D, T51.8X3S, T51.8X4, T51.8X4A, T51.8X4D, T51.8X4S, T51.9, T51.91, T51.91XA, T51.91XD, T51.91XS, T51.92, T51.92XA, T51.92XD, T51.92XS, T51.93, T51.93XA, T51.93XD, T51.93XS, T51.94, T51.94XA, T51.94XD, T51.94XS |
| **Arrhythmia** | ICD-10-CM: I47, I47.0, I48, I48.2, I48.9, I48.91, I49.0, I49.01, I49.3, I49.4, I49.49, I49.8, I49.9, R00, I47.1, I47.2, I47.9, I48.0, I48.1, I48.11, I48.19, I48.20, I48.21, I48.3, I48.4, I48.92, I49, I49.02, I49.1, I49.2, I49.40, I49.5, R00.0, R00.1, R00.2, R00.8, R00.9<br>https://www.sentinelinitiative.org/methods-data-tools/methods/master-protocol-development-covid-19-natural-history Coding Algorithms for Defining Comorbidities in Natural History Code List. |
| **Asthma** | ICD-10-CM: J45.20, J45.21, J45.22, J45.30, J45.31, J45.32, J45.40, J45.41, J45.42, J45.50, J45.51, J45.52, J45.901, J45.902, J45.909, J45.990, J45.991, J45.998, J45, J45.2, J45.3, J45.4, J45.5, J45.9, J45.90, J45.99<br>https://www.sentinelinitiative.org/methods-data-tools/methods/masterprotocol-development-covid-19-natural-history Coding Algorithms for Defining Comorbidities in Natural History Code List. |
| **Cancer** | ICD-10-CM: C00, C00.0, C00.1, C00.2, C00.3, C00.4, C00.5, C00.6, C00.8, C00.9, C01, C02, C02.0, C02.1, C02.2, C02.3, C02.4, C02.8, C02.9, C03, C03.0, C03.1, C03.9, C04, C04.0, C04.1, C04.8, |

| | |
|---|---|
| | C04.9, C05, C05.0, C05.1, C05.2, C05.8, C05.9, C06, C06.0, C06.1, C06.2, C06.8, C06.80, C06.89, C06.9, C07, C08, C08.0, C08.1, C08.9, C09, C09.0, C09.1, C09.8, C09.9, C10, C10.0, C10.1, C10.2, C10.3, C10.4, C10.8, C10.9, C11, C11.0, C11.1, C11.2, C11.3, C11.8, C11.9, C12, C13, C13.0, C13.1, C13.2, C13.8, C13.9, C14, C14.0, C14.2, C14.8, C15, C15.3, C15.4, C15.5, C15.8, C15.9, C16, C16.0, C16.1, C16.2, C16.3, C16.4, C16.5, C16.6, C16.8, C16.9, C17, C17.0, C17.1, C17.2, C17.3, C17.8, C17.9, C18, C18.0, C18.1, C18.2, C18.3, C18.4, C18.5, C18.6, C18.7, C18.8, C18.9, C19, C20, C21, C21.0, C21.1, C21.2, C21.8, C22, C22.0, C22.1, C22.2, C22.3, C22.4, C22.7, C22.8, C22.9, C23, C24, C24.0, C24.1, C24.8, C24.9, C25, C25.0, C25.1, C25.2, C25.3, C25.4, C25.7, C25.8, C25.9, C26, C26.0, C26.1, C26.9, C30, C30.0, C30.1, C31, C31.0, C31.1, C31.2, C31.3, C31.8, C31.9, C32, C32.0, C32.1, C32.2, C32.3, C32.8, C32.9, C33, C34, C34.0, C34.00, C34.01, C34.02, C34.1, C34.10, C34.11, C34.12, C34.2, C34.3, C34.30, C34.31, C34.32, C34.8, C34.80, C34.81, C34.82, C34.9, C34.90, C34.91, C34.92, C37, C38, C38.0, C38.1, C38.2, C38.3, C38.4, C38.8, C39, C39.0, C39.9, C40, C40.0, C40.00, C40.01, C40.02, C40.1, C40.10, C40.11, C40.12, C40.2, C40.20, C40.21, C40.22, C40.3, C40.30, C40.31, C40.32, C40.8, C40.80, C40.81, C40.82, C40.9, C40.90, C40.91, C40.92, C41, C41.0, C41.1, C41.2, C41.3, C41.4, C41.9, C43, C43.0, C43.1, C43.10, C43.11, C43.111, C43.112, C43.12, C43.121, C43.122, C43.2, C43.20, C43.21, C43.22, C43.3, C43.30, C43.31, C43.39, C43.4, C43.5, C43.51, C43.52, C43.59, C43.6, C43.60, C43.61, C43.62, C43.7, C43.70, C43.71, C43.72, C43.8, C43.9, C45, C45.0, C45.1, C45.2, C45.7, C45.9, C46, C46.0, C46.1, C46.2, C46.3, C46.4, C46.5, C46.50, C46.51, C46.52, C46.7, C46.9, C47, C47.0, C47.1, C47.10, C47.11, C47.12, C47.2, C47.20, C47.21, C47.22, C47.3, C47.4, C47.5, C47.6, C47.8, C47.9, C48, C48.0, C48.1, C48.2, C48.8, C49, C49.0, C49.1, C49.10, C49.11, C49.12, C49.2, C49.20, C49.21, C49.22, C49.3, C49.4, C49.5, C49.6, C49.8, C49.9, C49.A, C49.A0, C49.A1, C49.A2, C49.A3, C49.A4, C49.A5, C49.A9, C50, C50.0, C50.01, C50.011, C50.012, C50.019, C50.02, C50.021, C50.022, C50.029, C50.1, C50.11, C50.111, C50.112, C50.119, C50.12, C50.121, C50.122, C50.129, C50.2, C50.21, C50.211, C50.212, C50.219, C50.22, C50.221, C50.222, C50.229, C50.3, C50.31, C50.311, C50.312, C50.319, C50.32, C50.321, C50.322, C50.329, C50.4, C50.41, C50.411, C50.412, C50.419, C50.42, C50.421, C50.422, C50.429, C50.5, C50.51, C50.511, C50.512, C50.519, C50.52, C50.521, C50.522, C50.529, C50.6, C50.61, C50.611, C50.612, C50.619, C50.62, C50.621, C50.622, C50.629, C50.8, C50.81, C50.811, C50.812, C50.819, C50.82, C50.821, C50.822, C50.829, C50.9, C50.91, C50.911, C50.912, C50.919, C50.92, C50.921, C50.922, C50.929, C51, C51.0, C51.1, C51.2, C51.8, C51.9, C52, C53, C53.0, C53.1, C53.8, C53.9, C54, C54.0, C54.1, C54.2, C54.3, C54.8, C54.9, C55, C56, C56.1, C56.2, C56.9, C57, C57.0, C57.00, C57.01, C57.02, C57.1, C57.10, C57.11, C57.12, C57.2, C57.20, C57.21, C57.22, C57.3, C57.4, C57.7, C57.8, C57.9, C58, C60, C60.0, C60.1, C60.2, C60.8, C60.9, C61, C62, C62.0, C62.00, C62.01, C62.02, C62.1, C62.10, C62.11, C62.12, C62.9, C62.90, C62.91, C62.92, C63, C63.0, C63.00, C63.01, C63.02, C63.1, C63.10, C63.11, C63.12, C63.2, C63.7, C63.8, C63.9, C64, C64.1, C64.2, C64.9, C65, C65.1, C65.2, C65.9, C66, C66.1, C66.2, C66.9, C67, C67.0, C67.1, C67.2, C67.3, C67.4, C67.5, C67.6, C67.7, C67.8, C67.9, C68, C68.0, C68.1, C68.8, C68.9, C69, C69.0, C69.00, C69.01, C69.02, C69.1, C69.10, C69.11, C69.12, C69.2, C69.20, C69.21, C69.22, C69.3, C69.30, C69.31, C69.32, C69.4, C69.40, C69.41, C69.42, C69.5, C69.50, C69.51, C69.52, C69.6, C69.60, C69.61, C69.62, C69.8, C69.80, C69.81, C69.82, C69.9, C69.90, C69.91, C69.92, C70, C70.0, C70.1, C70.9, C71, C71.0, C71.1, C71.2, C71.3, C71.4, C71.5, C71.6, C71.7, C71.8, C71.9, C72, C72.0, C72.1, C72.2, C72.20, C72.21, C72.22, C72.3, C72.30, C72.31, C72.32, C72.4, C72.40, C72.41, C72.42, C72.5, C72.50, C72.59, C72.9, C73, C74, C74.0, C74.00, C74.01, C74.02, C74.1, C74.10, C74.11, C74.12, C74.9, C74.90, C74.91, C74.92, C75, C75.0, C75.1, C75.2, C75.3, C75.4, C75.5, C75.8, C75.9, C76, C76.0, C76.1, C76.2, C76.3, C76.4, C76.40, C76.41, C76.42, C76.5, C76.50, C76.51, C76.52, C76.8, C7A, C7A.0, C7A.00, C7A.01, C7A.010, C7A.011, C7A.012, C7A.019, C7A.02, C7A.020, C7A.021, C7A.022, C7A.023, C7A.024, C7A.025, C7A.026, C7A.029, |

| | |
|---|---|
| | C7A.09, C7A.090, C7A.091, C7A.092, C7A.093, C7A.094, C7A.095, C7A.096, C7A.098, C7A.1, C7A.8, C7B, C7B.0, C7B.00, C7B.01, C7B.02, C7B.03, C7B.04, C7B.09, C7B.1, C7B.8, C81, C81.0, C81.00, C81.01, C81.02, C81.03, C81.04, C81.05, C81.06, C81.07, C81.08, C81.09, C81.1, C81.10, C81.11, C81.12, C81.13, C81.14, C81.15, C81.16, C81.17, C81.18, C81.19, C81.2, C81.20, C81.21, C81.22, C81.23, C81.24, C81.25, C81.26, C81.27, C81.28, C81.29, C81.3, C81.30, C81.31, C81.32, C81.33, C81.34, C81.35, C81.36, C81.37, C81.38, C81.39, C81.4, C81.40, C81.41, C81.42, C81.43, C81.44, C81.45, C81.46, C81.47, C81.48, C81.49, C81.7, C81.70, C81.71, C81.72, C81.73, C81.74, C81.75, C81.76, C81.77, C81.78, C81.79, C81.9, C81.90, C81.91, C81.92, C81.93, C81.94, C81.95, C81.96, C81.97, C81.98, C81.99, C82, C82.0, C82.00, C82.01, C82.02, C82.03, C82.04, C82.05, C82.06, C82.07, C82.08, C82.09, C82.1, C82.10, C82.11, C82.12, C82.13, C82.14, C82.15, C82.16, C82.17, C82.18, C82.19, C82.2, C82.20, C82.21, C82.22, C82.23, C82.24, C82.25, C82.26, C82.27, C82.28, C82.29, C82.3, C82.30, C82.31, C82.32, C82.33, C82.34, C82.35, C82.36, C82.37, C82.38, C82.39, C82.4, C82.40, C82.41, C82.42, C82.43, C82.44, C82.45, C82.46, C82.47, C82.48, C82.49, C82.5, C82.50, C82.51, C82.52, C82.53, C82.54, C82.55, C82.56, C82.57, C82.58, C82.59, C82.6, C82.60, C82.61, C82.62, C82.63, C82.64, C82.65, C82.66, C82.67, C82.68, C82.69, C82.8, C82.80, C82.81, C82.82, C82.83, C82.84, C82.85, C82.86, C82.87, C82.88, C82.89, C82.9, C82.90, C82.91, C82.92, C82.93, C82.94, C82.95, C82.96, C82.97, C82.98, C82.99, C83, C83.0, C83.00, C83.01, C83.02, C83.03, C83.04, C83.05, C83.06, C83.07, C83.08, C83.09, C83.1, C83.10, C83.11, C83.12, C83.13, C83.14, C83.15, C83.16, C83.17, C83.18, C83.19, C83.3, C83.30, C83.31, C83.32, C83.33, C83.34, C83.35, C83.36, C83.37, C83.38, C83.39, C83.5, C83.50, C83.51, C83.52, C83.53, C83.54, C83.55, C83.56, C83.57, C83.58, C83.59, C83.7, C83.70, C83.71, C83.72, C83.73, C83.74, C83.75, C83.76, C83.77, C83.78, C83.79, C83.8, C83.80, C83.81, C83.82, C83.83, C83.84, C83.85, C83.86, C83.87, C83.88, C83.89, C83.9, C83.90, C83.91, C83.92, C83.93, C83.94, C83.95, C83.96, C83.97, C83.98, C83.99, C84, C84.0, C84.00, C84.01, C84.02, C84.03, C84.04, C84.05, C84.06, C84.07, C84.08, C84.09, C84.1, C84.10, C84.11, C84.12, C84.13, C84.14, C84.15, C84.16, C84.17, C84.18, C84.19, C84.4, C84.40, C84.41, C84.42, C84.43, C84.44, C84.45, C84.46, C84.47, C84.48, C84.49, C84.6, C84.60, C84.61, C84.62, C84.63, C84.64, C84.65, C84.66, C84.67, C84.68, C84.69, C84.7, C84.70, C84.71, C84.72, C84.73, C84.74, C84.75, C84.76, C84.77, C84.78, C84.79, C84.9, C84.90, C84.91, C84.92, C84.93, C84.94, C84.95, C84.96, C84.97, C84.98, C84.99, C84.A, C84.A0, C84.A1, C84.A2, C84.A3, C84.A4, C84.A5, C84.A6, C84.A7, C84.A8, C84.A9, C84.Z, C84.Z0, C84.Z1, C84.Z2, C84.Z3, C84.Z4, C84.Z5, C84.Z6, C84.Z7, C84.Z8, C84.Z9, C85, C85.1, C85.10, C85.11, C85.12, C85.13, C85.14, C85.15, C85.16, C85.17, C85.18, C85.19, C85.2, C85.20, C85.21, C85.22, C85.23, C85.24, C85.25, C85.26, C85.27, C85.28, C85.29, C85.8, C85.80, C85.81, C85.82, C85.83, C85.84, C85.85, C85.86, C85.87, C85.88, C85.89, C85.9, C85.90, C85.91, C85.92, C85.93, C85.94, C85.95, C85.96, C85.97, C85.98, C85.99, C88, C88.0, C88.2, C88.3, C88.4, C88.8, C88.9, C90, C90.0, C90.00, C90.01, C90.02, C90.1, C90.10, C90.11, C90.12, C90.2, C90.20, C90.21, C90.22, C90.3, C90.30, C90.31, C90.32, C91, C91.0, C91.00, C91.01, C91.02, C91.1, C91.10, C91.11, C91.12, C91.3, C91.30, C91.31, C91.32, C91.4, C91.40, C91.41, C91.42, C91.5, C91.50, C91.51, C91.52, C91.6, C91.60, C91.61, C91.62, C91.9, C91.90, C91.91, C91.92, C91.A, C91.A0, C91.A1, C91.A2, C91.Z, C91.Z0, C91.Z1, C91.Z2, C92, C92.0, C92.00, C92.01, C92.02, C92.1, C92.10, C92.11, C92.12, C92.2, C92.20, C92.21, C92.22, C92.3, C92.30, C92.31, C92.32, C92.4, C92.40, C92.41, C92.42, C92.5, C92.50, C92.51, C92.52, C92.6, C92.60, C92.61, C92.62, C92.9, C92.90, C92.91, C92.92, C92.A, C92.A0, C92.A1, C92.A2, C92.Z, C92.Z0, C92.Z1, C92.Z2, C93, C93.0, C93.00, C93.01, C93.02, C93.1, C93.10, C93.11, C93.12, C93.3, C93.30, C93.31, C93.32, C93.9, C93.90, C93.91, C93.92, C93.Z, C93.Z0, C93.Z1, C93.Z2, C94, C94.0, C94.00, C94.01, C94.02, C94.2, C94.20, C94.21, C94.22, C94.3, C94.30, C94.31, C94.32, C94.4, C94.40, C94.41, C94.42, C94.6, C94.8, C94.80, C94.81, C94.82, C95, C95.0, C95.00, C95.01, C95.02, C95.1, C95.10, C95.11, C95.12, C95.9, C95.90, C95.91, C95.92, C96, C96.0, C96.2, C96.20, C96.21, C96.22, C96.29, |

| | |
|---|---|
| | C96.4, C96.5, C96.6, C96.9, C96.A, C96.Z, C77, C77.0, C77.1, C77.2, C77.3, C77.4, C77.5, C77.8, C77.9, C78, C78.0, C78.00, C78.01, C78.02, C78.1, C78.2, C78.3, C78.30, C78.39, C78.4, C78.5, C78.6, C78.7, C78.8, C78.80, C78.89, C79, C79.0, C79.00, C79.01, C79.02, C79.1, C79.10, C79.11, C79.19, C79.2, C79.3, C79.31, C79.32, C79.4, C79.40, C79.49, C79.5, C79.51, C79.52, C79.6, C79.60, C79.61, C79.62, C79.7, C79.70, C79.71, C79.72, C79.8, C79.81, C79.82, C79.89, C79.9, C80, C80.0, C80.1, C80.2<br>https://www.sentinelinitiative.org/methods-data-tools/methods/masterprotocol-development-covid-19-natural-history Coding Algorithms for Defining Comorbidities in Natural History Code List. |
| **Cardiovascular disease** | ICD-10-CM: I01, I01.0, I01.1, I01.2, I01.8, I01.9, I02.0, I05, I05.0, I05.1, I05.2, I05.8, I05.9, I06, I06.0, I06.1, I06.2, I06.8, I06.9, I07, I07.0, I07.1, I07.2, I07.8, I07.9, I08, I08.0, I08.1, I08.2, I08.3, I08.8, I08.9, I09, I09.0, I09.1, I09.2, I09.8, I09.81, I09.89, I09.9, I10, I11, I11.0, I11.9, I12, I12.0, I12.9, I13, I13.0, I13.1, I13.10, I13.11, I13.2, I15, I15.0, I15.1, I15.2, I15.8, I15.9, I16, I16.0, I16.1, I16.9, I20, I20.0, I20.1, I20.8, I20.9, I21, I21.0, I21.01, I21.02, I21.09, I21.1, I21.11, I21.19, I21.2, I21.21, I21.29, I21.3, I21.4, I21.9, I21.A, I21.A1, I21.A9, I22, I22.0, I22.1, I22.2, I22.8, I22.9, I23, I23.0, I23.1, I23.2, I23.3, I23.4, I23.5, I23.6, I23.7, I23.8, I24, I24.0, I24.1, I24.8, I24.9, I25, I25.1, I25.10, I25.11, I25.110, I25.111, I25.118, I25.119, I25.2, I25.3, I25.4, I25.41, I25.42, I25.5, I25.6, I25.7, I25.70, I25.700, I25.701, I25.708, I25.709, I25.71, I25.710, I25.711, I25.718, I25.719, I25.72, I25.720, I25.721, I25.728, I25.729, I25.73, I25.730, I25.731, I25.738, I25.739, I25.75, I25.750, I25.751, I25.758, I25.759, I25.76, I25.760, I25.761, I25.768, I25.769, I25.79, I25.790, I25.791, I25.798, I25.799, I25.8, I25.81, I25.810, I25.811, I25.812, I25.82, I25.83, I25.84, I25.89, I25.9, I26, I26.0, I26.01, I26.02, I26.09, I26.9, I26.90, I26.92, I26.93, I26.94, I26.99, I27, I27.0, I27.1, I27.2, I27.20, I27.21, I27.22, I27.23, I27.24, I27.29, I27.8, I27.81, I27.82, I27.83, I27.89, I27.9, I28, I28.0, I28.1, I28.8, I28.9, I30, I30.0, I30.1, I30.8, I30.9, I31, I31.0, I31.1, I31.2, I31.3, I31.4, I31.8, I31.9, I32, I33, I33.0, I33.9, I34, I34.0, I34.1, I34.2, I34.8, I34.9, I35, I35.0, I35.1, I35.2, I35.8, I35.9, I36, I36.0, I36.1, I36.2, I36.8, I36.9, I37, I37.0, I37.1, I37.2, I37.8, I37.9, I38, I39, I40, I40.0, I40.1, I40.8, I40.9, I41, I42, I42.0, I42.1, I42.2, I42.3, I42.4, I42.5, I42.6, I42.7, I42.8, I42.9, I43, I44, I44.0, I44.1, I44.2, I44.3, I44.30, I44.39, I44.4, I44.5, I44.6, I44.60, I44.69, I44.7, I45, I45.0, I45.1, I45.10, I45.19, I45.2, I45.3, I45.4, I45.5, I45.6, I45.8, I45.81, I45.89, I45.9, I46, I46.2, I46.8, I46.9, I47, I47.0, I47.1, I47.2, I47.9, I48, I48.0, I48.1, I48.11, I48.19, I48.2, I48.20, I48.21, I48.3, I48.4, I48.9, I48.91, I48.92, I49, I49.0, I49.01, I49.02, I49.1, I49.2, I49.3, I49.4, I49.40, I49.49, I49.5, I49.8, I49.9, I50, I50.1, I50.2, I50.20, I50.21, I50.22, I50.23, I50.3, I50.30, I50.31, I50.32, I50.33, I50.4, I50.40, I50.41, I50.42, I50.43, I50.8, I50.81, I50.810, I50.811, I50.812, I50.813, I50.814, I50.82, I50.83, I50.84, I50.89, I50.9, I51, I51.0, I51.1, I51.2, I51.3, I51.4, I51.5, I51.7, I51.8, I51.81, I51.89, I51.9, I52, I70, I70.0, I70.1, I70.2, I70.20, I70.201, I70.202, I70.203, I70.208, I70.209, I70.21, I70.211, I70.212, I70.213, I70.218, I70.219, I70.22, I70.221, I70.222, I70.223, I70.228, I70.229, I70.23, I70.231, I70.232, I70.233, I70.234, I70.235, I70.238, I70.239, I70.24, I70.241, I70.242, I70.243, I70.244, I70.245, I70.248, I70.249, I70.25, I70.26, I70.261, I70.262, I70.263, I70.268, I70.269, I70.29, I70.291, I70.292, I70.293, I70.298, I70.299, I70.3, I70.30, I70.301, I70.302, I70.303, I70.308, I70.309, I70.31, I70.311, I70.312, I70.313, I70.318, I70.319, I70.32, I70.321, I70.322, I70.323, I70.328, I70.329, I70.33, I70.331, I70.332, I70.333, I70.334, I70.335, I70.338, I70.339, I70.34, I70.341, I70.342, I70.343, I70.344, I70.345, I70.348, I70.349, I70.35, I70.36, I70.361, I70.362, I70.363, I70.368, I70.369, I70.39, I70.391, I70.392, I70.393, I70.398, I70.399, I70.4, I70.40, I70.401, I70.402, I70.403, I70.408, I70.409, I70.41, I70.411, I70.412, I70.413, I70.418, I70.419, I70.42, I70.421, I70.422, I70.423, I70.428, I70.429, I70.43, I70.431, I70.432, I70.433, I70.434, I70.435, I70.438, I70.439, I70.44, I70.441, I70.442, I70.443, I70.444, I70.445, I70.448, I70.449, I70.45, I70.46, I70.461, I70.462, I70.463, I70.468, I70.469, I70.49, I70.491, I70.492, I70.493, I70.498, I70.499, I70.5, I70.50, I70.501, |

| | |
|---|---|
| | I70.502, I70.503, I70.508, I70.509, I70.51, I70.511, I70.512, I70.513, I70.518, I70.519, I70.52, I70.521, I70.522, I70.523, I70.528, I70.529, I70.53, I70.531, I70.532, I70.533, I70.534, I70.535, I70.538, I70.539, I70.54, I70.541, I70.542, I70.543, I70.544, I70.545, I70.548, I70.549, I70.55, I70.56, I70.561, I70.562, I70.563, I70.568, I70.569, I70.59, I70.591, I70.592, I70.593, I70.598, I70.599, I70.6, I70.60, I70.601, I70.602, I70.603, I70.608, I70.609, I70.61, I70.611, I70.612, I70.613, I70.618, I70.619, I70.62, I70.621, I70.622, I70.623, I70.628, I70.629, I70.63, I70.631, I70.632, I70.633, I70.634, I70.635, I70.638, I70.639, I70.64, I70.641, I70.642, I70.643, I70.644, I70.645, I70.648, I70.649, I70.65, I70.66, I70.661, I70.662, I70.663, I70.668, I70.669, I70.69, I70.691, I70.692, I70.693, I70.698, I70.699, I70.7, I70.70, I70.701, I70.702, I70.703, I70.708, I70.709, I70.71, I70.711, I70.712, I70.713, I70.718, I70.719, I70.72, I70.721, I70.722, I70.723, I70.728, I70.729, I70.73, I70.731, I70.732, I70.733, I70.734, I70.735, I70.738, I70.739, I70.74, I70.741, I70.742, I70.743, I70.744, I70.745, I70.748, I70.749, I70.75, I70.76, I70.761, I70.762, I70.763, I70.768, I70.769, I70.79, I70.791, I70.792, I70.793, I70.798, I70.799, I70.8, I70.9, I70.90, I70.91, I70.92, I71, I71.0, I71.00, I71.01, I71.02, I71.03, I71.1, I71.2, I71.3, I71.4, I71.5, I71.6, I71.8, I71.9, I72, I72.0, I72.1, I72.2, I72.3, I72.4, I72.5, I72.6, I72.8, I72.9, I73, I73.0, I73.00, I73.01, I73.1, I73.8, I73.81, I73.89, I73.9, I74, I74.0, I74.01, I74.09, I74.1, I74.10, I74.11, I74.19, I74.2, I74.3, I74.4, I74.5, I74.8, I74.9, I75, I75.0, I75.01, I75.011, I75.012, I75.013, I75.019, I75.02, I75.021, I75.022, I75.023, I75.029, I75.8, I75.81, I75.89, I76, I77, I77.0, I77.1, I77.2, I77.3, I77.4, I77.5, I77.6, I77.7, I77.70, I77.71, I77.72, I77.73, I77.74, I77.75, I77.76, I77.77, I77.79, I77.8, I77.81, I77.810, I77.811, I77.812, I77.819, I77.89, I77.9, I78, I78.0, I78.1, I78.8, I78.9, I79, I79.0, I79.1, I79.8, I80, I80.0, I80.00, I80.01, I80.02, I80.03, I80.1, I80.10, I80.11, I80.12, I80.13, I80.2, I80.20, I80.201, I80.202, I80.203, I80.209, I80.21, I80.211, I80.212, I80.213, I80.219, I80.22, I80.221, I80.222, I80.223, I80.229, I80.23, I80.231, I80.232, I80.233, I80.239, I80.24, I80.241, I80.242, I80.243, I80.249, I80.25, I80.251, I80.252, I80.253, I80.259, I80.29, I80.291, I80.292, I80.293, I80.299, I80.3, I80.8, I80.9, I81, I82, I82.0, I82.1, I82.2, I82.21, I82.210, I82.211, I82.22, I82.220, I82.221, I82.29, I82.290, I82.291, I82.3, I82.4, I82.40, I82.401, I82.402, I82.403, I82.409, I82.41, I82.411, I82.412, I82.413, I82.419, I82.42, I82.421, I82.422, I82.423, I82.429, I82.43, I82.431, I82.432, I82.433, I82.439, I82.44, I82.441, I82.442, I82.443, I82.449, I82.45, I82.451, I82.452, I82.453, I82.459, I82.46, I82.461, I82.462, I82.463, I82.469, I82.49, I82.491, I82.492, I82.493, I82.499, I82.4Y, I82.4Y1, I82.4Y2, I82.4Y3, I82.4Y9, I82.4Z, I82.4Z1, I82.4Z2, I82.4Z3, I82.4Z9, I82.5, I82.50, I82.501, I82.502, I82.503, I82.509, I82.51, I82.511, I82.512, I82.513, I82.519, I82.52, I82.521, I82.522, I82.523, I82.529, I82.53, I82.531, I82.532, I82.533, I82.539, I82.54, I82.541, I82.542, I82.543, I82.549, I82.55, I82.551, I82.552, I82.553, I82.559, I82.56, I82.561, I82.562, I82.563, I82.569, I82.59, I82.591, I82.592, I82.593, I82.599, I82.5Y, I82.5Y1, I82.5Y2, I82.5Y3, I82.5Y9, I82.5Z, I82.5Z1, I82.5Z2, I82.5Z3, I82.5Z9, I82.6, I82.60, I82.601, I82.602, I82.603, I82.609, I82.61, I82.611, I82.612, I82.613, I82.619, I82.62, I82.621, I82.622, I82.623, I82.629, I82.7, I82.70, I82.701, I82.702, I82.703, I82.709, I82.71, I82.711, I82.712, I82.713, I82.719, I82.72, I82.721, I82.722, I82.723, I82.729, I82.8, I82.81, I82.811, I82.812, I82.813, I82.819, I82.89, I82.890, I82.891, I82.9, I82.90, I82.91, I82.A, I82.A1, I82.A11, I82.A12, I82.A13, I82.A19, I82.A2, I82.A21, I82.A22, I82.A23, I82.A29, I82.B, I82.B1, I82.B11, I82.B12, I82.B13, I82.B19, I82.B2, I82.B21, I82.B22, I82.B23, I82.B29, I82.C, I82.C1, I82.C11, I82.C12, I82.C13, I82.C19, I82.C2, I82.C21, I82.C22, I82.C23, I82.C29, I83, I83.0, I83.00, I83.001, I83.002, I83.003, I83.004, I83.005, I83.008, I83.009, I83.01, I83.011, I83.012, I83.013, I83.014, I83.015, I83.018, I83.019, I83.02, I83.021, I83.022, I83.023, I83.024, I83.025, I83.028, I83.029, I83.1, I83.10, I83.11, I83.12, I83.2, I83.20, I83.201, I83.202, I83.203, I83.204, I83.205, I83.208, I83.209, I83.21, I83.211, I83.212, I83.213, I83.214, I83.215, I83.218, I83.219, I83.22, I83.221, I83.222, I83.223, I83.224, I83.225, I83.228, I83.229, I83.8, I83.81, I83.811, I83.812, I83.813, I83.819, I83.89, I83.891, I83.892, I83.893, I83.899, I83.9, I83.90, I83.91, I83.92, I83.93, I85, I85.0, I85.00, |

| | |
|---|---|
| | I85.01, I85.1, I85.10, I85.11, I86, I86.0, I86.1, I86.2, I86.3, I86.4, I86.8, I87, I87.0, I87.00, I87.001, I87.002, I87.003, I87.009, I87.01, I87.011, I87.012, I87.013, I87.019, I87.02, I87.021, I87.022, I87.023, I87.029, I87.03, I87.031, I87.032, I87.033, I87.039, I87.09, I87.091, I87.092, I87.093, I87.099, I87.1, I87.2, I87.3, I87.30, I87.301, I87.302, I87.303, I87.309, I87.31, I87.311, I87.312, I87.313, I87.319, I87.32, I87.321, I87.322, I87.323, I87.329, I87.33, I87.331, I87.332, I87.333, I87.339, I87.39, I87.391, I87.392, I87.393, I87.399, I87.8, I87.9, I88, I88.0, I88.1, I88.8, I88.9, I89, I89.0, I89.1, I89.8, I89.9, I95, I95.0, I95.1, I95.2, I95.3, I95.8, I95.81, I95.89, I95.9, I96, I97, I97.0, I97.1, I97.11, I97.110, I97.111, I97.12, I97.120, I97.121, I97.13, I97.130, I97.131, I97.19, I97.190, I97.191, I97.2, I97.3, I97.4, I97.41, I97.410, I97.411, I97.418, I97.42, I97.5, I97.51, I97.52, I97.6, I97.61, I97.610, I97.611, I97.618, I97.62, I97.620, I97.621, I97.622, I97.63, I97.630, I97.631, I97.638, I97.64, I97.640, I97.641, I97.648, I97.7, I97.71, I97.710, I97.711, I97.79, I97.790, I97.791, I97.8, I97.81, I97.810, I97.811, I97.82, I97.820, I97.821, I97.88, I97.89, I99, I99.8, I99.9<br>https://www.sentinelinitiative.org/methods-data-tools/methods/masterprotocol-development-covid-19-natural-history Coding Algorithms for<br>Defining Comorbidities in Natural History Code List. |
| **Cerebrovascular disease** | ICD-10: G45.x, G46.x, H34.0, I60.x–I69.x<br>Coding Algorithms for Defining Comorbidities in ICD-9-CM and ICD-10 Administrative Data (Quan et al. 2005) Coding Algorithms for Defining Comorbidities in Charlson and Elixhauser comorbidities |
| **Coronary artery disease** | ICD-10: I21, I21.01, I21.09, I21.2, I21.29, I21.3, I21.4, I22.1, I24.0, I24.9, I25.110, I25.119, I25.8, I25.83, I25.84, I25.89, I25.9, I21.02, I21.21, I21.A9, I22, I22.2, I22.8, I25.118, I25.81, I25.811, I21.0, I21.1, I22.9, I24, I25.1, I25.82, I21.19, I21.9, I21.A1, I24.8, I25.11, I25.810, I25.812, I20.0, I21.11, I22.0, I24.1, I25.10, I25.111, I25.2<br>FDA ML in the COVID Instance:<br>https://covid.aetion.com/measures/details/16999/0/0/basics<br>https://covid.aetion.com/measures/details/16999/0/0/basics |
| **Chronic kidney disease** | ICD-10-CM: I12.0, I13.1, N03.2*-N03.7*, N05.2*– N05.7*, N17*, N18*, N19*, N25.0, Z49.0*–Z49.2*, Z94.0*, Z99.2<br>Coding Algorithms for Defining Comorbidities in ICD-9-CM and ICD-10 Administrative Data (Quan et al. 2005) Coding Algorithms for Defining Comorbidities inICD-9-CM and ICD-10 Administrative Data (Quan et al. 2005) |
| **Chronic lung disease** | ICD-10-CM: I27.8*, I27.9*, J40*–J47*, J60*–J67*, J68.4, J70.1, J70.3<br>Coding Algorithms for Defining Comorbidities in ICD-9-CM and ICD-10 Administrative Data (Quan et al. 2005) |
| **Chronic obstructive pulmonary disease** | ICD-10-CM: I27.8, I27.9, J40.x–J47.x, J60.x–J67.x, J68.4, J70.1, J70.3<br>https://czresearch.com/dropbox/Quan_MedCare_2005v43p1130.pdf |
| **Dementia** | ICD-10: F00.x–F03.x, G30.x, G31.1, F05.1<br>https://czresearch.com/dropbox/Quan_MedCare_2005v43p1130.pdf |
| **Diabetes (type I and type II)** | ICD-10-CM: E10.0, E10.1, E10.6, E10.8, E10.9, E11.0, E11.1, E11.6, E11.8, E11.9, E12.0, E12.1, E12.6, E12.8, E12.9, E13.0, E13.1, E13.6, E13.8, E13.9, E14.0, E14.1, E14.6, E14.8, E14.9, E10.2–E10.5, E10.7, E11.2–E11.5, E11.7, E12.2–E12.5, E12.7, E13.2–E13.5, E13.7, E14.2–E14.5, E14.7<br>https://czresearch.com/dropbox/Quan_MedCare_2005v43p1130.pdf |
| **Down syndrome** | ICD-10-CM: Q90.x<br>https://www.ncbi.nlm.nih.gov/pmc/articles/PMC5958386/pdf/10.1177_0033354918763168.pdf |
| **Heart failure** | ICD-10: I09.9, I11.0, I13.0, I13.2, I25.5, I42.0, I42.5–I42.9, I43.x, I50.x, P29.0<br>Coding Algorithms for Defining Comorbidities in ICD-9-CM and ICD-10 Administrative Data (Quan et al. 2005) |

| | |
|---|---|
| **Hypertension** | ICD-10: I10.x, I11.x–I13.x, I15.x<br>Coding Algorithms for Defining Comorbidities in ICD-9-CM and ICD-10 Administrative Data (Quan et al. 2005) |
| **Irritable bowel syndrome** | ICD-10-CM: K25, K26, K27, K28, K29, K30, K31.0, K31.8, K31.9, R11, K31.1 - K31.7, K35, K38.0, K38.8, K55, K56, K57, K72, K70, K73.9, K73.8, K74.5, K76.0, K76.9, K75.0, K75.1, K72.9, K76.6, K76.7, K76.8, K76.1, K77.0, K75.9, K76.3, K76.8, K76.9, K80, K81.0, K81.8, K82, K91.5, K83, K85, K86.1, K86.2, K86.8, K86.9, K90, K91.2, R11, R12, R13, R14, R15, N23, N39.4, R30, R32, R33, R34, R35, R36, R39, R10, R16, R18, R19, N80, N94, F34.1, F40.1, F40.2, F41.0, F41.1, F41.8, F41.9, F42.8, F44.8, F44.9, F45.2, A04, A08, A02, A09<br>Lix LM, Yogendran MS, Shaw SY, Targownick LE, Jones J, Bataineh O. Comparing administrative and survey data for ascertaining cases of irritable bowel syndrome: a population-based investigation. *BMC Health Serv Res.* 2010;10:31. |
| **Obesity** | ICD-10: E66.x<br>Coding Algorithms for Defining Comorbidities in ICD-9-CM and ICD-10 Administrative Data (Quan et al. 2005) |
| **Other respiratory viruses (e.g. influenza, RSV)** | ICD-10-CM: J09, J10.0, J10.1, J10.8 (influenza), J12.1, J20.5, J21.0, B97.4 (RSV)<br>https://journals.plos.org/plosone/article?id=10.1371/journal.pone.0244746 |
| **Liver disease** | ICD-10-CM: B18.x, K70.0–K70.3, K70.9, K71.3–K71.5, K71.7, K73.x, K74.x, K76.0, K76.2–K76.4, K76.8, K76.9, Z94.4, I85.0, I86.4, K70.4, K71.1, K72.1, K72.9, K76.5, K76.6, K76.7, I85.9, I98.2<br>https://czresearch.com/dropbox/Quan_MedCare_2005v43p1130.pdf |
| **Psoriasis** | ICD-10-CM: L40.0<br>Schneeweiss M, Merola JF, Karlson EW, Solomon DH. Rationale and design of the Brigham Cohort for psoriasis and psoriatic arthritis registry (COPPAR). *BMC Dermatology* (2017);17(1):11. |
| **Psoriatic arthritis** | ICD-10-CM: L40.5<br>Schneeweiss M, Merola JF, Karlson EW, Solomon DH. Rationale and Design of the Brigham Cohort for psoriasis and psoriatic arthritis registry (COPPAR). *BMC Dermatology* (2017);17(1):11. |
| **Rheumatoid arthritis** | ICD-10-CM: M05.00, M05.30, M05.60, M06.1, M06.9, M08.00, M08.3, M08.40<br>Jafri K, Taylor L, Nezamzadeh M, Baker JF, Mehta NN, Bartels C, Williams CT, Ogdie A. Management of hyperlipidemia among patients with rheumatoid arthritis in the primary care setting. BMC Musculoskelet Disord. 2015;16:237. doi: 10.1186/s12891-015-0700-5. PubMed PMID: 26336889; PubMed Central PMCID: PMC4559905. |
| **Sickle cell disease or thalassemia** | ICD-10-CM: D57, D57.0, D57.00, D57.01, D57.02, D57.03, D57.09, D57.1, D57.2, D57.20, D57.21, D57.211, D57.212, D57.213, D57.218, D57.219, D57.3, D57.4, D57.40, D57.41, D57.411, D57.412, D57.413, D57.418, D57.419, D57.42, D57.43, D57.431, D57.432, D57.433, D57.438, D57.439, D57.44, D57.45, D57.451, D57.452, D57.453, D57.458, D57.459<br>Aetion modification based on: https://assets.milliman.com/ektron/A_claims-based_analysis_of_sickle_cell_disease_Prevalence_disease_complications_and_costs.pdf |
| **Tobacco use** | ICD-10-CM: F17.21*, F17.22*, F17.29*, Z71.6, Z72.0, Z87.891, 099.33*, T65.21*<br>CPT: 99406, 99407, G0436, G0437, G9016, S9453, S4995, G9276, G9458, 1034F, 4004F, 4001F<br>NDC Generic Names: *nicotine*, *varenicline*<br>Definition based in-part on Desai et al, 2016 and CMS General Equivalence Mappings. |
| **Number of unique immunosuppressive therapies** | Immunosuppressive therapies listed in **Table S2** |

a Drug class included topical agent, MTOR inhibitor, inosine monophosphate, anti-inflammatory, calcineurin inhibitor, multiple sclerosis agent, T-lymphocyte immunoglobulin, JAK inhibitor, antileprotics (thalidomide), antifibrotic agent, IL-1 inhibitor/IL-1 receptor agonist, IGF-R inhibitor, Fusion protein, immunosuppressive antimetabolite, antineoplastic agent, monoclonal antibody

**Table S5. Baseline variables hypothesized to be confounding variables included in a propensity score (PS) model**

| | Pre-weighted | | Post PS-weighted | | |
|---|---|---|---|---|---|
| | mRNA-1273 | BNT162b2 | mRNA-1273 | BNT162b2 | Weighted absolute standardized difference |
| **Number of patients, n** | | | 57,700 | 66,757 | |
| Sum of weights | - | - | 124,587.74 | 124,312.52 | |
| Effective sample size | - | - | 55,605 | 64,940 | |
| **Age (continuous)** | | | | | 0.009 |
| Mean age, y (SD) | 51.71 (13.42) | 50.18 (13.49) | 50.93 (13.44) | 50.81 (13.47) | |
| **Age, y (categorical), n (%)** | | | | | |
| 18-29 | 4,260 (7.4%) | 6,140 (9.2%) | 10,409.5 (8.4%) | 10,404.7 (8.4%) | 0.001 |
| 30-39 | 7,032 (12.2%) | 8,926 (13.4%) | 16,006.5 (12.9%) | 15,981.7 (12.9%) | 0.000 |
| 40=49 | 10,247 (17.8%) | 12,408 (18.6%) | 22,680.0 (18.2%) | 22,641.8 (18.2%) | 0.000 |
| 50=64 | 29,938 (51.9%) | 33,935 (50.8%) | 63,969.0 (51.3%) | 63,870.8 (51.4%) | 0.001 |
| 65=74 | 4,528 (7.8%) | 4,041 (6.1%) | 8,542.3 (6.9%) | 8,466.3 (6.8%) | 0.002 |
| ≥75 | 1,695 (2.9%) | 1,307 (2.0%) | 2,980.4 (2.4%) | 2,947.1 (2.4%) | 0.001 |
| **Sex, n (%)** | | | | | |
| Female | 30,006 (52.0%) | 35,475 (53.1%) | 65,568.3 (52.6%) | 65,440.1 (52.6%) | 0.000 |
| Male | 27,694 (48.0%) | 31,282 (46.9%) | 59,019.4 (47.4%) | 58,872.4 (47.4%) | 0.000 |
| **Primary payer type, n (%)** | | | | | |
| Commercial | 33,954 (58.8%) | 43,273 (64.8%) | 77,545.3 (62.2%) | 77,445.3 (62.3%) | 0.001 |
| Medicare | 3,134 (5.4%) | 1,824 (2.7%) | 4,919.7 (4.0%) | 4,831.0 (3.9%) | 0.003 |
| Medicaid | 19,856 (34.4%) | 20,402 (30.6%) | 40,094.5 (32.2%) | 40,014.1 (32.2%) | 0.000 |
| Missing | 756 (1.3%) | 1,258 (1.9%) | 2,028.2 (1.6%) | 2,022.1 (1.6%) | 0.000 |
| **State, n (%)** | | | | | |
| Alaska/Other/Puerto Rice | 29 (0.1%) | 10 (0.0%) | 38.2 (0.0%) | 31.9 (0.0%) | 0.003 |
| Alabama | 42 (0.1%) | 57 (0.1%) | 99.9 (0.1%) | 99.1 (0.1%) | 0.000 |
| Arkansas | 371 (0.6%) | 422 (0.6%) | 833.1 (0.7%) | 824.4 (0.7%) | 0.001 |

| | | | | | |
|---|---|---|---|---|---|
| Arizona | 1,493 (2.6%) | 1,297 (1.9%) | 2,763.9 (2.2%) | 2,758.8 (2.2%) | 0.000 |
| California | 6,264 (10.9%) | 4,843 (7.3%) | 11,003.5 (8.8%) | 10,867.5 (8.7%) | 0.003 |
| Colorado | 364 (0.6%) | 470 (0.7%) | 844.1 (0.7%) | 837.8 (0.7%) | 0.000 |
| Connecticut | 49 (0.1%) | 94 (0.1%) | 146.6 (0.1%) | 143.5 (0.1%) | 0.001 |
| Washington, DC | 194 (0.3%) | 151 (0.2%) | 338.4 (0.3%) | 334.3 (0.3%) | 0.001 |
| Delaware | 192 (0.3%) | 180 (0.3%) | 374.7 (0.3%) | 376.7 (0.3%) | 0.000 |
| Florida | 2,954 (5.1%) | 3,372 (5.1%) | 6,291.6 (5.1%) | 6,298.6 (5.1%) | 0.001 |
| Georgia | 1,504 (2.6%) | 1,480 (2.2%) | 2,974.1 (2.4%) | 2,959.4 (2.4%) | 0.000 |
| Hawaii | 77 (0.1%) | 76 (0.1%) | 155.1 (0.1%) | 153.2 (0.1%) | 0.000 |
| Iowa | 372 (0.6%) | 500 (0.7%) | 867.5 (0.7%) | 868.7 (0.7%) | 0.000 |
| Idaho/Wyoming | 34 (0.1%) | 49 (0.1%) | 82.9 (0.1%) | 83.2 (0.1%) | 0.000 |
| Illinois | 6,678 (11.6%) | 9,152 (13.7%) | 15,838.2 (12.7%) | 15,862.6 (12.8%) | 0.001 |
| Indiana | 795 (1.4%) | 1,433 (2.1%) | 2,238.6 (1.8%) | 2,230.7 (1.8%) | 0.000 |
| Kansas | 268 (0.5%) | 527 (0.8%) | 806.0 (0.7%) | 799.2 (0.6%) | 0.001 |
| Kentucky | 915 (1.6%) | 813 (1.2%) | 1,743.3 (1.4%) | 1,751.7 (1.4%) | 0.001 |
| Louisiana | 1,497 (2.6%) | 1,264 (1.9%) | 2,739.1 (2.2%) | 2,731.8 (2.2%) | 0.000 |
| Massachusetts/Rhode Island | 280 (0.5%) | 351 (0.5%) | 630.7 (0.5%) | 632.1 (0.5%) | 0.000 |
| Maryland | 482 (0.8%) | 431 (0.6%) | 913.0 (0.7%) | 913.9 (0.7%) | 0.000 |
| Maine | 27 (0.0%) | 38 (0.1%) | 64.0 (0.1%) | 64.6 (0.1%) | 0.000 |
| Michigan | 3,225 (5.6%) | 3,655 (5.5%) | 6,902.1 (5.5%) | 6,908.9 (5.6%) | 0.001 |
| Minnesota | 83 (0.1%) | 154 (0.2%) | 233.6 (0.2%) | 235.8 (0.2%) | 0.001 |
| Missouri | 418 (0.7%) | 626 (0.9%) | 1,052.4 (0.8%) | 1,047.7 (0.8%) | 0.000 |
| Mississippi | 238 (0.4%) | 212 (0.3%) | 445.0 (0.4%) | 445.8 (0.4%) | 0.000 |
| Montana | 154 (0.3%) | 149 (0.2%) | 307.6 (0.3%) | 307.2 (0.3%) | 0.000 |
| North/South Dakota | 28 (0.0%) | 38 (0.1%) | 66.2 (0.1%) | 66.2 (0.1%) | 0.000 |
| North Carolina | 479 (0.8%) | 592 (0.9%) | 1,080.1 (0.9%) | 1,075.5 (0.9%) | 0.000 |
| Nebraska | 95 (0.2%) | 66 (0.1%) | 158.3 (0.1%) | 158.0 (0.1%) | 0.000 |
| New Hampshire | 62 (0.1%) | 106 (0.2%) | 165.8 (0.1%) | 167.7 (0.1%) | 0.001 |
| New Jersey | 4,425 (7.7%) | 5,789 (8.7%) | 10,222.5 (8.2%) | 10,211.4 (8.2%) | 0.000 |
| New Mexico | 820 (1.4%) | 517 (0.8%) | 1,336.4 (1.1%) | 1,338.8 (1.1%) | 0.000 |
| Nevada | 411 (0.7%) | 584 (0.9%) | 987.6 (0.8%) | 991.2 (0.8%) | 0.001 |
| New York | 2,828 (4.9%) | 2,578 (3.9%) | 5,370.0 (4.3%) | 5,354.7 (4.3%) | 0.000 |

| | | | | | |
|---|---|---|---|---|---|
| Ohio | 1,056 (1.8%) | 1,477 (2.2%) | 2,574.1 (2.1%) | 2,559.0 (2.1%) | 0.001 |
| Oklahoma | 1,268 (2.2%) | 1,455 (2.2%) | 2,749.2 (2.2%) | 2,746.7 (2.2%) | 0.000 |
| Oregon | 179 (0.3%) | 206 (0.3%) | 372.6 (0.3%) | 371.3 (0.3%) | 0.000 |
| Pennsylvania | 3,380 (5.9%) | 3,020 (4.5%) | 6,380.9 (5.1%) | 6,381.8 (5.1%) | 0.001 |
| South Carolina | 604 (1.0%) | 643 (1.0%) | 1,255.7 (1.0%) | 1,251.5 (1.0%) | 0.000 |
| Tennessee | 404 (0.7%) | 455 (0.7%) | 854.9 (0.7%) | 855.0 (0.7%) | 0.000 |
| Texas | 8,316 (14.4%) | 11,922 (17.9%) | 20,381.8 (16.4%) | 20,301.6 (16.3%) | 0.001 |
| Utah | 44 (0.1%) | 83 (0.1%) | 125.0 (0.1%) | 126.4 (0.1%) | 0.000 |
| Virginia | 1,474 (2.6%) | 1,967 (2.9%) | 3,462.5 (2.8%) | 3,457.9 (2.8%) | 0.000 |
| Vermont | 60 (0.1%) | 90 (0.1%) | 149.9 (0.1%) | 149.8 (0.1%) | 0.000 |
| Washington | 936 (1.6%) | 992 (1.5%) | 1,923.9 (1.5%) | 1,927.1 (1.6%) | 0.001 |
| Wisconsin | 1,555 (2.7%) | 2,118 (3.2%) | 3,713.7 (3.0%) | 3,721.8 (3.0%) | 0.001 |
| West Virginia | 277 (0.5%) | 253 (0.4%) | 529.4 (0.4%) | 529.8 (0.4%) | 0.000 |
| **Month and year of cohort entry, n (%)** | | | | | |
| January 2021 | 962 (1.7%) | 1,506 (2.3%) | 2,545.1 (2.0%) | 2,515.6 (2.0%) | 0.001 |
| February 2021 | 6,188 (10.7%) | 5,761 (8.6%) | 12,014.9 (9.6%) | 12,009.3 (9.7%) | 0.001 |
| March 2021 | 10,587 (18.3%) | 12,146 (18.2%) | 22,815.3 (18.3%) | 22,693.6 (18.3%) | 0.002 |
| April 2021 | 20,073 (34.8%) | 23,287 (34.9%) | 43,364.0 (34.8%) | 43,320.2 (34.8%) | 0.001 |
| May 2021 | 10,752 (18.6%) | 10,685 (16.0%) | 21,328.6 (17.1%) | 21,284.4 (17.1%) | 0.000 |
| June 2021 | 4,644 (8.0%) | 5,255 (7.9%) | 9,883.2 (7.9%) | 9,876.5 (7.9%) | 0.000 |
| July 2021 | 1,978 (3.4%) | 3,228 (4.8%) | 5,186.6 (4.2%) | 5,199.5 (4.2%) | 0.001 |
| August 2021 | 1,983 (3.4%) | 4,050 (6.1%) | 6,072.2 (4.9%) | 6,040.6 (4.9%) | 0.001 |
| September 2021 | 533 (0.9%) | 839 (1.3%) | 1,377.9 (1.1%) | 1,372.8 (1.1%) | 0.000 |
| **Clinical conditions, n )%)** | | | | | |
| Alcohol use | 3,644 (6.3%) | 3,694 (5.5%) | 7,352.0 (5.9%) | 7,325.6 (5.9%) | 0.000 |
| Tobacco use/smoking | 18,488 (32.0%) | 19,702 (29.5%) | 38,237.7 (30.7%) | 38,124.7 (30.7%) | 0.001 |
| Obesity (BMI ≥30 kg/m$^2$) | 18,986 (32.9%) | 21,279 (31.9%) | 40,295.5 (32.3%) | 40,181.1 (32.3%) | 0.000 |
| Arrhythmia | 14,515 (25.2%) | 16,883 (25.3%) | 31,395.3 (25.2%) | 31,300.3 (25.2%) | 0.001 |
| Asthma | 9,298 (16.1%) | 10,153 (15.2%) | 19,475.5 (15.6%) | 19,422.9 (15.6%) | 0.000 |
| Cancer | 12,348 (21.4%) | 14,390 (21.6%) | 26,793.0 (21.5%) | 26,721.6 (21.5%) | 0.000 |
| Cardiovascular disease | 37,586 (65.1%) | 41,533 (62.2%) | 79,186.1 (63.6%) | 78,953.0 (63.5%) | 0.001 |
| Cerebrovascular disease | 5,000 (8.7%) | 5,382 (8.1%) | 10,422.2 (8.4%) | 10,365.4 (8.3%) | 0.001 |

| | | | | | |
|---|---|---|---|---|---|
| Chronic kidney disease | 9,230 (16.0%) | 10,046 (15.0%) | 19,277.4 (15.5%) | 19,205.6 (15.5%) | 0.001 |
| Chronic lung disease | 13,871 (24.0%) | 14,476 (21.7%) | 28,370.0 (22.8%) | 28,300.3 (22.8%) | 0.000 |
| Chronic obstructive pulmonary disease | 13,871 (24.0%) | 14,476 (21.7%) | 28,370.0 (22.8%) | 28,300.3 (22.8%) | 0.000 |
| Coronary artery disease | 7,268 (12.6%) | 7,566 (11.3%) | 14,851.2 (11.9%) | 14,796.4 (11.9%) | 0.001 |
| Dementia | 672 (1.2%) | 699 (1.0%) | 1,352.2 (1.1%) | 1,341.8 (1.1%) | 0.001 |
| Diabetes (type I or II) | 13,476 (23.4%) | 14,013 (21.0%) | 27,524.3 (22.1%) | 27,400.5 (22.0%) | 0.001 |
| Down syndrome | 49 (0.1%) | 70 (0.1%) | 116.6 (0.1%) | 117.6 (0.1%) | 0.000 |
| Heart failure | 4,347 (7.5%) | 4,493 (6.7%) | 8,822.6 (7.1%) | 8,773.1 (7.1%) | 0.001 |
| Hypertension | 31,149 (54.0%) | 33,526 (50.2%) | 64,728.4 (51.9%) | 64,515.5 (51.9%) | 0.001 |
| Liver disease | 9,950 (17.2%) | 10,780 (16.1%) | 20,740.4 (16.7%) | 20,656.9 (16.6%) | 0.001 |
| Other respiratory viruses (eg, influenza, RSV) | 1,493 (2.6%) | 1,835 (2.7%) | 3,352.7 (2.7%) | 3,346.3 (2.7%) | 0.000 |
| Sickle cell disease or thalassemia | 162 (0.3%) | 244 (0.4%) | 413.0 (0.3%) | 408.8 (0.3%) | 0.001 |
| Psoriasis | 4,346 (7.5%) | 5,245 (7.9%) | 9,612.0 (7.7%) | 9,602.4 (7.7%) | 0.000 |
| Psoriatic arthritis | 3,918 (6.8%) | 4,547 (6.8%) | 8,484.0 (6.8%) | 8,475.4 (6.8%) | 0.000 |
| Rheumatoid arthritis | 8,484 (14.7%) | 9,245 (13.8%) | 17,750.8 (14.2%) | 17,725.5 (14.3%) | 0.000 |
| Irritable bowel syndrome | 27,241 (47.2%) | 30,717 (46.0%) | 58,056.2 (46.6%) | 57,866.0 (46.6%) | 0.001 |

BMI, body mass index; PS, propensity score; RSV, respiratory syncytial virus.

**Table S6. Distribution of inverse probability of treatment weights.**

| | Weights | | |
|---|---|---|---|
| **Percentile** | **Overall** | **mRNA-1273** | **BNT162b2** |
| Min | 1.16 | 1.17 | 1.16 |
| 5% | 1.50 | 1.59 | 1.46 |
| 10% | 1.59 | 1.69 | 1.54 |
| 20% | 1.68 | 1.80 | 1.63 |
| 25% | 1.72 | 1.85 | 1.66 |
| 30% | 1.75 | 1.91 | 1.68 |
| 40% | 1.83 | 2.02 | 1.74 |
| 50% | 1.92 | 2.12 | 1.80 |
| 60% | 2.03 | 2.22 | 1.87 |
| 70% | 2.15 | 2.34 | 1.96 |
| 75% | 2.22 | 2.41 | 2.01 |
| 80% | 2.30 | 2.47 | 2.08 |
| 90% | 2.51 | 2.65 | 2.26 |
| 95% | 2.69 | 2.87 | 2.41 |
| Max | 6.91 | 6.91 | 5.00 |

**Table S7. Distribution of baseline variables between the index date and the start of follow-up**

| | mRNA-1273 | BNT162b2 | Weighted absolute standardized difference |
|---|---|---|---|
| **Number of patients** | 57,700 | 66,757 | |
| **Sum of weights** | 124,589.33 | 124,311.71 | |
| **Age, y (continuous)** | | | 0.008 |
| **Mean (SD)** | 50.92 (13.44) | 50.81 (13.48) | |
| **Age, y (categorical), n (%)** | | | |
| **18-29** | 10,415.6 (8.4%) | 10,407.6 (8.4%) | 0.000 |
| **30-39y** | 16,004.3 (12.9%) | 15,977.5 (12.9%) | 0.000 |
| **40-49y** | 22,678.6 (18.2%) | 22,643.9 (18.2%) | 0.000 |
| **50-64y** | 63,972.9 (51.3%) | 63,874.4 (51.4%) | 0.001 |
| **65-74y** | 8,540.5 (6.9%) | 8,461.4 (6.8%) | 0.002 |
| **≥75y** | 2,977.4 (2.4%) | 2,946.9 (2.4%) | 0.001 |
| **Sex, n (%)** | | | |
| **Female** | 65,574.5 (52.6%) | 65,437.3 (52.6%) | 0.000 |
| **Male** | 59,014.8 (47.4%) | 58,874.4 (47.4%) | 0.000 |
| **Primary payer type** | | | |
| **Missing** | 2,024.7 (1.6%) | 2,020.3 (1.6%) | 0.000 |
| **Commercial** | 77,546.1 (62.2%) | 77,437.9 (62.3%) | 0.001 |
| **Medicare** | 4,920.6 (4.0%) | 4,828.6 (3.9%) | 0.003 |
| **Medicaid** | 40,098.0 (32.2%) | 40,024.9 (32.2%) | 0.000 |

| | | | |
|---|---|---|---|
| **Place of service categorization, n (%)** | | | |
| **Pharmacy** | 0.0 (0.0%) | 0.0 (0.0%) | - |
| **Non-medical setting** | 476.2 (0.4%) | 456.8 (0.4%) | 0.002 |
| **Outpatient/clinic/doctor's office** | 6,490.7 (5.2%) | 6,459.2 (5.2%) | 0.001 |
| **Inpatient/hospital/ED** | 463.1 (0.4%) | 472.6 (0.4%) | 0.001 |
| **Ambulance** | 27.5 (0.0%) | 20.2 (0.0%) | 0.004 |
| **Mass immunization center** | 0.0 (0.0%) | 1.7 (0.0%) | 0.005 |
| **Long-term facility** | 48.8 (0.0%) | 80.7 (0.1%) | 0.011 |
| **Other** | 117,083.0 (94.0%) | 116,820.7 (94.0%) | 0.000 |
| **State, n (%)** | | | |
| **Alaska/Other/Puerto Rico** | 38.0 (0.0%) | 30.9 (0.0%) | 0.003 |
| **Alabama** | 99.9 (0.1%) | 99.2 (0.1%) | 0.000 |
| **Arkansas** | 834.3 (0.7%) | 824.8 (0.7%) | 0.001 |
| **Arizona** | 2,762.8 (2.2%) | 2,754.5 (2.2%) | 0.000 |
| **California** | 11,003.2 (8.8%) | 10,864.3 (8.7%) | 0.003 |
| **Colorado** | 843.3 (0.7%) | 837.9 (0.7%) | 0.000 |
| **Connecticut** | 146.1 (0.1%) | 143.4 (0.1%) | 0.001 |
| **Washington, DC** | 338.5 (0.3%) | 334.8 (0.3%) | 0.000 |
| **Delaware** | 375.0 (0.3%) | 377.5 (0.3%) | 0.001 |
| **Florida** | 6,290.1 (5.1%) | 6,296.8 (5.1%) | 0.001 |
| **Georgia** | 2,976.6 (2.4%) | 2,962.7 (2.4%) | 0.000 |
| **Hawaii** | 155.5 (0.1%) | 153.6 (0.1%) | 0.000 |

| | | | |
|---|---|---|---|
| **Iowa** | 867.3 (0.7%) | 868.7 (0.7%) | 0.000 |
| **Idaho/Wyoming** | 82.9 (0.1%) | 83.5 (0.1%) | 0.000 |
| **Illinois** | 15,843.0 (12.7%) | 15,867.0 (12.8%) | 0.001 |
| **Indiana** | 2,244.3 (1.8%) | 2,231.5 (1.8%) | 0.001 |
| **Kansas** | 803.7 (0.7%) | 799.0 (0.6%) | 0.000 |
| **Kentucky** | 1,744.4 (1.4%) | 1,753.9 (1.4%) | 0.001 |
| **Louisiana** | 2,737.2 (2.2%) | 2,728.1 (2.2%) | 0.000 |
| **Massachusetts/Rhode Island** | 629.0 (0.5%) | 630.5 (0.5%) | 0.000 |
| **Maryland** | 914.2 (0.7%) | 915.4 (0.7%) | 0.000 |
| **Maine** | 63.8 (0.1%) | 64.5 (0.1%) | 0.000 |
| **Michigan** | 6,905.8 (5.5%) | 6,911.3 (5.6%) | 0.001 |
| **Minnesota** | 232.3 (0.2%) | 235.4 (0.2%) | 0.001 |
| **Missouri** | 1,052.6 (0.8%) | 1,048.1 (0.8%) | 0.000 |
| **Mississippi** | 444.9 (0.4%) | 444.7 (0.4%) | 0.000 |
| **Montana** | 307.0 (0.3%) | 307.2 (0.3%) | 0.000 |
| **North/South Dakota** | 66.0 (0.1%) | 66.1 (0.1%) | 0.000 |
| **North Carolina** | 1,079.8 (0.9%) | 1,075.7 (0.9%) | 0.000 |
| **Nebraska** | 158.1 (0.1%) | 157.7 (0.1%) | 0.000 |
| **New Hampshire** | 166.5 (0.1%) | 168.1 (0.1%) | 0.000 |
| **New Jersey** | 10,228.2 (8.2%) | 10,214.8 (8.2%) | 0.000 |
| **New Mexico** | 1,336.0 (1.1%) | 1,339.4 (1.1%) | 0.001 |
| **Nevada** | 989.9 (0.8%) | 992.8 (0.8%) | 0.001 |

| | | | |
|---|---|---|---|
| **New York** | 5,375.4 (4.3%) | 5,359.5 (4.3%) | 0.000 |
| **Ohio** | 2,575.7 (2.1%) | 2,562.1 (2.1%) | 0.000 |
| **Oklahoma** | 2,744.9 (2.2%) | 2,744.0 (2.2%) | 0.000 |
| **Oregon** | 372.5 (0.3%) | 371.3 (0.3%) | 0.000 |
| **Pennsylvania** | 6,379.6 (5.1%) | 6,379.9 (5.1%) | 0.001 |
| **South Carolina** | 1,255.3 (1.0%) | 1,251.4 (1.0%) | 0.000 |
| **Tennessee** | 855.3 (0.7%) | 855.2 (0.7%) | 0.000 |
| **Texas** | 20,374.9 (16.4%) | 20,298.1 (16.3%) | 0.001 |
| **Utah** | 124.5 (0.1%) | 126.2 (0.1%) | 0.001 |
| **Virginia** | 3,457.4 (2.8%) | 3,455.9 (2.8%) | 0.000 |
| **Vermont** | 148.7 (0.1%) | 149.5 (0.1%) | 0.000 |
| **Washington** | 1,923.1 (1.5%) | 1,926.1 (1.6%) | 0.001 |
| **Wisconsin** | 3,712.1 (3.0%) | 3,718.4 (3.0%) | 0.001 |
| **West Virginia** | 529.7 (0.4%) | 530.4 (0.4%) | 0.000 |
| **Month and year of index date, n (%)** | | | |
| **January 2021** | 2,535.4 (2.0%) | 2,511.3 (2.0%) | 0.001 |
| **February 2021** | 12,007.1 (9.6%) | 12,004.1 (9.7%) | 0.001 |
| **March 2021** | 22,800.8 (18.3%) | 22,683.3 (18.3%) | 0.001 |
| **April 2021** | 43,377.9 (34.8%) | 43,325.1 (34.8%) | 0.001 |
| **May 2021** | 21,334.3 (17.1%) | 21,291.1 (17.1%) | 0.000 |
| **June 2021** | 9,889.0 (7.9%) | 9,882.0 (8.0%) | 0.000 |
| **July 2021** | 5,186.9 (4.2%) | 5,199.8 (4.2%) | 0.001 |

| | | | |
|---|---|---|---|
| **August 2021** | 6,078.5 (4.9%) | 6,041.8 (4.9%) | 0.001 |
| **September 2021** | 1,379.3 (1.1%) | 1,373.3 (1.1%) | 0.000 |
| **State transmission category on index, n (%)** | | | |
| **High transmission state** | 53,810.9 (43.2%) | 53,605.2 (43.1%) | 0.001 |
| **Substantial transmission state** | 34,854.2 (28.0%) | 35,321.6 (28.4%) | 0.010 |
| **Low transmission state** | 35,924.3 (28.8%) | 35,384.9 (28.5%) | 0.008 |
| **Mean (SD) count of hospitalization events, 365 days** | 1.57 (7.76) | 1.56 (7.04) | 0.001 |
| **Mean (SD) count of outpatient events, 365 days** | 34.80 (53.69) | 34.77 (52.84) | 0.001 |
| **Comorbid conditions and scores** | | | |
| **Charlson comorbidity index score, n (%)** | | | |
| **0** | 35,511.1 (28.5%) | 35,474.9 (28.5%) | 0.001 |
| **1** | 24,101.6 (19.3%) | 24,049.4 (19.4%) | 0.000 |
| **2+** | 64,976.6 (52.2%) | 64,787.4 (52.1%) | 0.001 |
| **Frailty score, n (%)** | | | |
| **Robust (0-0.149)** | 89,680.6 (72.0%) | 89,522.7 (72.0%) | 0.001 |
| **Prefrail (0.15-0.249)** | 31,490.1 (25.3%) | 31,379.7 (25.2%) | 0.001 |
| **Mild frailty (0.25-0.349)** | 2,994.2 (2.4%) | 2,987.0 (2.4%) | 0.000 |
| **Moderate to severe frailty (>=0.35)** | 424.3 (0.3%) | 422.3 (0.3%) | 0.000 |
| **Clinical conditions, n (%)** | | | |
| **Alcohol use** | 631.4 (0.5%) | 635.0 (0.5%) | 0.001 |
| **Tobacco use/smoking** | 3,342.2 (2.7%) | 3,345.4 (2.7%) | 0.001 |
| **Obesity (BMI ≥30 kg/m$^2$)** | 2,818.3 (2.3%) | 2,810.4 (2.3%) | 0.000 |

| | | | |
|---|---|---|---|
| **Arrhythmia** | 1,874.0 (1.5%) | 1,873.7 (1.5%) | 0.000 |
| **Asthma** | 1,444.1 (1.2%) | 1,443.0 (1.2%) | 0.000 |
| **Cancer** | 7,780.1 (6.2%) | 7,757.8 (6.2%) | 0.000 |
| **Cardiovascular disease** | 12,595.5 (10.1%) | 12,546.1 (10.1%) | 0.001 |
| **Cerebrovascular disease** | 781.3 (0.6%) | 783.4 (0.6%) | 0.000 |
| **Chronic kidney disease** | 2,614.3 (2.1%) | 2,610.1 (2.1%) | 0.000 |
| **Chronic lung disease** | 2,918.9 (2.3%) | 2,912.5 (2.3%) | 0.000 |
| **Chronic obstructive pulmonary disease** | 2,918.9 (2.3%) | 2,912.5 (2.3%) | 0.000 |
| **Coronary artery disease** | 1,498.4 (1.2%) | 1,496.6 (1.2%) | 0.000 |
| **Dementia** | 170.9 (0.1%) | 168.5 (0.1%) | 0.001 |
| **Diabetes** | 4,994.0 (4.0%) | 4,972.7 (4.0%) | 0.000 |
| **Down syndrome** | 25.2 (0.0%) | 25.6 (0.0%) | 0.000 |
| **Heart failure** | 1,101.0 (0.9%) | 1,094.9 (0.9%) | 0.000 |
| **Hypertension** | 9,791.1 (7.9%) | 9,752.0 (7.8%) | 0.001 |
| **Liver disease** | 1,676.9 (1.4%) | 1,666.9 (1.3%) | 0.000 |
| **Other respiratory viruses (eg, influenza, RSV)** | 3.7 (0.0%) | 4.0 (0.0%) | 0.001 |
| **Sickle cell disease or thalassemia** | 29.0 (0.0%) | 29.6 (0.0%) | 0.000 |
| **Psoriasis** | 766.0 (0.6%) | 763.7 (0.6%) | 0.000 |
| **Psoriatic Arthritis** | 1,164.8 (0.9%) | 1,165.2 (0.9%) | 0.000 |
| **Rheumatoid arthritis** | 1,689.9 (1.4%) | 1,688.2 (1.4%) | 0.000 |
| **Irritable bowel syndrome (IBS)** | 6,342.8 (5.1%) | 6,321.9 (5.1%) | 0.000 |
| **Immunocompromised subgroup** | | | |

| | | | |
|---|---|---|---|
| **Blood Transplant** | 10,071.3 (8.1%) | 10,745.6 (8.6%) | 0.020 |
| **Organ Transplant** | 8,944.8 (7.2%) | 9,223.1 (7.4%) | 0.009 |
| **Active Cancer** | 15,041.5 (12.1%) | 15,844.1 (12.8%) | 0.020 |
| **Primary immunodeficiency** | 32,505.8 (26.1%) | 30,966.7 (24.9%) | 0.027 |
| **HIV** | 25,681.0 (20.6%) | 24,212.0 (19.5%) | 0.028 |
| **Immunosuppressive therapy** | 75,003.0 (60.2%) | 75,067.6 (60.4%) | 0.004 |
| **Number of unique immunotherapy medications over 365 days from baseline** | | | 0.000 |
| **Number of unique immunotherapy medications over 365d baseline, mean (SD)** | 0.92 (0.99) | 0.92 (0.98) | 0.000 |
| **Mena (SD) cumulative duration of immunotherapy** | | | |
| **Topical agent** | 0.00 (0.00) | 0.00 (0.20) | 0.007 |
| **MTOR inhibitor** | 1.75 (23.81) | 2.12 (26.35) | 0.015 |
| **Inosine monophosphate** | 15.71 (70.54) | 16.51 (72.22) | 0.011 |
| **Anti-inflammatory** | 10.37 (52.01) | 10.79 (53.17) | 0.008 |
| **Any calcineurin inhibitor** | 29.56 (90.41) | 29.57 (90.10) | 0.000 |
| **Multiple sclerosis agent** | | | |
| **Multiple sclerosis agent** | 5.90 (44.62) | 6.45 (46.63) | 0.012 |
| **T-lymphocyte immunoglobulin** | 0.00 (0.00) | 0.00 (0.00) | - |
| **JAK inhibitor** | 5.94 (42.38) | 6.00 (42.52) | 0.002 |
| **Antileprotic therapy (thalidomide)** | 0.05 (3.82) | 0.02 (2.38) | 0.007 |
| **Antifibrotic therapy** | 0.10 (5.72) | 0.19 (7.92) | 0.012 |
| **IL-1 inhibitor/IL-1 receptor antagonist** | 0.14 (6.97) | 0.18 (7.79) | 0.005 |

| | | | |
|---|---|---|---|
| **IGF-R inhibitor** | 0.00 (0.35) | 0.00 (0.63) | 0.002 |
| **Any fusion protein** | 1.87 (24.21) | 1.73 (23.08) | 0.006 |
| **Immunosuppressive antimetabolite** | 11.36 (60.14) | 10.88 (58.75) | 0.008 |
| **Any antineoplastic agent** | 46.47 (111.59) | 45.02 (109.78) | 0.013 |
| **Any monoclonal antibody** | 62.93 (126.62) | 61.15 (125.05) | 0.014 |

BMI, body mass index; CED, cohort entry date; CI, confidence interval; Dx, diagnosis; ED, emergency department; IGF-R, insulin-like growth factor receptor; IL, interleukin; JAK, Janus kinase; MTOR, mammalian target of rapamycin; RSV, respiratory syncytial virus; SD, standard deviation.

**Table S8. Subgroup analysis: incidence and VE of mRNA-1273 and BNT162b2 on medically-attended breakthrough COVID-19 diagnosis among immunocompromised adults**

| Subgroup | No. of individuals | | No. of events | | Rate (95% CI) | | HR (95% CI) |
|---|---|---|---|---|---|---|---|
| | mRNA-1273 | BNT162b2 | mRNA-1273 | BNT162b2 | mRNA-1273 | BNT162b2 | mRNA-1273 vs BNT162b2 |
| **Age group** | | | | | | | |
| **<50 y** | 21,539 | 27,474 | 239 | 342 | 27.98 (24.60-31.82) | 33.29 (29.91-37.05) | 0.84 (0.71-0.99) |
| **50-64 y** | 29,938 | 33,935 | 308 | 426 | 24.77 (22.12-27.73) | 30.51 (27.72-33.59) | 0.81 (0.70-0.94) |
| **≥65 y** | 6,223 | 5,348 | 70 | 72 | 22.72 (17.82-28.98) | 26.26 (20.58-33.50) | 0.87 (0.61-1.22) |
| **Time period second dose received** | | | | | | | |
| **Second dose Q1 2021** | 17,737 | 19,413 | 286 | 384 | 30.27 (26.91-34.05) | 36.42 (32.91-40.30) | 0.83 (0.71-0.97) |
| **Second dose Q2 2021** | 35,469 | 39,227 | 322 | 435 | 23.41 (20.95-26.16) | 28.38 (25.80-31.23) | 0.81 (0.70-0.94) |
| **Second dose Q3 2021** | 4,494 | 8,117 | 9 | 21 | 14.36 (7.42-27.77) | 18.79 (12.11-29.17) | 0.78 (0.35-1.72) |
| **Transmission state CED** | | | | | | | |
| **High transmission state on CED** | 23,558 | 29,933 | 243 | 329 | 23.66 (20.81-26.90) | 27.46 (24.63-30.62) | 0.86 (0.73-1.02) |
| **Substantial transmission state on CED** | 16,203 | 18,945 | 206 | 309 | 29.51 (25.66-33.93) | 36.35 (32.44-40.73) | 0.80 (0.66-0.95) |
| **Low transmission state on CED** | 17,939 | 17,879 | 168 | 202 | 25.40 (21.75-29.67) | 31.03 (26.94-35.73) | 0.81 (0.66-1.00) |
| **Comorbidity, therapy** | | | | | | | |
| **Blood/stem cell transplant** | 4,536 | 5,867 | 83 | 103 | 44.06 (35.39-54.85) | 45.80 (37.63-55.75) | 0.96 (0.72-1.29) |
| **Solid organ transplant** | 4,029 | 5,043 | 92 | 117 | 56.85 (46.19-69.97) | 58.94 (48.98-70.92) | 0.96 (0.73-1.27) |
| **Active cancer** | 7,186 | 8,277 | 61 | 100 | 19.13 (14.81-24.70) | 27.87 (22.85-34.00) | 0.69 (0.50-0.95) |
| **Primary immunodeficiency syndrome** | 15,704 | 16,099 | 114 | 165 | 18.84 (15.60-22.75) | 25.30 (21.67-29.54) | 0.74 (0.58-0.95) |
| **HIV** | 12,510 | 12,505 | 93 | 125 | 19.76 (16.02-24.37) | 24.87 (20.81-29.71) | 0.79 (0.60-1.05) |
| **Immunosuppressive therapy** | 33,915 | 41,029 | 432 | 565 | 29.99 (27.26-32.99) | 34.14 (31.41-37.12) | 0.88 (0.77-1.00) |

CED, cohort entry date; CI, confidence interval; HR, hazard ratio

**Table S9. Subgroup analysis: incidence and VE of mRNA-1273 and BNT162b2 on COVID-19 hospitalizations among immunocompromised adults**

| Subgroup | No. of individuals | | No. of events | | Rate (95% CI) | | HR (95% CI) |
|---|---|---|---|---|---|---|---|
| | mRNA-1273 | BNT162b2 | mRNA-1273 | BNT162b2 | mRNA-1273 | BNT162b2 | mRNA-1273 vs BNT162b2 |
| **Age group** | | | | | | | |
| **<50 y** | 21,539 | 27,474 | 22 | 36 | 2.66 (1.73-4.07) | 3.51 (2.52-4.90) | 0.76 (0.44-1.31) |
| **50-64 y** | 29,938 | 33,935 | 47 | 70 | 3.64 (2.73-4.86) | 4.99 (3.94-6.34) | 0.73 (0.50-1.06) |
| **≥65+ y** | 6,223 | 5,348 | 21 | 19 | 6.83 (4.40-10.61) | 7.48 (4.61-12.15) | 0.91 (0.47-1.76) |
| **Time period second dose received** | | | | | | | |
| **Second dose Q1 2021** | 17,737 | 19,413 | 38 | 62 | 4.02 (2.91-5.55) | 5.87 (4.55-7.59) | 0.69 (0.45-1.03) |
| **Second dose Q2 2021** | 35,469 | 39,227 | 49 | 59 | 3.46 (2.60-4.60) | 3.88 (2.98-5.03) | 0.88 (0.60-1.30) |
| **Second dose Q3 2021** | 4,494 | 8,117 | 3 | 4 | 5.16 (1.64-16.16) | 3.96 (1.40-11.21) | 1.20 (0.25-5.67) |
| **Transmission state CED** | | | | | | | |
| **High transmission state on CED** | 23,558 | 29,933 | 33 | 56 | 3.21 (2.27-4.54) | 4.62 (3.54-6.02) | 0.69 (0.45-1.07) |
| **Substantial transmission state on CED** | 16,203 | 18,945 | 26 | 45 | 3.51 (2.37-5.20) | 5.34 (3.93-7.26) | 0.64 (0.39-1.06) |
| **Low transmission state on CED** | 17,939 | 17,879 | 31 | 24 | 4.41 (3.08-6.32) | 3.79 (2.51-5.73) | 1.16 (0.67-2.01) |
| **Comorbidity, therapy** | | | | | | | |
| **Blood/stem cell transplant** | 4,536 | 5,867 | 23 | 25 | 11.76 (7.74-17.86) | 10.66 (7.14-15.92) | 1.10 (0.62-1.96) |
| **Solid organ transplant** | 4,029 | 5,043 | 28 | 35 | 16.30 (11.15-23.84) | 17.42 (12.39-24.49) | 0.93 (0.56-1.56) |
| **Active cancer** | 7,186 | 8,277 | 8 | 18 | 2.21 (1.09-4.48) | 5.17 (3.24-8.27) | 0.43 (0.18-1.00) |
| **Primary immunodeficiency syndrome** | 15,704 | 16,099 | 14 | 20 | 2.39 (1.39-4.11) | 3.11 (1.99-4.88) | 0.77 (0.38-1.56) |
| **HIV** | 12,510 | 12,505 | 9 | 15 | 1.79 (0.91-3.51) | 2.85 (1.70-4.78) | 0.63 (0.27-1.47) |
| **Immunosuppressive therapy** | 33,915 | 41,029 | 65 | 81 | 4.34 (3.39-5.55) | 4.97 (3.97-6.21) | 0.87 (0.63-1.22) |

CED, cohort entry date; CI, confidence interval; HR, hazard ratio.

**Table S10. Sensitivity analysis: incidence and VE of mRNA-1273 and BNT162b2 on medically-attended breakthrough COVID-19 diagnosis among immunocompromised**

| | No. of individuals | | No. of events | | Rate (95% CI) | | HR (95% CI) |
|---|---|---|---|---|---|---|---|
| Analysis type | mRNA-1273 | BNT162b2 | mRNA-1273 | BNT162b2 | mRNA-1273 | BNT162b2 | mRNA-1273 vs BNT162b2 |
| **No truncation** | 57,777 | 66,879 | 620 | 842 | 17.97 (16.58-19.46) | 21.44 (20.02-22.96) | 0.83 (0.75-0.93) |
| **Alternative medically-attended COVID-19 definition** | 57,700 | 66,757 | 264 | 387 | 11.08 (9.80-12.52) | 14.18 (12.82-15.68) | 0.78 (0.67-0.92) |
| **Open claims for cohort entry and outcome capture** | 201,365 | 187,600 | 2,253 | 2,807 | 27.31 (26.19-28.49) | 37.06 (35.35-38.85) | 0.74 (0.70-0.79) |
| **Open claims for outcome capture** | 124,588 | 124,313 | 732 | 1,002 | 30.68 (28.51-33.02) | 37.42 (35.14-39.84) | 0.82 (0.74-0.90) |
| **1:1 propensity score matching** | 52,729 | 52,729 | 571 | 678 | 26.05 (23.91-28.11) | 30.93 (28.72-33.31) | 0.84 (0.75-0.94) |

CI, confidence interval; HR, hazard ratio.

**Table S11. Sensitivity analysis: incidence and VE of mRNA-1273 and BNT162b2 on COVID-19 hospitalization among immunocompromised adults**

| | No. of individuals | | No. of events | | Rate (95% CI) | | HR (95% CI) |
|---|---|---|---|---|---|---|---|
| **Analysis type** | **mRNA-1273** | **BNT162b2** | **mRNA-1273** | **BNT162b2** | **mRNA-1273** | **BNT162b2** | **mRNA-1273 vs BNT162b2** |
| **No truncation** | 57,777 | 66,879 | 90 | 125 | 2.53 (2.05-3.12) | 3.22 (2.69-3.85) | 0.78 (0.59-1.03) |
| **Alternative medically-attended COVID-19 definition** | 57,700 | 66,757 | N/A | N/A | N/A | N/A | N/A |
| **Open claims for cohort entry and outcome capture** | 201,365 | 187,600 | 386 | 488 | 4.46 (4.02-4.93) | 6.73 (6.14-7.38) | 0.66 (0.58-0.76) |
| **Open claims for outcome capture** | 124,588 | 124,313 | 113 | 169 | 4.58 (3.80-5.52) | 6.33 (5.43-7.39) | 0.72 (0.57-0.92) |
| **1:1 propensity score matching** | 52,729 | 52,729 | 84 | 97 | 3.81 (3.08-4.72) | 4.40 (3.61-5.37) | 0.87 (0.65-1.16) |

CI, confidence interval; HR, hazard ratio; N/A, not available.

**Supplementary Figures**

**Figure S1. Equations used in the bias analysis**

| **mRNA-1273** | | |
|---|---|---|
| | **PCR+** | **PCR-** |
| **ICD+** | aM | bM |
| **ICD-** | cM | dM |

**Sensitivity= aM/(aM+cM)**
**Specificity=dM/(bM+dM)**
**PPV(Moderna) = aM / (aM+bM)**

| **BNT162b2** | | |
|---|---|---|
| | **PCR+** | **PCR-** |
| **ICD+** | aP | bP |
| **ICD-** | cP | dP |

**Sensitivity= aP/(aP+cP)**
**Specificity=dP/(bP+dP)**
**PPV (Pfizer) = aP / (aP+bP)**

**Figure S2. A) Propensity score distribution pre-weighting and B) Propensity score distribution post-weighting**

**A**

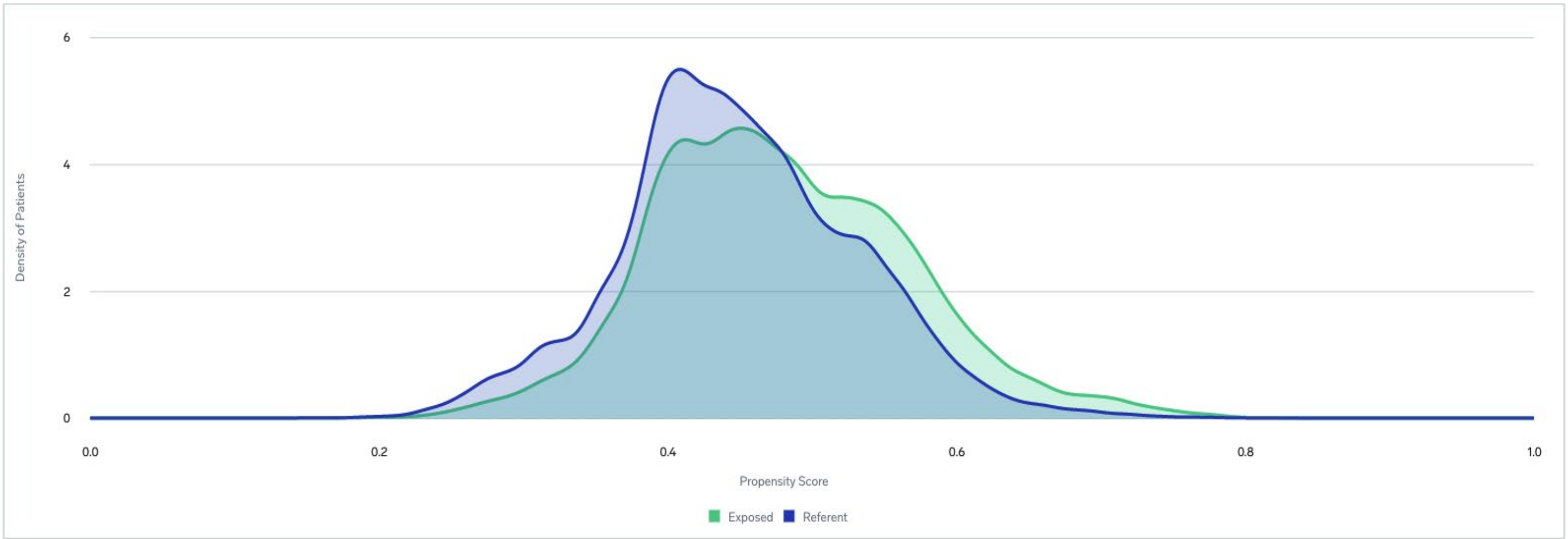


**B**

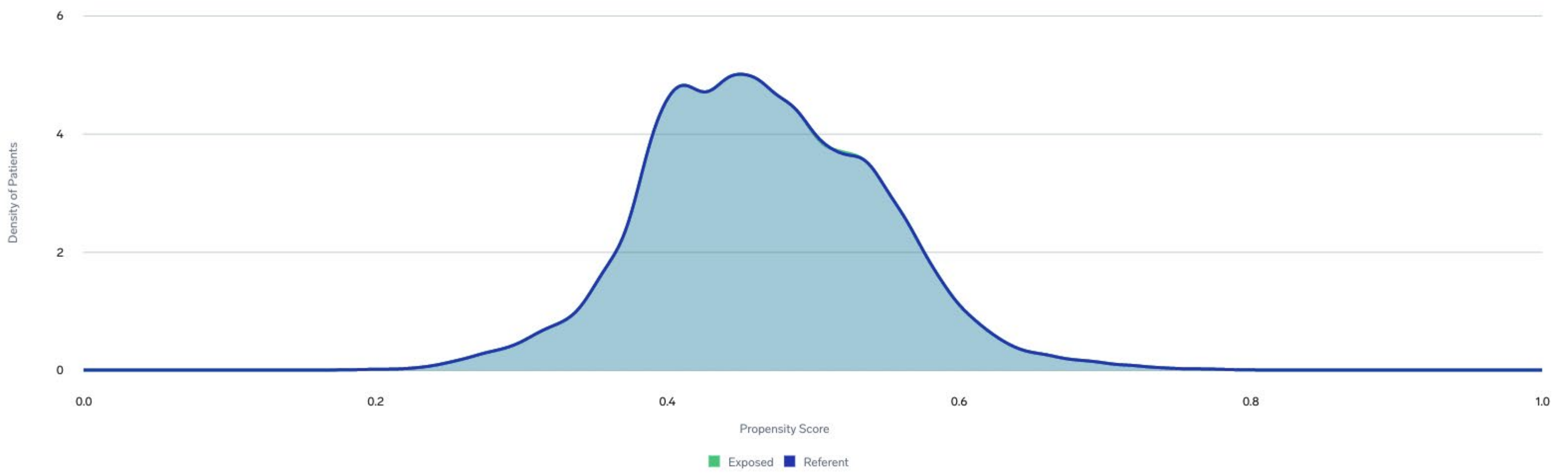
